# Multi-ancestry genome-wide meta-analysis of 56,241 individuals identifies *LRRC4C, LHX5-AS1* and nominates ancestry-specific loci *PTPRK*, *GRB14*, and *KIAA0825* as novel risk loci for Alzheimer’s disease: the Alzheimer’s Disease Genetics Consortium

**DOI:** 10.1101/2023.07.06.23292311

**Authors:** Farid Rajabli, Penelope Benchek, Giuseppe Tosto, Nicholas Kushch, Jin Sha, Katrina Bazemore, Congcong Zhu, Wan-Ping Lee, Jacob Haut, Kara L. Hamilton-Nelson, Nicholas R. Wheeler, Yi Zhao, John J. Farrell, Michelle A. Grunin, Yuk Yee Leung, Pavel P. Kuksa, Donghe Li, Eder Lucio da Fonseca, Jesse B. Mez, Ellen L. Palmer, Jagan Pillai, Richard M. Sherva, Yeunjoo E. Song, Xiaoling Zhang, Taha Iqbal, Omkar Pathak, Otto Valladares, Amanda B. Kuzma, Erin Abner, Perrie M. Adams, Alyssa Aguirre, Marilyn S. Albert, Roger L. Albin, Mariet Allen, Lisa Alvarez, Liana G. Apostolova, Steven E. Arnold, Sanjay Asthana, Craig S. Atwood, Gayle Ayres, Clinton T. Baldwin, Robert C. Barber, Lisa L. Barnes, Sandra Barral, Thomas G. Beach, James T. Becker, Gary W. Beecham, Duane Beekly, Bruno A. Benitez, David Bennett, John Bertelson, Thomas D. Bird, Deborah Blacker, Bradley F. Boeve, James D. Bowen, Adam Boxer, James Brewer, James R. Burke, Jeffrey M. Burns, Joseph D. Buxbaum, Nigel J. Cairns, Laura B. Cantwell, Chuanhai Cao, Christopher S. Carlson, Cynthia M. Carlsson, Regina M. Carney, Minerva M. Carrasquillo, Scott Chasse, Marie-Francoise Chesselet, Nathaniel A. Chin, Helena C. Chui, Jaeyoon Chung, Suzanne Craft, Paul K. Crane, David H. Cribbs, Elizabeth A. Crocco, Carlos Cruchaga, Michael L. Cuccaro, Munro Cullum, Eveleen Darby, Barbara Davis, Philip L. De Jager, Charles DeCarli, John DeToledo, Malcolm Dick, Dennis W. Dickson, Beth A. Dombroski, Rachelle S. Doody, Ranjan Duara, NIlüfer Ertekin-Taner, Denis A. Evans, Kelley M. Faber, Thomas J. Fairchild, Kenneth B. Fallon, David W. Fardo, Martin R. Farlow, Victoria Fernandez-Hernandez, Steven Ferris, Tatiana M. Foroud, Matthew P. Frosch, Brian Fulton-Howard, Douglas R. Galasko, Adriana Gamboa, Marla Gearing, Daniel H. Geschwind, Bernardino Ghetti, John R. Gilbert, Alison M. Goate, Thomas J. Grabowski, Neill R. Graff-Radford, Robert C. Green, John H. Growdon, Hakon Hakonarson, James Hall, Ronald L. Hamilton, Oscar Harari, John Hardy, Lindy E. Harrell, Elizabeth Head, Victor W. Henderson, Michelle Hernandez, Timothy Hohman, Lawrence S. Honig, Ryan M. Huebinger, Matthew J. Huentelman, Christine M. Hulette, Bradley T. Hyman, Linda S. Hynan, Laura Ibanez, Gail P. Jarvik, Suman Jayadev, Lee-Way Jin, Kim Johnson, Leigh Johnson, M. Ilyas Kamboh, Anna M. Karydas, Mindy J. Katz, John S. Kauwe, Jeffrey A. Kaye, C. Dirk Keene, Aisha Khaleeq, Ronald Kim, Janice Knebl, Neil W. Kowall, Joel H. Kramer, Walter A. Kukull, Frank M. LaFerla, James J. Lah, Eric B. Larson, Alan Lerner, James B. Leverenz, Allan I. Levey, Andrew P. Lieberman, Richard B. Lipton, Mark Logue, Oscar L. Lopez, Kathryn L. Lunetta, Constantine G. Lyketsos, Douglas Mains, Flanagan E. Margaret, Daniel C. Marson, Eden R R. Martin, Frank Martiniuk, Deborah C. Mash, Eliezer Masliah, Paul Massman, Arjun Masurkar, Wayne C. McCormick, Susan M. McCurry, Andrew N. McDavid, Stefan McDonough, Ann C. McKee, Marsel Mesulam, Bruce L. Miller, Carol A. Miller, Joshua W. Miller, Thomas J. Montine, Edwin S. Monuki, John C. Morris, Shubhabrata Mukherjee, Amanda J. Myers, Trung Nguyen, Sid O’Bryant, John M. Olichney, Marcia Ory, Raymond Palmer, Joseph E. Parisi, Henry L. Paulson, Valory Pavlik, David Paydarfar, Victoria Perez, Elaine Peskind, Ronald C. Petersen, Aimee Pierce, Marsha Polk, Wayne W. Poon, Huntington Potter, Liming Qu, Mary Quiceno, Joseph F. Quinn, Ashok Raj, Murray Raskind, Eric M. Reiman, Barry Reisberg, Joan S. Reisch, John M. Ringman, Erik D. Roberson, Monica Rodriguear, Ekaterina Rogaeva, Howard J. Rosen, Roger N. Rosenberg, Donald R. Royall, Mark A. Sager, Mary Sano, Andrew J. Saykin, Julie A. Schneider, Lon S. Schneider, William W. Seeley, Susan H. Slifer, Scott Small, Amanda G. Smith, Janet P. Smith, Joshua A. Sonnen, Salvatore Spina, Peter St George-Hyslop, Robert A. Stern, Alan B. Stevens, Stephen M. Strittmatter, David Sultzer, Russell H. Swerdlow, Rudolph E. Tanzi, Jeffrey L. Tilson, John Q. Trojanowski, Juan C. Troncoso, Debby W. Tsuang, Vivianna M. Van Deerlin, Linda J. van Eldik, Jeffery M. Vance, Badri N. Vardarajan, Robert Vassar, Harry V. Vinters, Jean-Paul Vonsattel, Sandra Weintraub, Kathleen A. Welsh-Bohmer, Patrice L. Whitehead, Ellen M. Wijsman, Kirk C. Wilhelmsen, Benjamin Williams, Jennifer Williamson, Henrik Wilms, Thomas S. Wingo, Thomas Wisniewski, Randall L. Woltjer, Martin Woon, Clinton B. Wright, Chuang-Kuo Wu, Steven G. Younkin, Chang-En Yu, Lei Yu, Xiongwei Zhu, Brian W. Kunkle, William S. Bush, Li-San Wang, Lindsay A. Farrer, Jonathan L. Haines, Richard Mayeux, Margaret A. Pericak-Vance, Gerard D. Schellenberg, Gyungah R. Jun, Christiane Reitz, Adam C. Naj

## Abstract

Limited ancestral diversity has impaired our ability to detect risk variants more prevalent in non-European ancestry groups in genome-wide association studies (GWAS). We constructed and analyzed a multi-ancestry GWAS dataset in the Alzheimer’s Disease (AD) Genetics Consortium (ADGC) to test for novel shared and ancestry-specific AD susceptibility loci and evaluate underlying genetic architecture in 37,382 non-Hispanic White (NHW), 6,728 African American, 8,899 Hispanic (HIS), and 3,232 East Asian individuals, performing within-ancestry fixed-effects meta-analysis followed by a cross-ancestry random-effects meta-analysis. We identified 13 loci with cross-ancestry associations including known loci at/near *CR1*, *BIN1*, *TREM2*, *CD2AP*, *PTK2B*, *CLU*, *SHARPIN*, *MS4A6A*, *PICALM*, *ABCA7*, *APOE* and two novel loci not previously reported at 11p12 (*LRRC4C*) and 12q24.13 (*LHX5-AS1*). Reflecting the power of diverse ancestry in GWAS, we observed the *SHARPIN* locus using 7.1% the sample size of the original discovering single-ancestry GWAS (n=788,989). We additionally identified three GWS ancestry-specific loci at/near (*PTPRK* (*P*=2.4×10^-8^) and *GRB14* (*P*=1.7×10^-8^) in HIS), and *KIAA0825* (*P*=2.9×10^-8^ in NHW). Pathway analysis implicated multiple amyloid regulation pathways (strongest with *P*_adjusted_=1.6×10^-4^) and the classical complement pathway (*P*_adjusted_=1.3×10^-3^). Genes at/near our novel loci have known roles in neuronal development (*LRRC4C, LHX5-AS1*, and *PTPRK*) and insulin receptor activity regulation (*GRB14*). These findings provide compelling support for using traditionally-underrepresented populations for gene discovery, even with smaller sample sizes.

## INTRODUCTION

Alzheimer’s disease (AD) affects over a third of people aged 80 years or older^1^, with AD prevalence continuing to increase with the growth in the global population of elderly. Currently, there are approximately 50 million persons with AD worldwide, projected to increase to 135-200 million by 2050^1^. Coupled with the facts that there are limited therapeutic interventions for AD and that this disease affects individuals of all ancestries, the expanding burden of AD makes this disease an urgent global public health crisis. Genome-wide association studies (GWAS) have identified over 75 risk loci associated with risk of AD and related dementias^2-5^, providing critical insights into molecular mechanisms underlying disease development. These studies, however, have been conducted predominantly in populations of European ancestry, although it is clear that ethnobiological origin (i.e. genetic ancestry) impacts genotypic risk, with rare variants in particular usually being shared only between genetically closely-related populations. This has limited the ability to identify ancestry-specific variants and loci^6-8^, determine shared genetic risk and protective factors across ancestrally-diverse populations^9^, and to capitalize on differential linkage disequilibrium (LD) structures across varied ancestral genetic backgrounds to improve fine-mapping of causal loci^10-13^. Thus, progress towards genetic prediction and precision medicine is significantly hampered by lack of data from non-European populations^14^.

Though concepts of race, ethnicity, and ancestry are often conflated^15^, our study focuses on genetic ancestry as shared genetic background through biological descent from common ancestors, where common ancestors may be defined over varying windows of generational time. For the purposes of this analysis, this may include individuals of more genetically homogeneous groups such as European (non-Hispanic whites (NHW)) and East Asian (EAS) ancestry groups, as well as groups with more recent admixture of previously ancestrally-divergent groups, such as African Americans (AFA), whose genetic ancestries includes admixture from African and European ancestries, and Hispanics (HIS), whose genetic ancestries include highly variable levels of admixture predominantly between Amerindian, European, and African ancestries and with distinctly different admixture patterns geographically.

To identify novel loci associated across ancestrally-diverse populations and explore differences in genetic association across these groups, the ADGC initiated the first large-scale multi-ancestry GWAS meta-analysis with direct genotyping and either clinical evaluation of, or pathological confirmation of, AD status on over 56,000 individuals of diverse ancestral backgrounds, including 6,728 AFA, 8,899 HIS, 3,232 EAS, and 37,382 NHW participants. We analyzed imputed genotypes derived from the multi-ancestry Trans-Omics for Precision Medicine (TOPMed) R5 haplotype reference panel (308,107,085 variants from 97,256 sequenced individuals). We conducted a two-stage, cross-ancestry GWAS meta-analysis of AD followed by secondary analyses including ancestry-aware fine-mapping to understand genetic architecture underlying novel genetic loci and to identify shared and distinct differences of AD risk and protective loci in ancestrally-diverse populations.

## RESULTS

The ADGC assembled the largest multi-ancestry collection of AD samples to date, containing 24,388 participants with AD and 31,853 cognitively unimpaired individuals passing quality control (QC) from 41 NHW, 12 AFA, eight HIS, and five EAS datasets (**Supplementary Table 1**). Notably, in contrast to several previously reported studies^4,5^, all samples utilized in these analyses were directly genotyped and either clinically assessed or pathologically evaluated, which significantly increased the precision of the analyses. Descriptions of recruitment and diagnosis of AD for newly added and previously available datasets are reported in the **Supplementary Note,** and sample sizes and genome-wide single nucleotide polymorphism (SNP) array platforms for the 66 individual datasets are reported in **Supplementary Table 2**. We excluded individuals younger than age 65 years at the time of censoring (age-at-onset [AAO] of symptoms for AD cases or age-at-last-examination/ age-at-death [AAE] for cognitively-normal elders [CNEs]). Intact cognition among living CNEs was confirmed with cognitive evaluations and absence of AD pathology among deceased CNEs.

### Cross-Ancestry Genome-wide Association Meta-analysis

We first conducted a genome-wide association study (GWAS) for AD separately in each dataset using the two covariate adjustment models (without and with adjustment for *APOE* □4 dosage). Subsequently, within each model, we performed a fixed-effects meta-analysis to combine results across datasets within each ancestry (NHW, HIS, AFA, and EAS) (**Supplementary Tables 3-6, Supplementary Figures 1a-d** and **2a-d**). Q-Q plots for ancestry-specific GWAS results (**Supplementary Figures 3a-d** and **4a-d**) for both models in HIS, AFA, and EAS showed no genomic inflation (λ=0.96-1.00), while those in NHW in the *APOE* □4- unadjusted model showed moderate genomic inflation (λ=1.09; after *APOE* □ 4 adjustment, λ=1.06) (**Supplementary Figure 3a**). After cross-ancestry meta-analysis without (**Figure 1**) and with *APOE* □4 adjustment (**Supplementary Figure 5**, **Supplementary Table 7**) allowing for between-ancestry heterogeneity, we observed modest genomic deflation for either model (λ=0.94-0.95) (**Supplementary Figure 6**).

**Figure 1.**
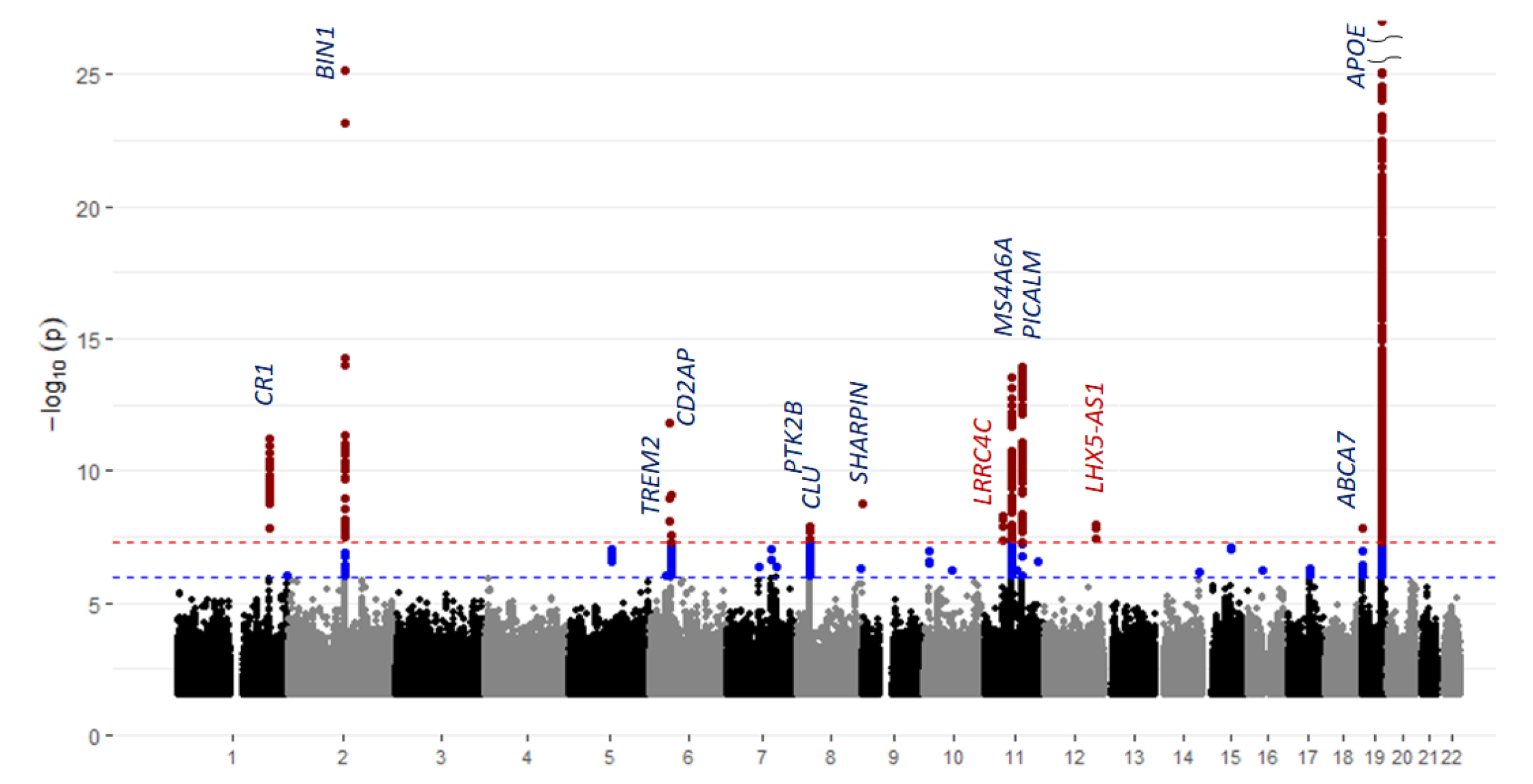
Manhattan plot of cross-ancestry meta-analyses of genome-wide associations estimated with covariate adjustment for age-at-onset (cases)/age-at-last-exam (controls), sex, and PCs for population substructure.

**Figure 2.**
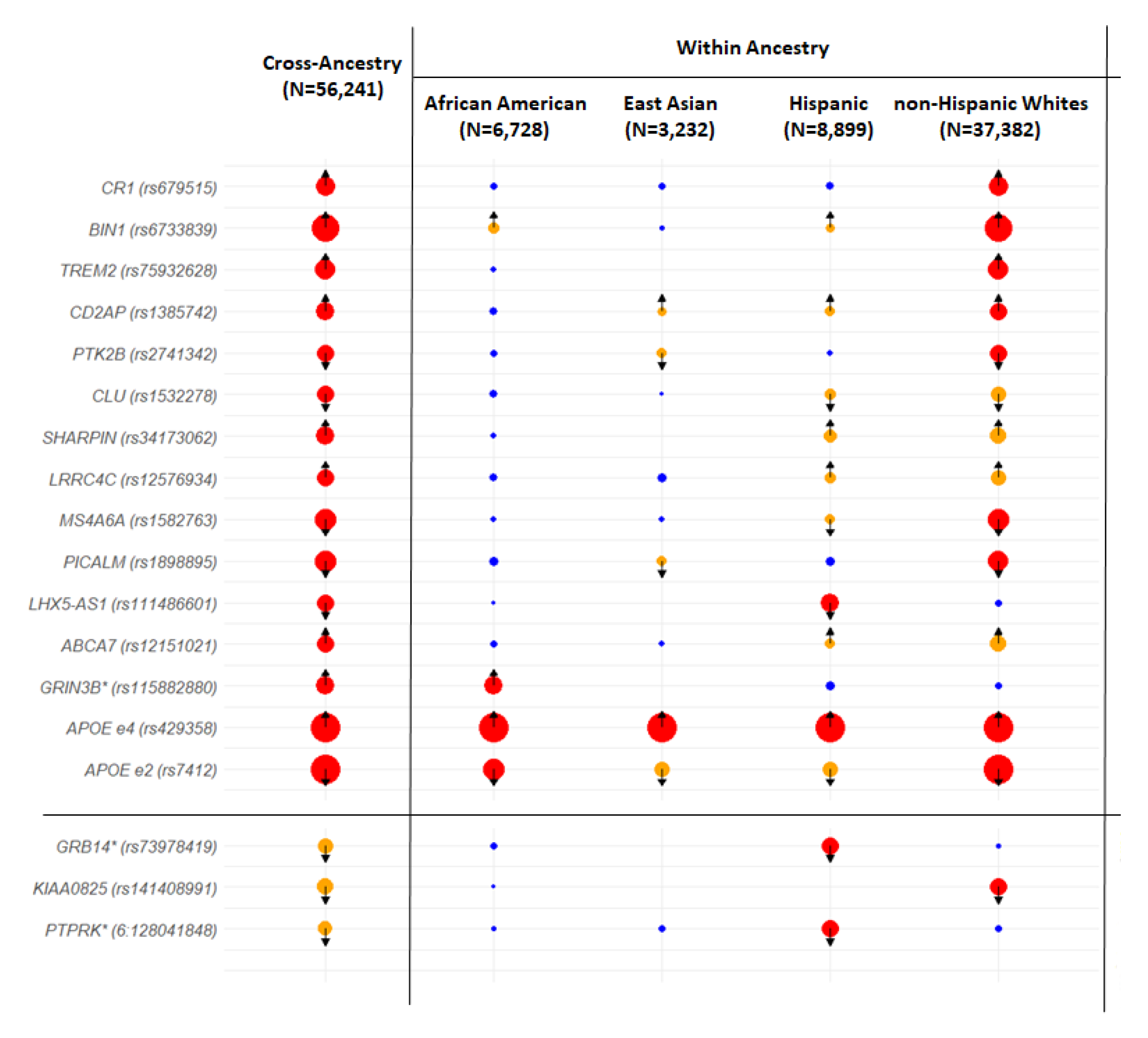
**Ancestry comparison plot of the multi-ancestry GWAS associations.** The plot displays results for 18 loci associated with AD risk within and across multiple ancestries. Circle size corresponds to the significance level, -log_10_(*P*-value), and circle color represents the significance threshold: red, *P*<5×10^-8^; orange, 0.05>*P*>5×10^-8^; and blue, *P*>0.05. Arrows represent the direction of effect of the risk allele, with “up” indicating increased risk and down decreased risk. Asterisks (“*”) indicate the association was found in *APOE* ε4-adjusted models.

#### *APOE* □*4-Unadjusted Model*

In cross-ancestry meta-analysis under the *APOE* □4-unadjusted model, two novel genome-wide significant (GWS; *P*≤5×10^−8^) variants were detected near the genes *LRRC4C* on chromosome 11 (rs12576934: odds ratio [OR]=1.12, 95% confidence interval [95% CI]=1.08- 1.16, *P*=5.4×10^−9^) and *LHX5-AS1* on chromosome 12 (rs111486601; OR=0.63, 95% CI=0.32-1.23, *P*=1.1×10^−8^) (**Figures 1-2** and **Table 1**). We observed the association signal in the *LRRC4C* locus across multiple ancestries including NHW (*P*=1.8×10^−6^), HIS (*P*=7.6×10^−3^), and EAS (*P*=0.06) (**Figure 3** and **Table 1**) with the minor allele C (range of minor allele frequencies [MAFs]=0.17-0.33) increasing risk in all ancestries (ORs from 1.11 to 1.14; **Table 1**). In contrast, ancestry-specific evaluation of rs111486601 in the *LHX5-AS1* region revealed the association signal to be driven predominantly by HIS (MAF=0.04, OR=0.44, 95% CI=0.34-0.57, *P*=4.8×10^−10^) (**Figure 4** and **Table 1**).

**Figure 3.**
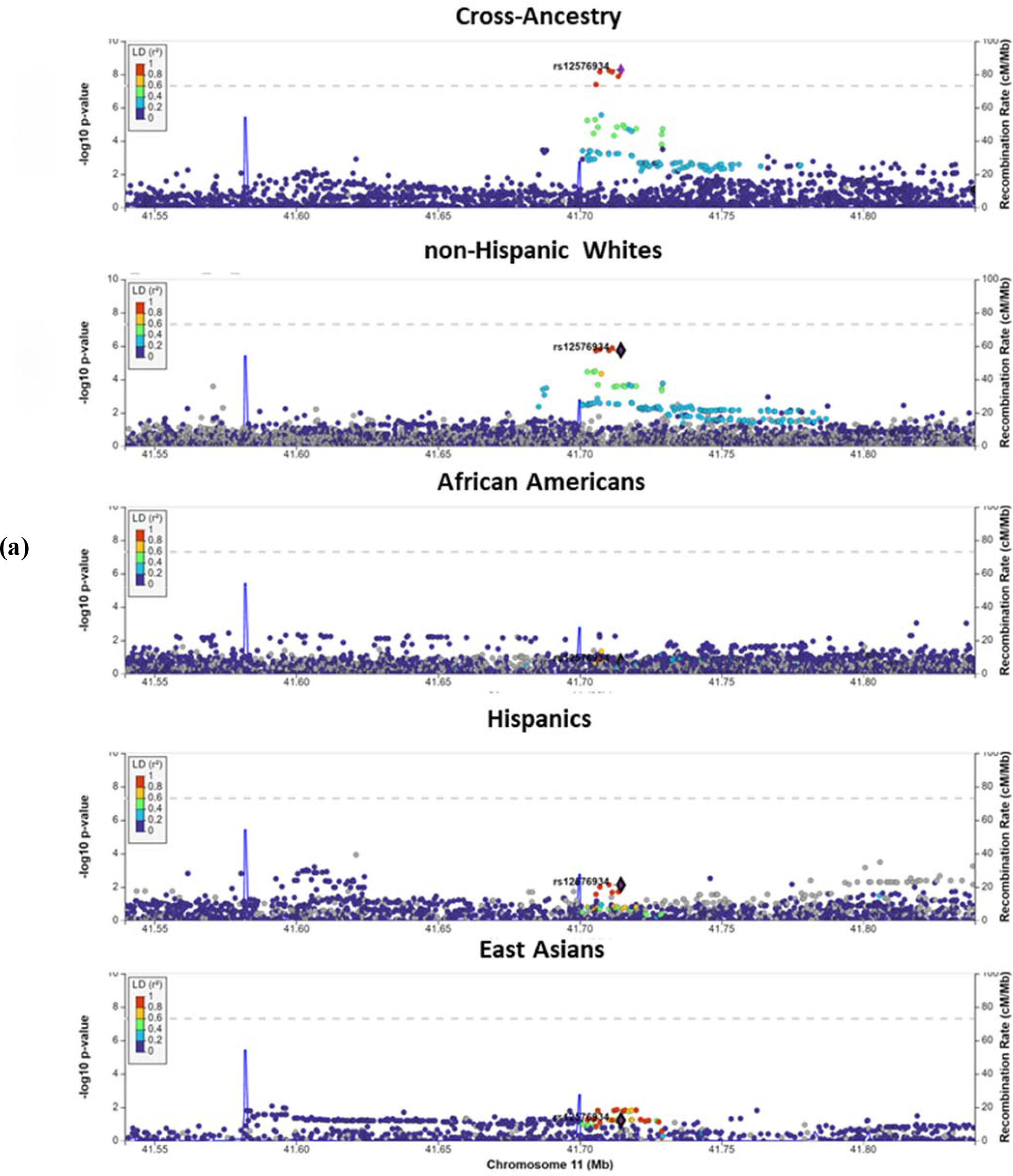
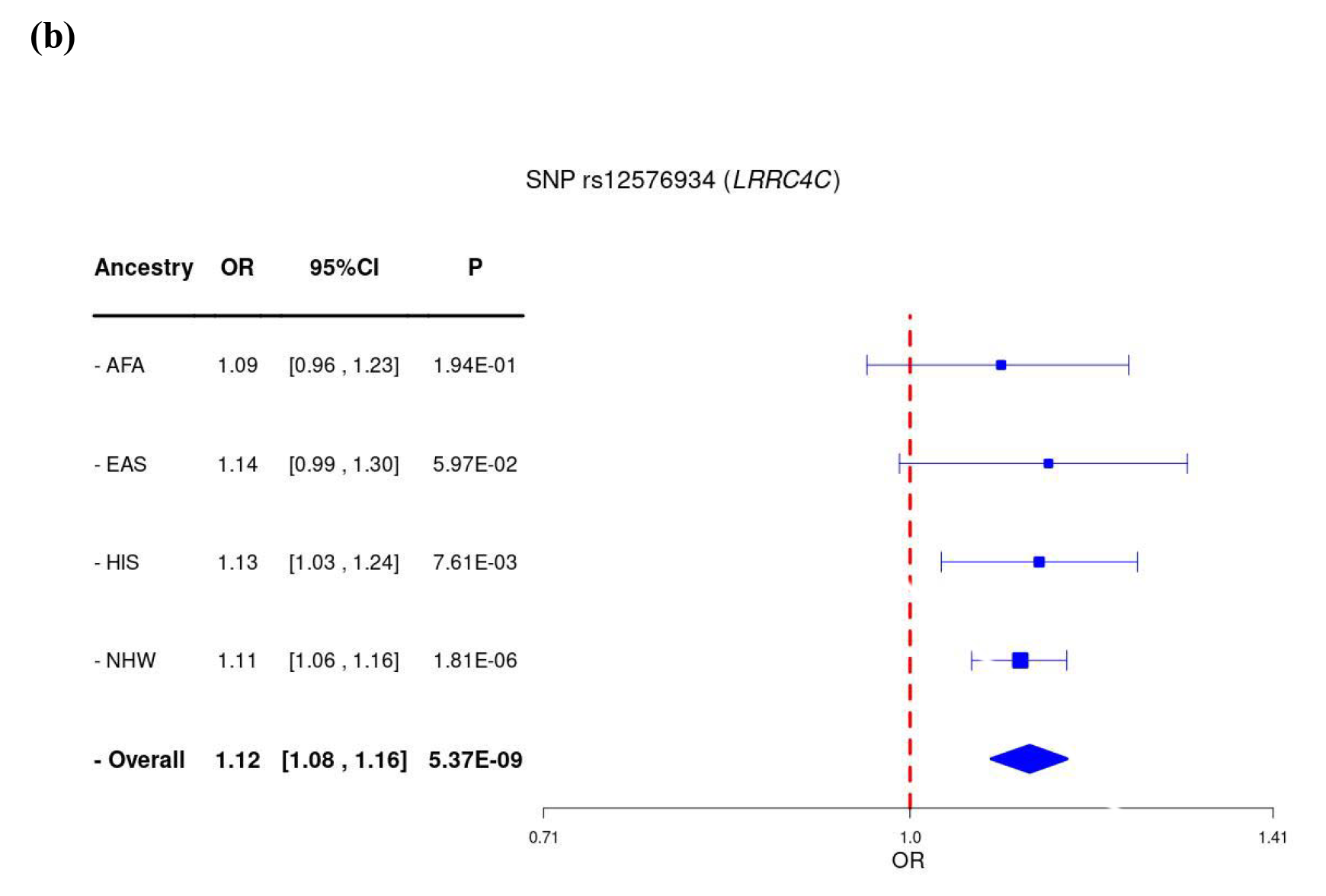
**(a)** Regional association plots and **(b)** Forest plot for the novel *LRRC4C* locus cross-ancestry and within-ancestry associations with AD depicting all associations within ±500□kb of the intergenic lead variant, rs12576934.

**Figure 4.**
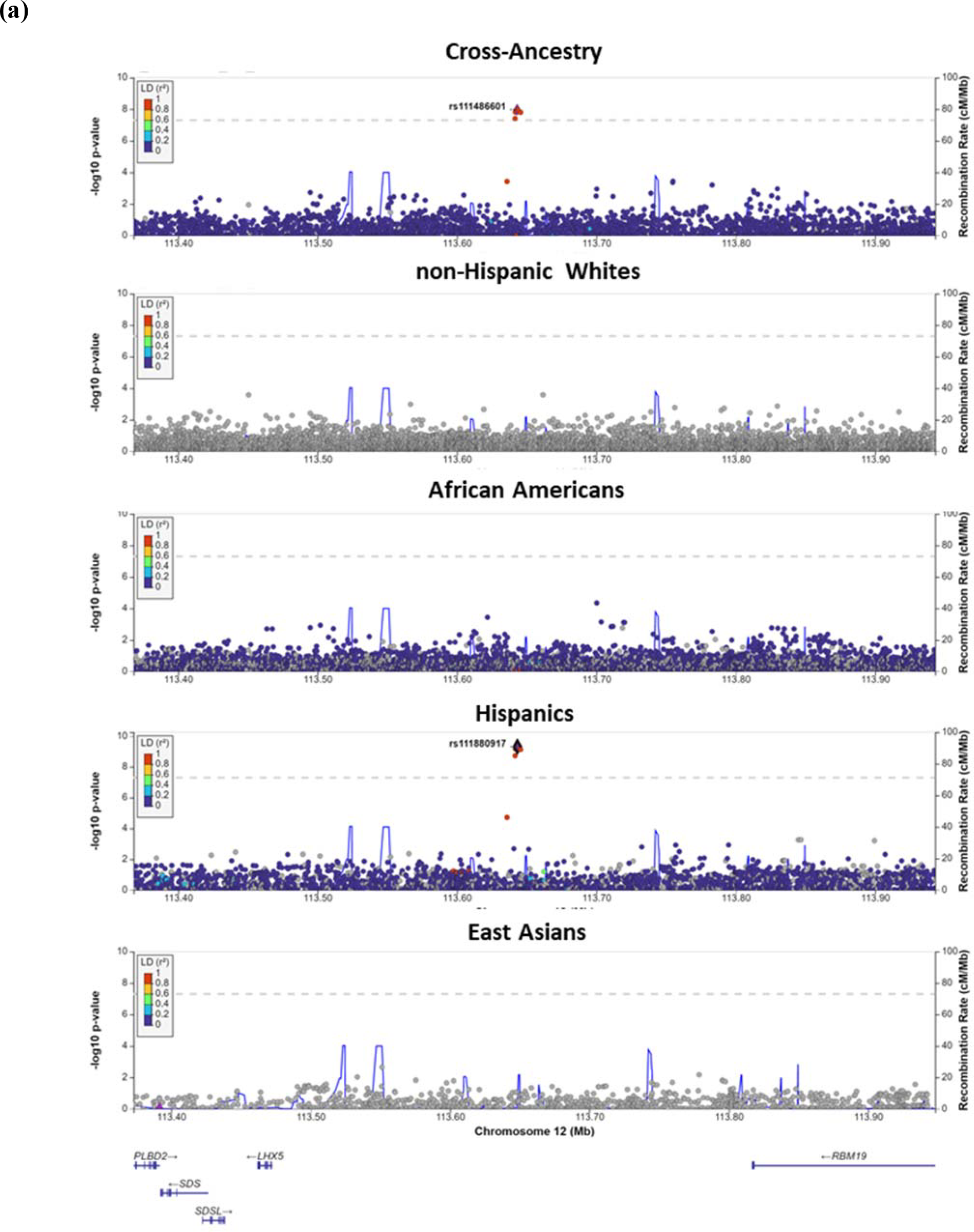
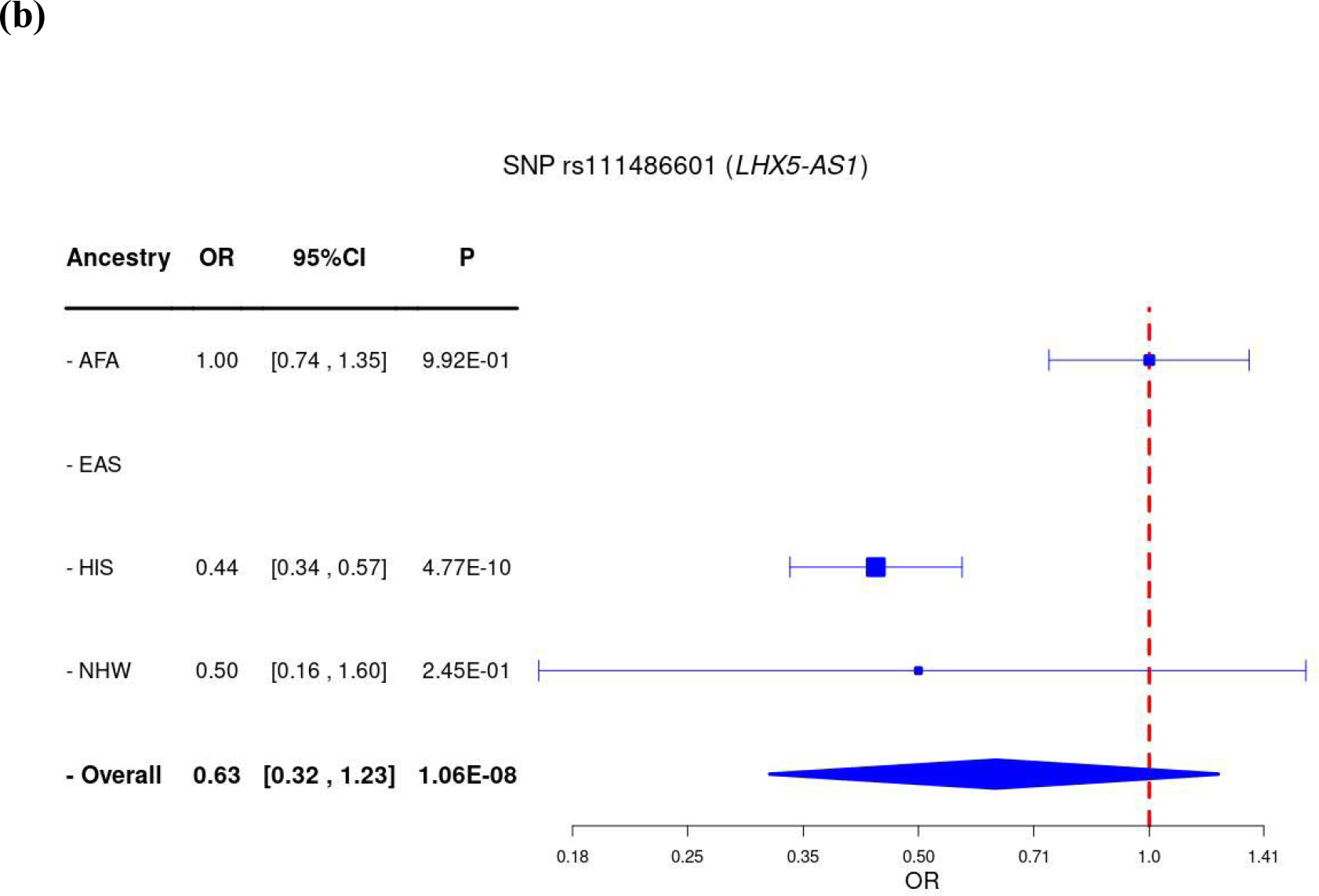
**(a)** Regional association plots and **(b)** Forest plot for the novel *LHX5-AS1* locus cross-ancestry and within-ancestry associations with AD depicting all associations within ±500□kb of the intergenic lead variant, rs111486601.

**Figure 4.**
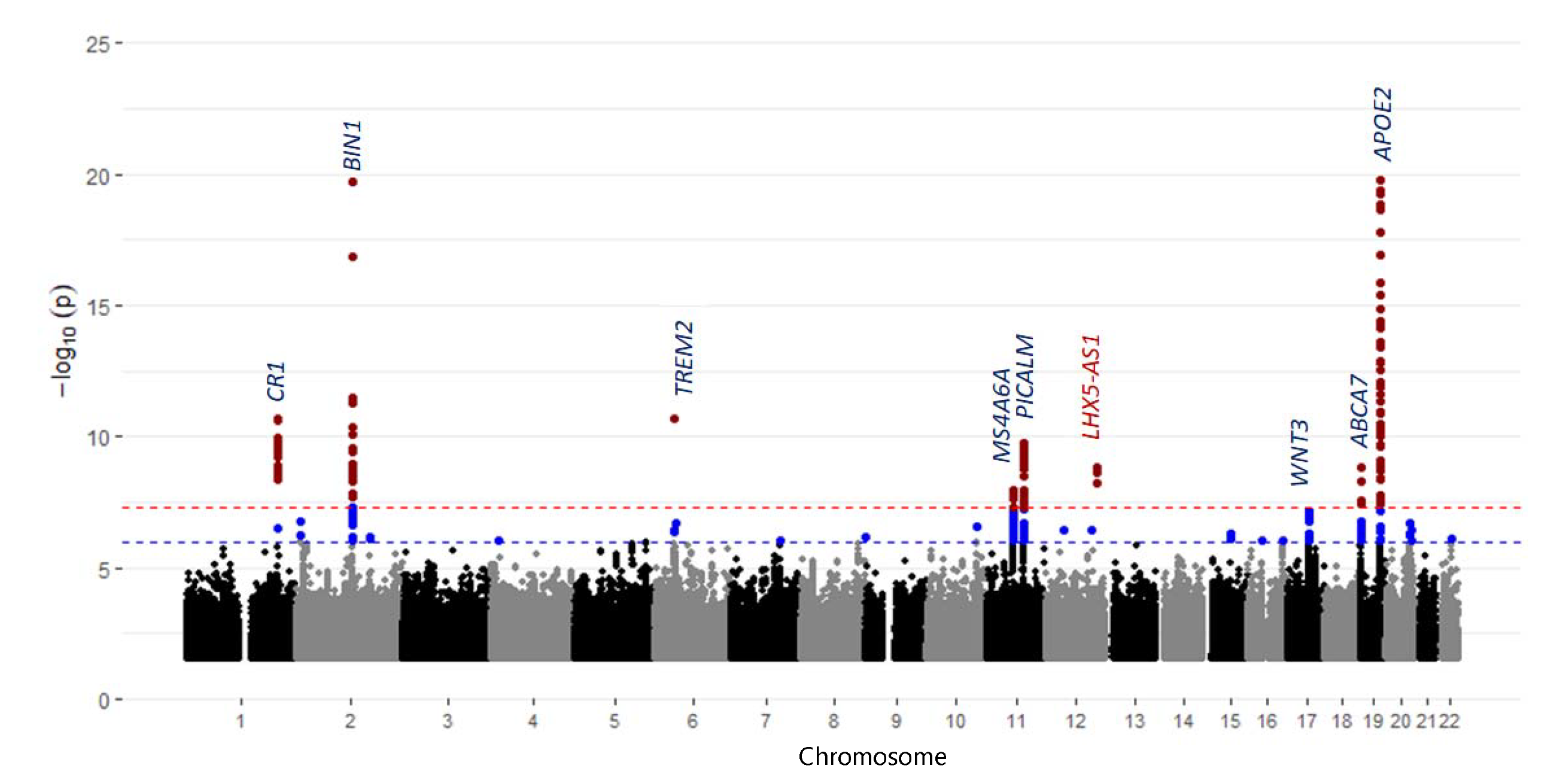
Manhattan plot of cross-ancestry meta-analyses of genome-wide associations estimated with covariate adjustment for age-at-onset (cases)/age-at-last-exam (controls), sex, dosage of the *APOE* ε4 allele, and PCs for population substructure.

**Table 1.** Summary statistics for identified loci reaching genome-wide statistical significance.in cross-ancestry meta-analyses of genome-wide associations estimated with covariate adjustment for age-at-onset (cases)/age-at-last-exam (controls), sex, and PCs for population substructure.

| Variant | Chr | Basepair | Nearest Gene | Ref/<br>Eff Alleles | Cross-Ancestry |  | AFA |  |  | EAS |  |  | HIS |  |  | NHW |  |  |
| --- | --- | --- | --- | --- | --- | --- | --- | --- | --- | --- | --- | --- | --- | --- | --- | --- | --- | --- |
|  |  |  |  |  | OR (95% CI) | P | EAF | OR (95% CI) | P | EAF | OR (95% CI) | P | EAF | OR (95% CI) | P | EAF | OR (95% CI) | P |
| Previous genome-wide-significant loci still reaching significance |  |  |  |  |  |  |  |  |  |  |  |  |  |  |  |  |  |  |
| rs679515 | 1 | 207577223 | CR1 | T/C | 1.14<br>(1.10-1.19) | 5.76E-12 | 0.043 | 1.11<br>(0.89-1.38) | 0.356 | 0.059 | 1.66<br>(0.70-3.93) | 0.245 | 0.121 | 1.07<br>(0.96-1.2) | 0.232 | 0.203 | 1.16<br>(1.11-1.20) | 6.92E-12 |
| rs6733839 | 2 | 127135234 | BIN1 | C/T | 1.12<br>(1.05-1.2) | 7.18E-26 | 0.398 | 1.13<br>(1.04-1.23) | 0.00315 | 0.348 | 0.99<br>(0.83-1.18) | 0.940 | 0.407 | 1.08<br>(1.00-1.16) | 0.0422 | 0.402 | 1.20<br>(1.16-1.24) | 4.86E-25 |
| rs75932628 | 6 | 41161514 | TREM2 | C/T | 1.44<br>(0.47-4.4) | 1.66E-12 | 0.001 | 0.66<br>(0.18-2.43) | 0.532 | NA | NA | NA | NA | NA | NA | 0.007 | 2.18<br>(1.77-2.69) | 2.39E-13 |
| rs1385742 | 6 | 47627419 | CD2AP | A/T | 1.10<br>(1.06-1.14) | 7.59E-10 | 0.422 | 1.07<br>(0.98-1.16) | 0.133 | 0.339 | 1.80<br>(1.02-3.18) | 0.0433 | 0.363 | 1.10<br>(1.02-1.19) | 0.0175 | 0.369 | 1.10<br>(1.06-1.14) | 3.75E-8 |
| rs2741342 | 8 | 27472579 | PTK2B | C/T | 0.92<br>(0.86-0.98) | 1.22E-8 | 0.319 | 0.97<br>(0.89-1.05) | 0.423 | 0.387 | 0.79<br>(0.66-0.95) | 0.0109 | 0.22 | 0.98<br>(0.90-1.07) | 0.634 | 0.225 | 0.89<br>(0.86-0.93) | 8.18E-9 |
| rs1532278 | 8 | 27608798 | CLU | T/C | 0.92<br>(0.89-0.95) | 3.70E-8 | 0.199 | 0.92<br>(0.83-1.02) | 0.0979 | 0.276 | 1.00<br>(0.84-1.20) | 0.977 | 0.29 | 0.88<br>(0.81-0.96) | 0.00220 | 0.382 | 0.92<br>(0.89-0.96) | 5.18E-6 |
| rs34173062 | 8 | 144103704 | SHARPIN | G/A | 1.20<br>(1.04-1.39) | 1.81E-9 | 0.019 | 0.91<br>(0.66-1.28) | 0.601 | NA | NA | NA | 0.057 | 1.37<br>(1.16-1.61) | 1.88E-4 | 0.086 | 1.19<br>(1.12-1.27) | 1.38E-7 |
| rs1582763 | 11 | 60254475 | MS4A6A | G/A | 0.92<br>(0.86-0.98) | 6.76E-14 | 0.104 | 1.04<br>(0.91-1.19) | 0.603 | 0.175 | 0.96<br>(0.83-1.12) | 0.602 | 0.306 | 0.90<br>(0.83-0.98) | 0.0142 | 0.366 | 0.88<br>(0.85-0.91) | 3.75E-14 |
| rs1898895 | 11 | 86126627 | PICALM | T/C | 0.89<br>(0.86-0.91) | 1.11E-14 | 0.13 | 0.89<br>(0.78-1.00) | 0.0515 | 0.391 | 0.87<br>(0.77-0.97) | 0.0128 | 0.258 | 0.92<br>(0.85-1.01) | 0.0696 | 0.313 | 0.88<br>(0.85-0.91) | 2.97E-12 |
| rs12151021 | 19 | 1050875 | ABCA7 | A/G | 1.10<br>(1.06-1.13) | 1.59E-8 | 0.404 | 1.04<br>(0.95-1.13) | 0.382 | 0.434 | 1.17<br>(0.74-1.84) | 0.505 | 0.327 | 1.10<br>(1.02-1.19) | 0.0177 | 0.338 | 1.11<br>(1.07-1.15) | 1.33E-7 |
| rs429358 | 19 | 44908684 | APOE ε4 | C/T | 3.10<br>(2.75-3.49) | <1E-320 | 0.241 | 2.66<br>(2.41-2.94) | 1.31E-85 | 0.167 | 4.94<br>(4.56-5.36) | 1.81E-85 | 0.203 | 2.26<br>(2.06-2.47) | 1.49E-71 | 0.259 | 3.20<br>(3.08-3.33) | 8.9E-734 |
| rs7412 | 19 | 44908822 | APOE ε2 | C/T | 0.54<br>(0.50-0.60) | 3.79E-106 | 0.104 | 0.59<br>(0.52-0.68) | 5.65E-14 | 0.04 | 0.44<br>(0.37-0.52) | 8.89E-7 | 0.056 | 0.67<br>(0.57-0.78) | 7.86E-7 | 0.058 | 0.48<br>(0.45-0.52) | 4.35E-86 |
| Novel genome-wide-significant loci |  |  |  |  |  |  |  |  |  |  |  |  |  |  |  |  |  |  |
| rs12576934 | 11 | 41714561 | LRRC4C | C/A | 1.12<br>(1.08-1.16) | 5.37E-9 | 0.127 | 1.09<br>(0.96-1.23) | 0.194 | 0.335 | 1.14<br>(0.99-1.30) | 0.0597 | 0.206 | 1.13<br>(1.03-1.24) | 0.00761 | 0.168 | 1.11<br>(1.06-1.16) | 1.81E-6 |
| rs111486601 | 12 | 113643122 | LHX5-AS1 | C/T | 0.63<br>(0.32-0.81) | 1.06E-8 | 0.018 | 1.00<br>(0.74-1.35) | 0.992 | NA | NA | NA | 0.039 | 0.44<br>(0.34-0.57) | 4.77E-10 | 0.001 | 0.50<br>(0.16-1.60) | 0.245 |

In addition to these two novel loci, 11 known AD loci were genome-wide significant in cross-ancestry meta-analysis (**Table 1**). Among these, the association signals from *BIN1*, *CD2AP, PTK2B, CLU, SHARPIN, MS4A6A, PICALM, ABCA7*, and *APOE* were supported by contributions from at least two ancestry groups with *P*<0.05 (**Supplementary Figures 7-15**).

In the *APOE* □4-unadjusted model, of the genome-wide significant SNPs at the 13 loci, two (15%) were GWS and seven (54%) were nominally significant (*P*<0.05) in HIS; one (7.7%) was GWS and three (23%) were nominally significant in EAS; one (7.7%) each were GWS and nominally significant in AFA; and seven (54%) were at least nominally significant in both NHW and HIS indicating the strongest contribution to the signals by NHW (NHW>HIS>EAS>AFA) (**Figure 2**, **Supplementary Figures 7-15**). While sample sizes differed between ancestry groups, marker allele frequencies also varied widely, though directions of association were largely consistent across ancestry groups for most reported loci.

Following up other previously-reported loci reaching genome-wide significance in cross-ancestry analyses (**Supplementary Table 8**), we observe strong associations near but not attaining GWS at several loci including *SORL1* (rs117618017, *P*=9.6×10^−8^), *ECHDC3* (rs7920721, *P*=1.1×10^−7^), and *ABCA7* (rs115882880, *P*=1.0×10^−7^). Within ancestries, the *PALM2AKAP2*, previously observed in a GWAS of Japanese subjects that is a subset of our EAS sample, was observed only among EAS participants (rs913360, *P*=2.7×10^−8^), and the noted association at *ABCA7* was driven almost exclusively by association in AFA individuals (rs117618017, *P*=9.6×10^−8^) which included subjects from the dataset that originally identified *ABCA7* in AFA. Notably, a variant in the *APOE* region of chromosome 19 near *APOC1*, rs157591, demonstrated GWS associations in AFA (*P*=3.3×10^−17^) and HIS participants (*P*=1.7×10^−20^), but only nominal significance in NHW (*P*=6.3×10^−3^); further adjustment for *APOE* □4 modestly reduced strength of association in AFA (*P*=4.0×10^−14^), with no association in HIS (*P*=0.573) or NHW (*P*=0.237), suggesting an ancestry-specific association independent of *APOE* □4 in AFA.

#### *APOE* □*4-Adjusted Model*

In cross-ancestry meta-analysis adjusting for *APOE* □4, (**Supplementary Figure 3** and **Table 2)**, rs111486601 near *LHX5-AS1* remained genome-wide significant (*P*=1.47×10^-9^) while rs12576934 near *LRRC4C* showed reduced genome-wide significance (*P*=7.13×10^-6^ (**Table 2**). Besides the *APOE* ε2 SNP, six known AD loci, including *CR1*, *BIN1*, *TREM2*, *MS4A6A*, *PICALM*, and *ABCA7*, remained genome-wide significant when adjusting for the *APOE* □4 dosage. In addition, in this model a locus within *WNT3* (rs430685; *P*=7.4×10^-8^) near *MAPT* emerged with near genome-wide significance (**Table 2**).

**Table 2.** Summary statistics for identified loci reaching genome-wide statistical significance in cross-ancestry meta-analyses of genome-wide associations estimated with covariate adjustment for age-at-onset (cases)/age-at-last-exam (controls), sex, dosage of *APOE* ε4 alleles, and PCs for population substructure.

| Variant | Chr | Basepair | Nearest Gene | Ref/<br>Eff Alleles | Cross-Ancestry |  | AFA |  |  | EAS |  |  | HIS |  |  | NHW |  |  |
| --- | --- | --- | --- | --- | --- | --- | --- | --- | --- | --- | --- | --- | --- | --- | --- | --- | --- | --- |
|  |  |  |  |  | OR (95% CI) | P | EAF | OR (95% CI) | P | EAF | OR (95% CI) | P | EAF | OR (95% CI) | P | EAF | OR (95% CI) | P |
| Previous genome-wide-significant loci still reaching significance |  |  |  |  |  |  |  |  |  |  |  |  |  |  |  |  |  |  |
| rs679515 | 1 | 207577223 | CR1 | T/C | 1.14<br>(1.04-1.24) | 2.24E-11 | 0.042 | 1.15<br>(0.91-1.46) | 0.235 | 0.059 | 1.56<br>(0.64-3.82) | 0.327 | 0.122 | 1.03<br>(0.92-1.16) | 0.572 | 0.203 | 1.18<br>(1.13-1.24) | 4.54E-12 |
| rs6733839 | 2 | 127135234 | BIN1 | C/T | 1.15<br>(1.10-1.21) | 1.84E-20 | 0.398 | 1.16<br>(1.06-1.27) | 9.15E-4 | 0.347 | 1.01<br>(0.84-1.22) | 0.890 | 0.407 | 1.11<br>(1.03-1.20) | 0.00791 | 0.402 | 1.19<br>(1.14-1.24) | 6.20E-18 |
| rs75932628 | 6 | 41161514 | TREM2 | C/T | 1.10<br>(0.21-5.66) | 2.08E-11 | 0.001 | 0.41<br>(0.11-1.56) | 0.193 | -- | -- | -- | -- | -- | -- | 0.007 | 2.25<br>(1.80-2.81) | 1.36E-12 |
| rs10792263 | 11 | 60312178 | MS4A6A | C/T | 1.09<br>(1.03-1.14) | 1.12E-8 | 0.622 | 1.02<br>(0.94-1.12) | 0.587 | 0.716 | 1<br>(0.88-1.14) | 0.998 | 0.59 | 1.13<br>(1.04-1.22) | 0.00235 | 0.627 | 1.12<br>(1.07-1.16) | 6.44E-8 |
| rs543293 | 11 | 86109035 | PICALM | A/G | 0.89<br>(0.86-0.92) | 1.88E-10 | 0.136 | 0.92<br>(0.81-1.04) | 0.181 | 0.392 | 0.88<br>(0.78-0.99) | 0.0326 | 0.263 | 0.91<br>(0.84-1.00) | 0.0429 | 0.32 | 0.89<br>(0.85-0.93) | 1.78E-8 |
| rs430685 | 17 | 46781782 | WNT3 | T/C | 0.89<br>(0.85-0.93) | 7.42E-8 | 0.137 | 0.92<br>(0.81-1.05) | 0.229 | -- | -- | -- | 0.185 | 0.87<br>(0.79-0.97) | 0.00902 | 0.223 | 0.89<br>(0.85-0.94) | 3.14E-6 |
| rs115882880 | 19 | 1001778 | GRIN3B | C/A | 1.23<br>(0.94-1.59) | 1.45E-9 | 0.101 | 1.50<br>(1.32-1.70) | 2.45E-10 | -- | -- | -- | 0.042 | 1.21<br>(1.00-1.47) | 0.0556 | 0.003 | 0.82<br>(0.53-1.28) | 0.392 |
| rs7412 | 19 | 44908822 | APOE ε2 | C/T | 0.70<br>(0.62-0.79) | 2.00E-30 | 0.103 | 0.73<br>(0.63-0.85) | 4.59E-5 | 0.04 | 0.79<br>(0.49-1.27) | 0.331 | 0.056 | 0.77<br>(0.65-0.91) | 0.00251 | 0.059 | 0.63<br>(0.58-0.69) | 3.67E-26 |
| Novel genome-wide-significant loci |  |  |  |  |  |  |  |  |  |  |  |  |  |  |  |  |  |  |
| rs111486601 | 12 | 113643122 | LHX5-AS1 | C/T | 0.57<br>(0.28-1.17) | 1.47E-9 | 0.017 | 0.96<br>(0.71-1.28) | 0.767 | NA | NA (NA) | NA | 0.037 | 0.40<br>(0.30-0.52) | 9.13E-11 | 0.001 | 0.42<br>(0.14-1.27) | 0.124 |

### Novel ancestry-specific loci

In the *APOE* ε4-unadjusted model, ancestry-specific meta-analysis identified one novel genome-wide significant SNP for AD in NHW near *KIAA0825* (rs141408991; OR=0.50, CI=0.39-0.64, *P*=2.9×10^−8^) (**Table 3**). Under the *APOE* ε4-adjusted model, ancestry-specific meta-analyses revealed two novel loci in HIS, one near *GRB14* (rs73978419; OR=0.0.44, 95% CI=0.33-0.59, *P*=1.3×10^−8^) and one near *PTPRK* (rs67714619; OR=1.34, 95% CI=1.21-1.48, *P*=2.4×10^−8^) (**Table 3**).

**Table 3.**
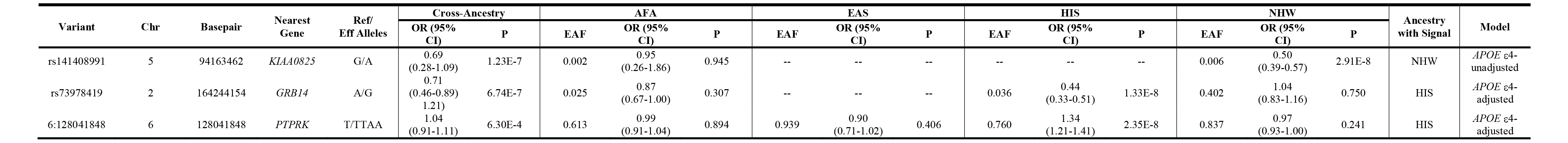
Summary statistics for identified loci reaching genome-wide statistical significance in within-ancestry meta-analyses estimated under minimum covariate adjustment (age-at-onset (cases)/age-at-last-exam (controls), sex, and PCs for population substructure) and extended covariate adjustment models (age-at-onset (cases)/age-at-last-exam (controls), sex, dosage of *APOE* ε4 alleles, and PCs for population substructure) for identified loci reaching genome-wide significance.

### Functional Annotation of Significantly-Associated Multi-Ancestry Loci

The lead variant of the *LRRC4C* locus (rs12576934) lies in the intergenic region 254,788 basepairs upstream from the *LRRC4C* (closest protein-coding gene; negative strand) transcription start site (TSS) and overlaps enhancers in cultured neurons suggesting a potential role in enhancer regulation in tissues/cell types related to AD pathogenesis (**Supplementary Figure 16**). Notably, rs12576934 is a significant splicing quantitative trait locus (sQTL) for *LRRC4C* in brain frontal cortex (*P*=5.89×10^-4^) from the Genotype-Tissue Expression (GTEx) Consortium^16^. However, rs12576934 does not show a significant association with *LRRC4C* expression in any brain-tissue related expression QTL (eQTL) data, including GTEx, Metabrain^17^, the Microglia Genome Atlas (MiGA)^18^, and Accelerating Medicine Partnerships-Alzheimer’s Disease (AMP-AD)^19^ resources.

For the *LHX5-AS1* locus, the lead variant (rs111486601) is located 170,842 base pairs upstream of the *LHX5* TSS (closest protein-coding gene; negative strand). Multiple variants in linkage disequilibrium (LD; estimated using 1000 Genomes NHW populations^20-22^) with rs111486601 overlap ROADMAP^23^ ChromHMM^24^ brain enhancers in neuronal culture cells, hippocampus middle region dorsolateral prefrontal cortex, as well as blood enhancers in primary B and NK cells, suggesting potential regulatory roles in immune cell types and brain regions that are known to be related to AD. The variant rs111486601 overlaps and potentially disrupts multiple transcription factor binding sites (TFBS) as predicted by Hypergeometric Optimization of Motif Enrichment (HOMER^25^). Given that rs111486601 is rare in NHW (MAF=0.0006), it is not presently observed in any QTL database (GTEx, Metabrain, MiGA, or AMP-AD), as these databases have been generated using common European ancestry variants.

### Differential Expression Analysis of Significantly Associated Loci

Of eight genes flanking intergenic SNPs or containing GWS SNPs from the five novel loci, *LRRC4C* and *KIAA0825* were differentially expressed between autopsy-confirmed AD brains and brains of cognitively-intact decedents with *P*<0.05 in secondary analysis of previously-reported^26^ differential expression analyses using autopsied Framingham Heart Study (FHS, **Supplementary Table 9**) brains (strongest association at *LRRC4C*: log_2_FC=-0.01, *P*=8.8×10^-4^), while only expression of *LRRC4C* was significantly associated with Braak stage in FHS in autopsied AD brains (best association at *LRRC4C*: β=-0.11, *P*=4.7×10^-3^).

### Pathway analyses

We performed pathway enrichment analyses including genes containing or flanking suggestively associated SNPs (*P*<10^-6^) from the cross-ancestry meta-analysis. We selected 86 and 60 genes for *APOE* □4-unadjusted and -adjusted models, respectively. For both models, amyloid-beta related pathways were the top-ranked pathway, indicating that the relevance of this pathway in cross-ancestry meta-analysis was unaffected by *APOE* □4 adjustment. In the *APOE* □4-unadjusted model, lipid transport, endocytosis, and classical complement pathways were in addition significantly associated (*P*_adjusted_<0.05) (**Supplementary Table 10a**). Examining the *APOE* □4-adjusted model, classical complement and phospholipid efflux pathways were now more significantly associated and ranked higher than chylomicron remnant clearance, cholesterol, and endocytosis pathways with lower significance (**Supplementary Table 10b**).

Employing the ancestry-specific GWAS results from the two models, we determined pathways shared across ancestry groups (**Supplementary Table 11**). Among the significant pathways in each ancestry (*P*_adjusted_<0.05) from the *APOE* □4-unadjusted model, seven pathways were shared across all four ancestries related to lipid metabolism pathways (**Supplementary Figure 17a**). In the *APOE* □4-adjusted model no pathways with *P*_adjusted_<0.05 were shared across ancestries (**Supplementary Figure 17b**).

## DISCUSSION

In the largest cross-ancestry GWAS meta-analysis in AD to date, we identified two novel genome-wide cross-ancestry associations near *LRRC4C* and *LHX5-AS1*, and three ancestry-specific genome-wide significant loci near *KIAA0825* (NHW), and *GRB14* and *PTPRK* (HIS). Functional annotation follow-up revealed the *LRRC4C* variant’s potential regulatory role in enhancer regulation in tissues and cell types related to AD pathogenesis, and the involvement of the *LHX5-AS1* variant in regulatory roles in immune cell types and brain regions. *In silico* differential expression analyses across brain tissues suggested differential expressions of *LRRC4C* and *KIAA0825* between AD and control brains and association of *LRRC4C* expression with Braak stage. Pathway analyses implicated pathways associated with amyloid metabolism, cholesterol transport, and inflammation, consistent with previous pathway-based findings in AD.

While the available AD GWAS data from non-European ancestry groups continue to be dwarfed by the sample sizes of AD GWAS in European ancestry subjects, our analyses demonstrate the power of multi-ancestry datasets. Of particular interest is the significant association at the *SHARPIN* locus in our study. The *SHARPIN* locus was first observed in a study of 111,326 clinically-diagnosed and ‘proxy’ AD cases and 677,663 controls individuals of European ancestry (rs34173062, MAF=0.081, OR=1.13, *P*=1.7×10^-6^) and later confirmed in much larger sample sizes^4,27,28^. While NHW participants in this study had similar MAF (0.086 here vs. 0.081) and similar effect size (OR=1.19 here vs. 1.13), the much larger effect size in HIS participants (OR_HIS_=1.37; MAF_HIS_=0.057) ensured sufficient power to detect a GWS cross-ancestry association in just 56,241 subjects, only 7.1% the sample size of Bellenguez et al., an astonishing reflection on the unique power leveraging diverse ancestries can provide.

Among the novel loci identified in cross-ancestry analyses, both *LRRC4C* and *LHX5-AS1* are potential biological candidates for roles in AD. *LRRC4C* encodes leucin-rich repeat (LRR) containing adhesion molecules, which are key organizers of inhibitory and excitatory synapses. Mutations within the gene have been implicated in neurodevelopmental disorders, such as autism^29^ and intellectual disability^30^. *LRRC4C* variants with suggestive associations with CSF b-site APP cleaving enzyme (BACE) have been previously discovered in ADNI^31^. Additionally, one study used *LRRC4C* knock-out mice in an experimental multiple sclerosis model and found a neuron-protective role for *LRRC4C*, concluding that the ectopic expression of *LRRC4C* protected neurons from immune damage^32^. Associated variants in the *LRRC4C* locus are significantly associated with sQTL for *LRRC4C* in GTEx brain frontal cortex, and fall in a regulatory region that are notably enhancers in neuron tissue^33^, with transcription factor (TF) Chromatin ImmunoPrecipitation Sequencing (ChIP-Seq) experiments showing the region overlapping our index variant as being a target of Srebf1 in exocrine gland, epithelium and mammary gland tissues. Srebf1 is a transcriptional activator required for lipid homeostasis and has been identified as a proneural transcription factor in radial glia^34^. Variants of *SREBF1* influence AD risk by moderating the deleterious effect of the *APOE* ε4 allele^35^.

*LHX5-AS1* is an antisense RNA gene, which is complementary to the mRNA *LHX5* with which it hybridizes and blocks its translation into protein. *LHX5* encodes LIM homeobox 5, which is essential for the regulation of precursor cell proliferation. This gene controls neuronal differentiation and migration during hippocampal development. *LHX5* and *LHX5-AS1* are similarly expressed in various regions of the adult central nervous system including hypothalamus, spinal cord, cerebellum, basal ganglia, and cerebellar hemisphere. Significantly associated variants for this locus fall in a regulatory region, for example, bivalent enhancers in hematopoietic multipotent progenitor cells and brain^33^. Our specific index variant is an enhancer in blood and a transcription variant in neural progenitor cells^33^. Variants in 12q24 have previously been associated with hippocampal volume^36^. This novel locus is ∼360 kb away from the recently-reported locus at *TPCN1*^4^.

Notably, most of the novel loci identified in ancestry-specific analyses act in biologically plausible pathways. The protein encoded by *Grb14* functions as a negative modulator of insulin receptor activity (IR) activity. Both insulin resistance and type 2 diabetes (T2D) and have been implicated in AD by a large body of epidemiological, neuropathological, and experimental studies^37^. Brain tissue from individuals with AD show major abnormalities in insulin signaling and increased presence of disease-specific pathological lesions, neurodegeneration, and neuronal vulnerability^37,38^. In structural and functional neuroimaging studies, T2D and insulin resistance are associated with white increased burden of white matter lesions, decreased hippocampal volume, regional cerebral blood flow and oxygenation^39-43^. *PTPRK* encodes a member of the protein tyrosine phosphatase (PTP) family regulating a variety of cellular processes including cell growth, differentiation, mitotic cycle, and oncogenic transformation. Variants in *PTPRK* were recently found to be associated with reaction time in a GWAS on cognitive function in 300,486 individuals of European ancestry^44^. *KIAA0825* encodes a protein of largely unknown function; however, it is differentially expressed in AD vs. control in brain RNAseq data (see URL below).

Leveraging GWAS on diverse ancestries, here we observed evidence for novel loci not seen in prior multi-ancestry or large NHW-only studies. Especially salient is the association at the *SHARPIN* locus, first observed as a dementia locus in a single-ancestry NHW GWAS almost 13 times larger than the current study, and observed here with far fewer samples (rs34173062, *P*=1.81×10^-9^).

While this study yielded several novel AD susceptibility loci, the fewer non-European ancestry samples relative to European ancestry samples (57.2% of participants) remains a limitation. This constrains the identification of rare variants and novel ancestry-specific loci in highly admixed groups including AFA and HIS, where heterogeneity in levels of European, African, and Amerindian admixture may strongly limit statistical power. Future work planned for these data includes global and local admixture analyses to provide detailed characterization of patterns of admixture and adjust for these positional and ancestry-specific background differences. While more non-European ancestry AD GWAS are valuable for cross-ancestry comparisons of AD genetic risk profiles, even modest amounts of diverse data tremendously boost our ability to detect novel AD susceptibility loci.

## METHODS

### Variant-and sample-level quality control (QC)

We performed QC on individual ADGC datasets using a multi-step pipeline with key functions implemented in PLINK v1.9^45-47^. All datasets included in analysis included only cases that met either NINCDS-ADRDA^48^ or NIA-AA^49^ criteria for clinical diagnosis of AD and had an age-at-onset >60 years or had autopsy-confirmation of late-onset AD pathology; controls/non-cases (“cognitively-normal elders”, CNEs) were individuals with a mini-mental status examination (MMSE)>26 or modified mini-mental status examination (3MS) score>87 at most recent exam and/or were reported to be cognitively intact at time of death (details in **Supplementary Note 1**). We identified and excluded low-quality variants and samples after re-estimating all quality metrics following an initial filter excluding variants with a genotype missingness rate of >10%. Variant-level QC filters implemented thereafter included exclusions of (a) SNPs with call rates below 98% for Illumina and 95% for Affymetrix panels; (b) SNPs with departure from Hardy-Weinberg Equilibrium (HWE) of *P*<10^-6^ among controls for variants of MAF>0.01; and (c) SNPs with informative missingness by case-CNE status of *P*<10^-6^. For sample-level QC, samples were excluded if (a) the individual genotyping call rate was <95%; (b) if X chromosome heterozygosity indicated inconsistency between predicted and reported sex; or (c) if population substructure analyses (described below) indicated the sample did not cluster with samples in its reported ancestry grouping when compared against multi-ancestry data from the 1000 Genomes (1kG) Phase 3 reference panel. For examples, reported NHW samples were expected to cluster with 1kG European ancestry sample groups while reported AFA samples being admixed were expected to cluster between and around 1kG European and African ancestry sample groups.

### Relatedness Checks

Relatedness was assessed using the “--genome” function of PLINK v1.9. Using a common set of ∼20,000 linkage disequilibrium (LD)-pruned SNPs sampled from among genotyped variants across ancestry groups, 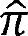 (the proportion of alleles shared IBD) was estimated across all pairs of participants across all ADGC datasets. Among pairs of participants with no known familial relationships, one sample was excluded among pairs with 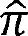>0.95 if phenotype and covariate data were identical, otherwise both samples were excluded; among all pairs with 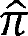>0.4 but less than 0.95, one sample was kept giving preference to cases over CNEs, age (earlier age-at-onset among case pairs, later age-at-exam among CNE pairs). Pairs of relatives were dropped from family datasets if 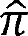 differed substantially from expectation based on their reported relationships.

### Population substructure

To identify outlier samples within each dataset with ancestry group, we performed a principal components analysis (PCA) using ‘smartpca’ in EIGENSOFT^50,51^ on the subset of ∼20,000 LD-pruned SNPs used for relatedness checks on genotypes from all samples within each individual dataset and from the 1kG Phase 3 reference panels. Individuals not clustering with their reported ancestry groups (or between reported ancestry groups for admixed subjects) were excluded from analysis when including 1kG groups. To account for the effects of population substructure in our analysis, a second PCA was performed using only the remaining individuals in each dataset. Principal components (PCs) 1-10 were examined for association with AD case-control status and eigenvector loading, and only PCs showing nominal association with AD (*P*<0.05) and eigenvector loadings >3 were used in covariate adjustment for populations substructure (average number of PCs used is 3; range: 2-4).

### TOPMed Imputation

For each dataset, SNPs not directly genotyped were imputed on the Trans-Omics for Precision Medicine (TOPMed) Imputation Server (TIS)^52^ using samples of all ancestries available on the release 5 (R5) reference panel^53^, which includes 308,107,085 SNPs observed on 194,512 haplotypes (from 97,256 participants), all with an estimated minor allele count (MAC) ≥5 and observed in samples from at least two separately-ascertained data sources. Phasing on the TIS was done with EAGLE^54^, while imputation was performed using Minimac4^55^. Quality of imputation for all variants was assessed using *R^2^* for imputation quality, although all variants were retained and not filtered prior to analysis. Following imputation and analysis, SNPs of all frequency were filtered using a conservative quality threshold, *R^2^* ≥ 0.8, to assure high quality of rare variants with MAF≤0.01. Analysis of SNP imputation quality by bin of MAF revealed that more than 80% of variants of MAF>0.0005 had *R^2^*≥0.8; among variants with MAF≤0.0005, approximately 50% of variants had *R^2^* ≥ 0.8 in most datasets with n>1,000 participants. After association analyses at several cross-dataset and within-dataset filtering thresholds, reported analyses include only variants of *R^2^* ≥ 0.8 within each dataset for the purpose of noise reduction. Genotyped and imputed variants were all mapped to the GRCh138/hg38 human genome build.

### Single-variant Association Analysis and Within-Ancestry Meta-analysis for Common Variants (MAF>0.01)

Single variant-based association analysis on datasets of unrelated cases and CNEs were performed in SNPTEST^56^ using score-based logistic regression under an additive model with covariate adjustment for PCs, age (defined as age-at-onset for cases and age-at-last exam for CNEs), sex, and in certain datasets, additional adjustment for study-specific indicator variables (“*APOE*-unadjusted”; Model 1). A second model (“*APOE*-adjusted”; Model 2) included covariate adjustment for dosage of *APOE* ε4 (0/1/2 copies), which is commonly observed as an effect modifier. Family-based datasets were analyzed using GMMAT, an R package for performing association tests using generalized linear mixed models (GLMMs). These GLMMs allow for both a binary outcome and adjustment for relatedness via a genetic relationship matrix (GRM). Score tests were performed for each genetic variant assuming an additive model with fixed effect covariate adjustment for PCs, age (defined as age-at-onset for cases and age-at-last exam for CNEs), sex and random effect adjustment for the GRM (calculated using GEMMA) in the family-based datasets. After association analysis on imputed data, variants with regression coefficient of |β|>5 and any erroneous estimates (negative standard errors or *P*-values equal to 0 or 1) were excluded from further analysis. Within-study association results for variants common (MAF>0.01) to at least one study were meta-analyzed using a fixed-effects approach with inverse variance-weighting in METAL^57^.

### Single-variant Association Analysis and Within-Ancestry Meta-analysis for Rare Variants (MAF≤0.01)

Rare variant association and meta-analysis was performed for individual variants using the SeqMeta^58^ package in R^59^. SeqMeta performs a score-based logistic regression, estimating scores in individuals using ‘prepScores()’ and performing meta-analysis using ‘singleSNPMeta()’. Family-based datasets were analyzed using GMMAT as described previously for common variants and no datasets with fewer than 100 cases and/or CNEs were analyzed. As in common variant analyses, models evaluated included covariate adjustment for PCs. As for common variants, any SNPs with a regression coefficient of |β|>5 and any erroneous estimates were excluded from further analysis after meta-analysis of imputed rare variants.

### Single-variant Cross-Ancestry Meta-Analysis

All cross-ancestry meta-analyses were performed using the Han-Eskin random-effects (RE_HE_) model as implemented in METASOFT^60^, which is optimized for the detection of cross-ancestry associations in the presence of effect heterogeneity between ancestry groups. The RE_HE_ model has similar power to a fixed-effects model when effect heterogeneity between ancestry groups is modest. Cross-ancestry meta-analyses incorporated within-ancestry genome-wide summary statistics from the single variant fixed-effects meta-analyses described above for NHW, AFA, HIS, and EAS ancestry groups (SNPTEST/GMMAT METAL for common variants [MAF≥0.01]; SeqMeta/GMMAT for rare variants [MAF<0.01]). In the primary cross-ancestry analyses, all variants with a within-ancestry MAF>0.01 in at least one dataset were incorporated into analyses.

### Functional Annotation of Significantly Associated Loci

We used all annotations in Functional genomics repository (FILER)^61^ for our analyses. To reveal potential biological functions of the variants, experimental studies have shown that matching tissues/cell types to the phenotype of interest is one of the keys to success. Thus, we focused our analyses on the brain-related tracks harmonized in FILER. These brain-related tracks originated from ROADMAP^23^ (for enhancer and active histone marks), Encyclopedia of DNA Elements (ENCODE^62^; active histone marks) and HOMER^25^ (predicted TFBSs). We annotated the five-novel genome-wide significant signals (cross ancestry and ancestry-specific results) using brain-related tracks harmonized in FILER. By annotating variants with these various functional genomics data, we identified variants with converging functional evidence and determined which variants had a higher chance of being functional in the brain.

### Differential Expression Analysis of Significantly Associated Loci

We followed up SNPS with genome-wide significant associations (GWS; *P*<5×10^-8^). If the GWS SNP is intergenic, we selected closest genes of the SNP. If the GWS SNP resides in the gene, we selected the gene. The selected genes were assessed for differential expression between AD and control brains from prefrontal cortex tissue specimens of 208 participants (64 autopsy-confirmed AD cases and 129 controls) of the Framingham Heart Study (FHS) and Boston University Alzheimer’s Disease Center (BUADC). Details of diagnosis, data cleaning, and analysis methods were previously reported^26,63^. In brief, differential gene expression between AD and control brains was performed using the LIMMA^64,65^ software. Expression of a gene was compared in AD and control brains using linear regression models including the log_2_-transformed normalized expression values and terms for age at death (AAD), sex, and RNA integrity number (RIN). We also evaluated associations of gene expression for Braak staging for neurofibrillary tangles and the Consortium to Establish a Registry for Alzheimer Disease (CERAD) semi-quantitative criteria for neuritic plaques (CERAD Score)^66^. Values for each trait were adjusted for AAD and sex, and the residuals were rank-transformed as previously described^67^. Association of log_2_-transformed expression levels for each rank-transformed neuropathological trait was evaluated using linear regression models that adjusted for RIN in the FHS/BUADC dataset.

### Pathway Analysis

For pathway analysis, we selected flanking genes of suggestive SNPs with *P*<10^-6^ from cross-ancestry meta-analysis and each within-ancestry meta-analysis. We conducted pathway analysis using GO Biological Process in the EnrichR program^68^. For cross-ancestry signals, we compared significantly enriched pathways after FDR corrected *P*-value (*P*_adjusted_<0.05) between *APOE*-unadjusted and -adjusted models. For each analysis model, we created Venn diagrams of pathways with *P*_adjusted_<0.05 from each ancestry. We reported both unique and shared pathways across different ancestry populations.

## Data Availability

Data Availability: Data are available through the National Institute on Aging Genetics of Alzheimer Disease Data Storage Site (NIAGADS) Data Sharing Service (DSS)

https://dss.niagads.org/datasets/ng00067/.

## ACKNOWLEDGEMENTS

The National Institutes of Health, National Institute on Aging (NIH-NIA) supported this work through the following grants: ADGC, U01 AG032984, RC2 AG036528; Samples from the National Cell Repository for Alzheimer’s Disease (NCRAD), which receives government support under a cooperative agreement grant (U24 AG21886) awarded by the National Institute on Aging (NIA), were used in this study. We thank contributors who collected samples used in this study, as well as patients and their families, whose help and participation made this work possible; Data for this study were prepared, archived, and distributed by the National Institute on Aging Alzheimer’s Disease Data Storage Site (NIAGADS) at the University of Pennsylvania (U24-AG041689); GCAD, U54 AG052427; NACC, U01 AG016976; NIA LOAD (Columbia University), U24AG056270; Banner Sun Health Research Institute P30 AG019610; Boston University, P30 AG013846, U01 AG10483, R01 CA129769, R01 MH080295, R01 AG017173, AG025259, R01 AG048927, RF1 AG057519, R01AG33193, R01 AG009029; Columbia University, P30AG066462, R01 AG072474, R01 AG067501; Duke University, P30 AG028377, AG05128; Emory University, AG025688; Group Health Research Institute, U01 AG006781, U01 HG004610, U01 HG006375, U01 HG008657; Indiana University, P30 AG10133, R01 AG009956, RC2 AG036650; Johns Hopkins University, P50 AG005146, R01 AG020688; Massachusetts General Hospital, P50 AG005134, P30 AG062421; Mayo Clinic, P50 AG016574, R01 AG032990, KL2 RR024151; Mount Sinai School of Medicine, P50 AG005138, P01 AG002219; New York University, P30 AG08051, UL1 RR029893, 5R01AG012101, 5R01AG022374, 5R01AG013616, 1RC2AG036502, 1R01AG035137; North Carolina A&T University, P20 MD000546, R01 AG28786-01A1; Northwestern University, P30 AG013854; Oregon Health & Science University, P30 AG008017, R01 AG026916; Rush University, P30 AG010161, R01 AG019085, R01 AG15819, R01 AG17917, R01 AG030146, R01 AG01101, RC2 AG036650, R01 AG22018; TGen, R01 NS059873; University of Alabama at Birmingham, P50 AG016582; University of Arizona, R01 AG031581; University of California, Davis, P30 AG010129; University of California, Irvine, P50 AG016573; University of California, Los Angeles, P50 AG016570; University of California, San Diego, P50 AG005131; University of California, San Francisco, P50 AG023501, P01 AG019724; University of Kentucky, P30 AG028383, AG05144; University of Michigan, P50 AG008671, P30 AG053760; University of Pennsylvania, P30 AG010124; University of Pittsburgh, P50 AG030653, P50 AG041718, P50 AG064877, P30 AG066468; University of Southern California, P50 AG005142; University of Texas Southwestern, P30 AG012300; University of Miami, R01 AG070864, AG052410, AG074527 and U01 AG058654, AG057659, AG062943, AG066767, AG076482 AND U19 AG074865; University of Washington and Kaiser Foundation Research Institute, P50 AG005136, R01 AG042437, P30 AG066509, U19 AG066567; University of Wisconsin, P50 AG033514; Vanderbilt University, R01 AG019085; and Washington University, P50 AG005681, P01 AG03991, P01 AG026276. The Kathleen Price Bryan Brain Bank at Duke University Medical Center is funded by NINDS grant # NS39764, NIMH MH60451 and by Glaxo Smith Kline. Support was also from the Alzheimer’s Association (LAF, IIRG-08-89720; MP-V, IIRG-05-14147), the US Department of Veterans Affairs Administration, Office of Research and Development, Biomedical Laboratory Research Program, and BrightFocus Foundation (M.P.-V., A2111048). P.S.G.-H. is supported by Wellcome Trust, Howard Hughes Medical Institute, and the Canadian Institute of Health Research. Genotyping of the TGEN2 cohort was supported by Kronos Science. The TGen series was also funded by NIA grant AG041232 to AJM and MJH, The Banner Alzheimer’s Foundation, The Johnnie B. Byrd Sr.

Alzheimer’s Institute, the Medical Research Council, and the state of Arizona and also includes samples from the following sites: Newcastle Brain Tissue Resource (funding via the Medical Research Council, local NHS trusts and Newcastle University), MRC London Brain Bank for Neurodegenerative Diseases (funding via the Medical Research Council),South West Dementia Brain Bank (funding via numerous sources including the Higher Education Funding Council for England (HEFCE), Alzheimer’s Research Trust (ART), BRACE, Alzheimer’s Brain Bank UK, and Development and Alumni Relations (DARO) Office, as well as North Bristol NHS Trust Research and Innovation Department and DeNDRoN), The Netherlands Brain Bank (funding via numerous sources including Stichting MS Research, Brain Net Europe, Hersenstichting Nederland Breinbrekend Werk, International Parkinson Fonds, Internationale Stiching Alzheimer Onderzoek), Institut de Neuropatologia, Servei Anatomia Patologica, Universitat de Barcelona. ADNI data collection and sharing was funded by the National Institutes of Health Grant U01 AG024904 and Department of Defense award number W81XWH-12-2-0012. Funding for Saarland University was provided by the German Federal Ministry of Education and Research (BMBF), grant number 01GS08125 to Matthias Riemenschneider. ADNI is funded by the National Institute on Aging, the National Institute of Biomedical Imaging and Bioengineering, and through generous contributions from the following: AbbVie, Alzheimer’s Association; Alzheimer’s Drug Discovery Foundation; Araclon Biotech; BioClinica, Inc.; Biogen; Bristol-Myers Squibb Company; CereSpir, Inc.; Eisai Inc.; Elan Pharmaceuticals, Inc.; Eli Lilly and Company; EuroImmun; F. Hoffmann-La Roche Ltd and its affiliated company Genentech, Inc.; Fujirebio; GE Healthcare; IXICO Ltd.; Janssen Alzheimer Immunotherapy Research & Development, LLC.; Johnson & Johnson Pharmaceutical Research & Development LLC.; Lumosity; Lundbeck; Merck & Co., Inc.; Meso Scale Diagnostics, LLC.; NeuroRx Research; Neurotrack Technologies; Novartis Pharmaceuticals Corporation; Pfizer Inc.; Piramal Imaging; Servier; Takeda Pharmaceutical Company; and Transition Therapeutics. The Canadian Institutes of Health Research is providing funds to support ADNI clinical sites in Canada. Private sector contributions are facilitated by the Foundation for the National Institutes of Health (www.fnih.org). The grantee organization is the Northern California Institute for Research and Education, and the study is coordinated by the Alzheimer’s Disease Cooperative Study at the University of California, San Diego. ADNI data are disseminated by the Laboratory for Neuro Imaging at the University of Southern California. Additional salary and analytical support were provided by NIA grants R01 AG054060 and RF1 AG061351. We thank Drs. D. Stephen Snyder and Marilyn Miller from NIA who are *ex-officio* ADGC members.

## URLs

Agora AD Knowledge Portal entry for KIAA0825 expression across brain regions: https://agora.adknowledgeportal.org/genes/ENSG00000185261/evidence/rna

## Author Contributions

*Study Design and Conception:* F.R., P.B., G.T., K.L.L., L.-S.W., R.M., L.A.F., J.L.H., M.A.P.-V., G.D.S., G.R.J., C.R., and A.C.N. *Sample Contribution:* E.A., P.M.A., A.A., M.S.A., R.L.A., M.A., L.A., L.G.A., S.E.A., S.Asthana, C.S.A., G.A., C.T.B., R.C.B., L.L.B., T.G.B., J.T.B., G.W.B., D.Beekly, B.B., D.Bennett, J.B., M.E.F., T.D.B., D.Blacker, B.F.B., J.D.B., A.Boxer, J.B.B., J.R.B., J.M.B., J.D.Buxbaum, N.J.C., L.B.C., C.Cao, C.S.C., C.M.C., R.M.C., M.M.C., S.C., M.-F.C., N.A.C., H.C.C., J.C., S.Craft, P.K.C., D.H.C., E.A.C., C.Cruchaga, M.L.C., M.C., E.D., B.D., P.L.D., C.D., J.C.D., M.Dick, D.W.D., B.A.D., R.S.Doody, R.D., N.E.-T., D.A.E., K.M.F., T.J.F., K.B.F., D.W.F., M.R.F., V.F.-H., S.F., T.M.F., M.P.F., D.R.G., A.G., M.Gearing, D.H.G., B.G., J.R.G., A.M.G., T.G., N.R.G.-R., R.C.G., J.H.G., H.H., J.H., R.L.H., O.Harari, J.Hardy, L.E.H, E.H., V.H., M.H., L.S.H., R.M.H., M.J.H., C.M.H., B.T.H., L.S.Hynan, L.I., G.P.J., S.J., L.W.J., K.J., L.J., M.I.K., A.Karydas, M.J.K., J.S.K., J.A.K., C.D.K., A.Khaleeq, R.K., J.K., N.W.K., J.H.K., W.K., F.M.L., J.J.L., E.B.L., A.L., J.B.L., A.I.L., A.P.L., R.B.L., M.Logue, O.L.L., C.G.L., D.M., D.C.M., E.R.M., F.M., D.C.Mash, E.M., P.M., A.M., W.C.M., S.M.M., A.N.M., S.M., A.C.M., M.Mesulam, J.Mez, B.L.M., C.A.M., J.W.M., T.J.M., E.S.Monuki, J.C.M., S.Mukherjee, A.J.M., T.N., S.O., J.M.O., M.O., R.P., J.E.P., H.L.P., V.P., D.P., V.Perez, E.P., R.C.P., A.P., M.P., W.W.P., H.P., L.Q., M.Q., J.F.Q., A.R., M.R., E.M.R., B.R., J.S.R., J.M.R., E.D.R., M.Rodriguear, E.R., H.J.R., R.N.R., D.R.R., M.A.S., M.S., A.J.S., J.A.S., L.S.S., W.W.S., S.H.S., S.Small, A.G.S., J.P.S., Y.E.Song, J.A.Sonnen, S.Spina, P.S.G.- H., R.A.S., A.B.Stevens, S.M.S., D.Sultzer, R.H.S., R.E.T., J.T., G.T., J.Q.T., J.C.T., D.W.T., V.M.VD., L.J.VE., J.M.V., R.Vassar, H.V.V., J.-P.V., S.W., K.A.W.-B., P.L.W., E.M.W., K.C.W., B.W., J.Williamson, H.W., T.S.W., T.W., R.L.W., M.Woon, C.B.W., C.-K.W., S.G.Y., C.-E.Y., L.Y., X.Z., X.Zhou, and C.Z.; *Data Generation:* A.C.N., J.S., Y.Z., D.L.D., J.J.F., K.H.-N., J.Haut, A.B.K., O.V., J.M., L.S.Hynan, C.D.K., J.P.S., A.C.; *Analysis:* F.R., P.B., G.T., N.K., J.S., C.Z., W.-P.L., J.Haut, K.H.-N., N.R.W., Y.Z., J.J.F., M.A.G., Y.Y.L., P.P.K., D.L., E.L.daF., J.B.M., E.L.P., J.P., R.M.S., Y.E.S., X.Z., T.I., O.P., O.V., A.B.K., G.R.J., C.R., and A.C.N. *Manuscript Preparation:* F.R., P.B., G.T., N.K., J.S., C.Z., L.-S.W., R.M., L.A.F., J.L.H., M.A.P.-V., G.D.S., G.R.J., C.R., and A.C.N.; *Study Supervision/ Management:* F.R., P.B., G.T., L.-S.W., L.A.F., J.L.H., R.M., M.A.P.-V., G.D.S., G.R.J., C.R., and A.C.N.

