## Supplementary Materials for "Multi-ancestry genome-wide meta-analysis of 56,241 individuals identifies *LRRC4C, LHX5-AS1* and nominates ancestry-specific loci *PTPRK*, *GRB14*, and *KIAA0825* as novel risk loci for Alzheimer’s disease: the Alzheimer’s Disease Genetics Consortium"

**Supplementary Note 1** . . . . . . . . . . . . . . . . . . . . . . . . . . . . . . . . . . . . . . . . . . . . 3
Supplementary methods and dataset descriptions

**Supplementary Table 1** . . . . . . . . . . . . . . . . . . . . . . . . . . . . . . . . . . . . . . . . . . 32
Basic characteristics of ADGC subjects by ancestry group

**Supplementary Table 2** . . . . . . . . . . . . . . . . . . . . . . . . . . . . . . . . . . . . . . . . . . Excel
Counts of cases/controls and genotyping platforms by dataset

**Supplementary Table 3** . . . . . . . . . . . . . . . . . . . . . . . . . . . . . . . . . . . . . . . . . . Excel
Genome-wide association results for within-ancestry NHW analysis

**Supplementary Table 4** . . . . . . . . . . . . . . . . . . . . . . . . . . . . . . . . . . . . . . . . . . Excel
Genome-wide association results for within-ancestry AFA analysis

**Supplementary Table 5** . . . . . . . . . . . . . . . . . . . . . . . . . . . . . . . . . . . . . . . . . . Excel
Genome-wide association results for within-ancestry HIS analysis

**Supplementary Table 6** . . . . . . . . . . . . . . . . . . . . . . . . . . . . . . . . . . . . . . . . . . Excel
Genome-wide association results for within-ancestry EAS analysis

**Supplementary Table 7** . . . . . . . . . . . . . . . . . . . . . . . . . . . . . . . . . . . . . . . . . . Excel
Genome-wide association results for cross-ancestry analysis

**Supplementary Table 8** . . . . . . . . . . . . . . . . . . . . . . . . . . . . . . . . . . . . . . . . . . Excel
Associations at loci identified in previous cross-ancestry/ancestry-specific GWAS

**Supplementary Table 9** . . . . . . . . . . . . . . . . . . . . . . . . . . . . . . . . . . . . . . . . . . 34
Differential expression in Framingham Heart Study brains

**Supplementary Table 10** . . . . . . . . . . . . . . . . . . . . . . . . . . . . . . . . . . . . . . . . . Excel
Pathway analysis using genes flanking SNPs associated with $P<10^{-6}$

**Supplementary Note 1.** Dataset Descriptions and Supplementary Methods

### Dataset Descriptions

#### *Non-Hispanic Whites (NHW)*

The ADGC NHW dataset comprises subjects from 35 datasets including two waves of the Adult Changes in Thought (ACT) cohort study [ACT1/ACT2]; ten waves of cases and cognitively normal controls from the National Institute on Aging (NIA) Alzheimer Disease Centers (ADCs); the Alzheimer Disease Neuroimaging Initiative (ADNI); the Biomarkers of Cognitive Decline Among Normal Individuals (BIOCARD) Cohort; two waves of the Religious Orders Study/Memory and Aging Project (ROSMAP1-2) and the Chicago Health and Aging Project (CHAP) cohort studies at Rush University; the Einstein Aging Study (EAS); the Multi-Site Collaborative Study for Genotype-Phenotype Associations in Alzheimer's Disease (GenADA) Study by GlaxoSmithKline; Mayo Clinic Jacksonville (MAYO) and Rochester (RMAYO) case-control datasets; the Multi-Institutional Research in Alzheimer's Genetic Epidemiology (MIRAGE) study; the NIA Late-Onset Alzheimer's Disease (LOAD) Family Study (NIA-LOAD); the Netherlands Brain Bank (NBB) case-control dataset; the Oregon Health and Science University (OHSU) case-control dataset; the Pfizer case-control dataset; the Texas Alzheimer's Research and Care Consortium (TARCC) dataset; the Translational Genomics Research Institute series 2 (TGEN2) dataset; the University of Miami (UM)/Case Western Reserve University (CWRU)/Mt. Sinai School of Medicine (MSSM) and UM/CWRU/TARCC wave 2 datasets [UM/CWRU/MSSM and UM/CWRU/TARCC2]; the Universitatsklinikum Saarlandes (UKS) case-control dataset; the University of Pittsburgh (UPITT) case-control dataset; Washington University (WASHU) wave 1 and 2 case-control datasets

[WASHU1/WASHU2]; and the Washington Heights-Inwood Community Aging Project (WHICAP) study datasets.

Descriptions of the ACT1, ADC waves 1-7, ADNI, BIOCARD, CHAP, EAS, GenADA, MAYO, MIRAGE, NBB, NIA-LOAD, OHSU, PFIZER, RMAYO, ROSMAP1, ROSMAP2, TARCC, TGEN2, UKS, UM/CWRU/MSSM, UM/CWRU/TARCC2, UPITT, WASHU1, WASHU2, and WHICAP cohorts were provided in previous ADGC and IGAP studies[1-6]. Here we update descriptions of these studies, where applicable, and provide descriptions for ACT2, ADC wave 8-12. All subjects were recruited under protocols approved by the appropriate Institutional Review Boards (IRBs).

*ACT1/ACT2*: The ACT cohort is an urban and suburban elderly population from a stable HMO that includes 2,581 cognitively intact subjects age ≥ 65 years who were enrolled between 1994 and 1998[7,8]. An additional 811 subjects were enrolled in 2000-2002 using the same methods except oversampling clinics with more minorities. More recently, a Continuous Enrollment strategy was initiated in which new subjects are contacted, screened, and enrolled to keep 2,000 active at-risk person-years accruing in each calendar year. This resulted in an enrollment of 4,146 participants as of May 2009. All clinical data are reviewed at a consensus conference. Dementia onset is assigned half-way between the prior biennial and the exam that diagnosed dementia. A waiver of consent was obtained from the IRB to enroll deceased ACT participants. In total, ACT contributed data on 553 individuals with probable or possible Alzheimer's disease (70 with autopsy-confirmation) and on 1,579 cognitively normal elders (CNEs; 155 with autopsy-confirmation) who were included in the analyses, with 2,103 cases/1,571 CNEs in the first wave (ACT1) and 29 cases/8 CNEs in the second wave (ACT2).

*NIA ADC Samples (ADC1-10)*: The NIA ADC cohort included subjects ascertained and evaluated by the clinical and neuropathology cores of the 32 NIA-funded ADCs. Data collection is coordinated by the National Alzheimer's Coordinating Center (NACC). NACC coordinates collection of phenotype data from the 32 ADCs, cleans all data, coordinates implementation of definitions of Alzheimer's disease cases and controls, and coordinates collection of samples. The ADC cohort consists of 3,311 autopsy-confirmed and 2,889 clinically-confirmed Alzheimer's disease cases, and 247 cognitively normal elders (CNEs) with complete neuropathology data who were older than 60 years at age of death, and 3,687 living CNEs evaluated using the Uniform dataset (UDS) protocol[9,10] who were documented to not have mild cognitive impairment (MCI) and were between 60 and 100 years of age at assessment. Based on the data collected by NACC, the ADGC Neuropathology Core Leaders Subcommittee derived inclusion and exclusion criteria for Alzheimer's disease and control samples. All autopsied subjects were age ≥ 60 years at death. Based on the data collected by NACC, the ADGC Neuropathology Core Leaders Subcommittee derived inclusion and exclusion criteria for Alzheimer's disease and control samples. Alzheimer's disease cases were classified as demented according to NINCDS-ADRDA/DSMIV-V[11] or more recent NIA-AA criteria[12] or Clinical Dementia Rating (CDR) ≥ 137. Neuropathologic stratification of cases followed NIA/Reagan criteria explicitly or used a similar approach when NIA/Reagan criteria were coded as not done, missing, or unknown[13,14]. Cases were intermediate or high likelihood by NIA/Reagan criteria with moderate to frequent amyloid plaques[15] and neurofibrillary tangle (NFT) Braak stage of III-VI[16,17]. Persons with Down's syndrome, non-Alzheimer's disease tauopathies and synucleinopathies were excluded. All autopsied controls had a clinical evaluation within two years of death. Controls did not meet

NINCDS-ADRDA/DSMIV-V criteria for dementia, did not have a diagnosis of mild cognitive impairment (MCI), and had a CDR of 0, if performed. Controls did not meet or were low-likelihood Alzheimer's disease by NIA/Reagan criteria, had sparse or no amyloid plaques, and a Braak NFT stage of 0 – II. ADCs sent frozen tissue from autopsied subjects and DNA samples from some autopsied subjects and from living subjects to the ADCs to the National Cell Repository for Alzheimer's Disease (NCRAD). DNA was prepared by NCRAD for genotyping and sent to the genotyping site at Children's Hospital of Philadelphia. ADC samples were genotyped and analyzed in separate batches (waves 1-10). The ADC data used in IGAP discovery analyses (ADC1-10) consist of 6,292 cases and 4,980 CNEs in total.

*ADNI*: ADNI is a longitudinal, multi-site observational study including Alzheimer's disease, mild cognitive impairment (MCI), and elderly individuals with normal cognition assessing clinical and cognitive measures, MRI and PET scans (FDG and 11C PIB) and blood and CNS biomarkers. For this study, ADNI contributed data on 268 Alzheimer's disease cases with MRI confirmation of Alzheimer's disease diagnosis and 173 healthy controls with Alzheimer's disease-free status confirmed as of most recent follow-up. Alzheimer's disease subjects were between the ages of 55–90, had an MMSE score of 20–26 inclusive, met NINCDS-ADRDA/NIA-AA criteria for probable Alzheimer's disease[11,12], and had an MRI consistent with the diagnosis of Alzheimer's disease. Control subjects had MMSE scores between 28 and 30 and a Clinical Dementia Rating of 0 without symptoms of depression, MCI or other dementia and no current use of psychoactive medications. According to the ADNI protocol, subjects were ascertained at regular intervals over 3 years, but for the purpose of our analysis we only used the final ascertainment status to classify case-control status. Additional details of the study design are available elsewhere[5,18,19].

*BIOCARD*: The BIOCARD study is supported by a grant jointly funded by the National Institute on Aging (NIA) and the National Institute of Mental Health (NIMH). The overarching goal of the BIOCARD Study is to identify biomarkers associated with progression from normal cognitive status to cognitive impairment or dementia, with a particular focus on Alzheimer's Disease. Please see Albert et al. 2014[20] for a detailed description of the study. A total of 354 individuals were initially enrolled in the study. Recruitment was conducted by the staff of the Geriatric Psychiatry Branch (GPB) of the intramural program of the NIMH, beginning in 1995 and ending in 2005. The domains of information collected as part of the study include: cognitive testing, magnetic resonance imaging (MRI), cerebrospinal fluid (CSF), amyloid imaging (using PET-PiB), and blood specimens. Investigators at the Johns Hopkins University School of Medicine began evaluating participants in 2009, and subjects are seen annually. At each visit there are assessments of medical and cognitive status, as well as acquisition of MRI, CSF, PET-PiB, and blood. Each subject in the analyses received a consensus diagnosis by a team of neurologists, neuropsychologists, research nurses and research assistants of the BIOCARD Clinical Core at Johns Hopkins with diagnoses based on evidence of clinical or cognitive dysfunction (i.e., individuals with a CDR score > 0 and/or evidence of decline on cognitive testing). To the extent possible, this diagnosis did not use the cognitive test scores. In brief, (1) clinical data relating to the medical, neurologic and psychiatric status of the subject were examined, (2) reports of changes in cognition by the subject and other sources were examined, and (3) decline in cognitive performance was established. Cognitive test scores were used to: (1)

determine whether the subject had become cognitively impaired, and (2) determine the likely etiology of such impairment. These diagnostic procedures are comparable to those implemented in the Alzheimer's Disease Centers (ADC) program, supported by the NIA. For this study, BIOCARD contributed data on 6 Alzheimer's disease cases and 112 healthy controls with Alzheimer's disease-free status confirmed from the most recent follow-up.

*CHAP*: CHAP is an on-going, community-based study of individuals from a geographically defined community of 3 neighborhoods in Chicago, Illinois (Morgan Park, Washington Heights, and Beverly), with 6,158 participants in the first phase of the study[21] (78.7% overall; 80.5% of the blacks, 74.6% of the whites). Data were collected in cycles of approximately 3 years; each consisting of an in-home interview of all participants and clinical evaluation of a random, stratified sample. The baseline cycle measured disease prevalence and provided risk factor data prior to incident disease onset. A cohort of 3,838 persons free of Alzheimer's disease was identified; 729 persons were sampled for baseline clinical evaluation. Persons in the disease-free cohort had either good cognitive function at baseline, or if cognitive function was intermediate or poor, were free from Alzheimer's disease at the baseline clinical evaluation. This disease-free cohort was evaluated for incident disease after an average of 4.1 years. Sampling for incident clinical evaluation was based on age, sex, race, and change in cognitive function (i.e., stable or improved, small decline, or large decline). The sample set available in the ADGC for genetic analyses included 27 Alzheimer's disease cases and 144 persons free of Alzheimer's disease at time of last assessment. All subjects were age 65 years or older at last evaluation.

*EAS*: Based at the Albert Einstein College of Medicine, the EAS is an ongoing community-based cohort study of cognitive aging and Alzheimer's disease in the elderly which began over four decades ago. Please see Barzilai et al. 2004[22] and Katz et al. 2012[23] for details. The EAS cohort has employed systematic recruiting methods to reduce the selection biases that arise from clinic-based samples and to capture the racial diversity within the Bronx community. Since 1993, a total of 1,944 participants have been enrolled. Between 1993 and 2004, Health Care Financing Administration/Centers for Medicaid and Medicare Services (HCFA/CMS) rosters of Medicare eligible persons aged 70 and above were used to develop sampling frames of community residing participants in Bronx County. Since 2004, New York City Board of Elections registered voter lists for the Bronx have been used due to changes in policies for release of HCFA/CMS rosters. Individuals were mailed introductory letters regarding the study and were then telephoned to complete a brief screening interview. Eligible participants were at least 70 years of age, Bronx residents, non-institutionalized, and English speaking. Exclusion criteria included visual or auditory impairments that preclude neuropsychological testing, active psychiatric symptomatology that interfered with the ability to complete assessments, and non-ambulatory status. Written informed consent was obtained at the initial clinic visit. In-person evaluations were completed at baseline and at subsequent 12-month intervals. Functional status was assessed by the self-administered CERAD C1-ALT, a cognitive/functional impairment instrument, and the Instrumental Activities of Daily Living scale (IADL), a subscale on the Lawton Brody Activities of Daily Living Scale. The score on the IADL was based on 5 domains of function that were common to both elderly men and women. Scores for each domain were dichotomized as impaired vs. not impaired and then the domain scores were summed. If the participant agreed, an informant completed the CERAD C2-ALT, a cognitive/functional impairment instrument, and the Informant Questionnaire on Cognitive Decline in the Elderly (IQ-CODE) 14 forms. The standard neurological physical examination was adapted from the Unified Parkinson's Disease Rating Scale. The evaluation assessed the participant's memory for significant recent events in the news and personal events. The coherence and focus of responses, repetitiveness, and language were determined. When possible, informants were interviewed to ascertain whether they noted any cognitive changes in the participant, and to assess accuracy of the participant's responses. The neurologist also assessed each participant for abnormal behaviors, fluctuation in cognition, and history of sleep disturbance and visual/auditory hallucinations. The neurologist assigned an Hachinski Ischemic Score (HIS), the Clinical Dementia Rating (CDR), and provided a clinical impression of presence or absence of dementia. A diagnosis of dementia was based on standardized clinical criteria from the Diagnostic and Statistical Manual, Fourth Edition (DSM-IV) and required impairment in memory plus at least one additional cognitive domain, accompanied by evidence of functional decline. Diagnoses were assigned at consensus case conferences, which included comprehensive review of cognitive test results, relevant neurological signs and symptoms, and functional status. Memory impairment was defined as scores in the impaired range on any of the memory tests in the neuropsychological battery. (FCSRT $\leq$ 2430 or 1.5 standard deviations (SD) below the age-adjusted mean on Logical Memory) Functional decline was determined at case conference based on information from self or informant report, impairment score on the IADL Lawton Brody Scale, clinical evaluation, and informant questionnaires. Alzheimer's disease was diagnosed in participants with dementia meeting clinical criteria for probable or possible disease established by the NINCDS-ADRDA/ NIA-AA. Incident dementia and Alzheimer's disease were diagnosed in persons free of dementia at baseline who met criteria at follow-up. A subset of individuals who participated in the clinical studies of the EAS came to autopsy, providing an important quality control for diagnostic accuracy. A clinical diagnosis of dementia had a positive predictive value (PPV) of 96% for significant pathology upon autopsy. A clinical diagnosis of possible or probable Alzheimer's disease had a PPV of 79% for the presence of NIA-Reagan intermediate or high likelihood Alzheimer type pathology based on an autopsy sample of 175. For this study, EAS contributed data on 9 Alzheimer's disease cases and 141 healthy controls with Alzheimer's disease-free status confirmed as of most recent follow-up.

*GenADA*: GenADA study data analyzed included 666 Alzheimer's disease cases and 712 CNEs ascertained from nine memory referral clinics in Canada between 2002 and 2005. Patients and CNEs were of non-Hispanic White (NHW) ancestry from Northern Europe. All patients with Alzheimer's disease satisfied NINCDS-ADRDA/NIA-AA[11,12] and DSM-IV criteria for probable Alzheimer's disease with Global Deterioration Scale scores of 3-7. CNEs had MMSE test scores higher than 25 (mean 29.2 ± 1.1), a Mattis Dementia Rating Scale score of ≥ 136, a Clock Test without error,

and no impairments on seven instrumental activities of daily living questions from the Duke Older American Resources and Services Procedures test. Data were collected under an academic-industrial grant from Glaxo-Smith-Kline, Canada by Principal Investigator P. St George-Hyslop. Detailed characteristics of this cohort have been described previously[24].

*MAYO/RMAYO*: All 671 cases and 1,279 controls consisted of NHW subjects from the United States ascertained at the Mayo Clinic[25]. All subjects were diagnosed by a neurologist at the Mayo Clinic in Jacksonville, Florida or Rochester, Minnesota. The neurologist confirmed a Clinical

Dementia Rating score of 0 for all controls; cases had diagnoses of possible or probable Alzheimer's disease made according to NINCDS-ADRDA/NIA-AA criteria[12,25]. Autopsy-confirmed samples (221 cases, 216 CNEs) came from the brain bank at the Mayo Clinic in Jacksonville, FL and were evaluated by a single neuropathologist. In clinically-identified cases, the diagnosis of definite Alzheimer's disease was made according to NINCDS-ADRDA criteria. All Alzheimer's disease brains analyzed in the study had a Braak score of 4.0 or greater. Brains employed as controls had a Braak score of 2.5 or lower but often had brain pathology unrelated to Alzheimer's disease and pathological diagnoses that included vascular dementia, frontotemporal dementia, dementia with Lewy bodies, multi-system atrophy, amyotrophic lateral sclerosis, and progressive supranuclear palsy.

*MIRAGE:* The MIRAGE study is a family-based genetic epidemiology study of Alzheimer's disease that enrolled Alzheimer's disease cases and unaffected sibling controls at 17 clinical centers in the United States, Canada, Germany, and Greece (details elsewhere[26]), and contributed 1,229 subjects (491 Alzheimer's disease cases and 738 CNEs), a subset of the cases and controls that were incorporated into our prior studies[1,5] which met more stringent QC criteria for this study. Briefly, families were ascertained through a proband meeting the NINCDS-ADRDA/NIA-AA[11,12] criteria for definite or probable Alzheimer's disease. Unaffected sibling controls were verified as cognitively healthy based on a Modified Telephone Interview of Cognitive Status score $\geq 86$[28].

*NIA-LOAD*: The NIA LOAD Family Study[29] recruited families with two or more affected siblings with LOAD and unrelated, CNEs similar in age and ethnic background. A total of 1,819 cases and 1,969 CNEs from 1,802 families were recruited through the NIA LOAD study, NCRAD, and the University of Kentucky, with 1,798 cases and 1,568 CNEs included for analysis. One case per family was selected after determining the individual with the strictest diagnosis (definite > probable > possible LOAD). If there were multiple individuals with the strictest diagnosis, then the individual with the earliest age of onset was selected. The controls included only those samples that were neurologically evaluated to be normal and were not related to a study participant.

*NBB*: The Netherlands Brain Bank, which has been previously described elsewhere[3], is a department of the Netherlands Institute for Neuroscience, an institute of the Royal Netherlands Academy of Arts and Sciences. The NBB is a non-profit organization that collects human brain tissue from donors with a variety of neurological and psychiatric disorders and brain tissue from non-diseased donors, as well as anonymized summaries of donors' medical records to be made available for neuroscience research[30]. The sample set available in the ADGC for genetic analyses included 80 pathologically-confirmed Alzheimer's disease cases and 48 subjects free of Alzheimer's pathology at autopsy. All cases were age 65 years or older at time of diagnosis, and all controls were age 65 years or older at time of death.

*OHSU*: The OHSU dataset includes 132 autopsy-confirmed Alzheimer's disease cases and 153 deceased controls that were evaluated for dementia within 12 months prior to death (age at death > 65 years), which are a subset of the 193 cases and 451 controls examined in our previous study[5] meeting more stringent QC criteria in this study. Subjects were recruited from aging research cohorts at 10 NIA-funded ADC, and did not overlap other samples assembled by the ADGC. A more extensive description of control samples can be found elsewhere[31].

*Pfizer:* The Pfizer sample collection comprises Alzheimer's disease cases taken from the Lipitor's Effect in Alzheimer's Disease (LEADe) trial, including subjects who converted to Alzheimer's disease after ascertainment as MCI, as well as 216 probable Alzheimer's disease subjects enrolled by PrecisionMed for a case-control study and 149 subjects from a Phase II trial (#A3041005) of CP-457920 (a selective α5 GABAA receptor inverse agonist) in Alzheimer's disease. Samples were collected from multiple clinical sites, and with appropriate IRB/ethics committee approvals at each individual site, with written and informed consent given by subjects for use in follow-up studies. All subjects were diagnosed with probable or possible Alzheimer's disease if they met NINCDS-ADRDA/NIA-AA[11,12] and/or DSM-IV criteria, and had Mini-Mental Status Exam (MMSE) scores < 25 at baseline. The control group included subjects from two studies: 1) the PrecisionMed case-control study (#A9010012), which recruited elderly subjects free of neurological or psychiatric conditions, and 2) 999-GEN-0583-001, which obtained a reference population of cognitively, neurologically, and psychiatrically normal subjects. Controls have no neuropsychiatric conditions or diseases and had MMSE>27 at the time of enrollment. For Alzheimer's disease analysis, all cases with age-at-onset (AAO) less than 65 years were removed to exclude early-onset Alzheimer's disease subjects. All controls were re-matched with remaining cases according to gender, age (all controls are older than cases), and ethnicity (only individuals with NHW background were analyzed). The final Pfizer Alzheimer's disease case-control GWAS dataset included 696 cases and 762 controls. Cases from the PrecisionMed/ A3041005 and LEADe studies and age-matched controls were genotyped using the Illumina HumanHap550 array. *APOE* genotypes were determined from genotypes for rs429358 and rs7412 obtained using Taqman assays.

*ROSMAP*: ROSMAP are two community-based cohort studies. The ROS has been ongoing since 1993, with a rolling admission. Through July of 2010, 1,139 older nuns, priests, and brothers from across the United States initially free of dementia who agreed to annual clinical evaluation and brain donation at the time of death completed their baseline evaluation. The MAP has been on-going since 1997, also with a rolling admission. Through July of 2010, 1,356 older persons from across northeastern Illinois initially free of dementia who agreed to annual clinical evaluation and organ donation at the time of death completed their baseline evaluation. Details of the clinical and neuropathologic evaluations have been previously reported[32-34]. A total of 1,064 persons passed genotyping QC. Of these, 295 met clinical criteria for Alzheimer's disease at the time of their last clinical evaluation or time of death and met neuropathologic criteria for Alzheimer's disease for those on whom neuropathologic data were available, and 769 were without dementia or MCI at the time of their last clinical evaluation or time of death and did not meet neuropathologic criteria for Alzheimer's disease for those on whom neuropathologic data were available. A second wave of ROSMAP (referred to as ROSMAP2 in this study) included 59 persons who met clinical criteria for Alzheimer's disease at the time of their last clinical evaluation or time of death and met neuropathologic criteria for Alzheimer's disease for those on whom neuropathologic data were available, and 217 persons who were without dementia or MCI at the time of their last clinical evaluation or time of death and did not meet neuropathologic criteria for Alzheimer's disease for those on whom neuropathologic data were available.

*TARCC1/3*: The TARCC is a collaborative Alzheimer’s research effort directed and funded by the Texas Council on Alzheimer’s Disease and Related Disorders (the Council), as part of the Darrell K Royal Texas Alzheimer’s Initiative. Composed of Baylor College of Medicine (BCM), Texas Tech University Health Sciences Center (TTUHSC), University of North Texas Health Science Center (UNTHSC), the UT Southwestern Medical Center at Dallas (UTSW), University of Texas Health Science Center at San Antonio (UTHSCSA), Texas A&M Health Science Center (TAMHSC), and the University of Texas at Austin (UTA), this consortium was created to establish a comprehensive research cohort of well characterized subjects to address better diagnosis, treatment, and ultimately prevention of Alzheimer’s disease[35]. The resulting prospective cohort, the Texas Harris Alzheimer's Research Study, contains clinical, neuropsychiatric, genetic, and blood biomarker data on more than 3,000 participants diagnosed with Alzheimer's disease, mild cognitive impairment (MCI), and cognitively normal individuals. Longitudinal data/sample collection and follow-up on participants occurs on an annual basis. Three waves of sample data from TARCC were examined as part of genetic analyses in the ADGC. Data from the TARCC included 323 cases and 181 CNEs in the first wave (included in the TARCC1 cohort); 84 cases and 115 CNEs in the second wave (included in the UM/CWRU/TARCC2 cohort); and an additional 268 cases and 211 CNEs in TARCC3. All TARCC subjects were greater than 65 years of age at disease onset (cases) or at last disease-free exam (non-cases).

*TGEN2*: Among the TGEN2 data analyzed were 668 clinically- and neuropathologically-characterized brain donors, and 365 CNEs without dementia or significant Alzheimer’s disease pathology. Of these cases and CNEs, 667 were genotyped as a part of the TGEN1 series[36].

Samples were obtained from twenty-one different National Institute on Aging-supported Alzheimer's disease Center brain banks and from the Miami Brain Bank as previously described[36-39]. Additional individual samples from other brain banks in the United States, United Kingdom, and the Netherlands were also obtained in the same manner. The criteria for inclusion were as follows: self-defined ethnicity of European descent, neuropathologically confirmed Alzheimer's disease or neuropathology present at levels consistent with status as a control, and age of death greater than 65. Autopsy diagnosis was performed by board-certified neuropathologists and was based on the presence or absence of the characterization of probable or possible Alzheimer's disease. Where possible, Braak staging and/or CERAD classification were employed. Samples derived from subjects with a clinical history of stroke, cerebrovascular disease, comorbidity with any other known neurological disease, or with the neuropathological finding of Lewy bodies were excluded.

*UKS*: The UKS cohort is a thoroughly diagnosed case-control cohort from Universitatsklinikum Saarlandes, consisting of individuals clinically diagnosed with sporadic Alzheimer's disease (N = 596; age-at-onset [mean ± SD]: 72.2 ± 6.6 years) and cognitively healthy, age-, gender-, and ethnicity-matched population-based controls (N = 170; age-at-exam: 64.1 ± 3.0 years).

*UM/CWRU/MSSM*:  The UM/CWRU/MSSM dataset (formerly UM/VU/MSSM)[40-43] contains 1,177 cases and 1,126 CNEs ascertained at the University of Miami, Case Western Reserve University and Mt. Sinai School of Medicine, including 409 autopsy-confirmed cases and 136 controls, primarily from the Mt. Sinai School of Medicine[44]. An additional 16 cases were included and 34 controls excluded from the data analyzed in the Jun et al. 2010 study[5]. Each affected individual met NINCDS-ADRDA/NIA-AA[11,12] criteria for probably or definite Alzheimer's disease with age at onset greater than 60 years as determined from specific probe questions within the clinical history provided by a reliable family informant or from documentation of significant cognitive impairment in the medical record. Cognitively healthy controls were unrelated individuals from the same catchment areas and frequency matched by age and gender, and had a documented MMSE or 3MS score in the normal range. Cases and controls had similar demographics: both had similar ages-at-onset/ages-at-exam of 71.1 (±17.4 SD) for cases and 73.5 (±10.6 SD) for controls, and cases and controls were 64.5% and 61.3% female, respectively.

*UM/CWRU/TARCC2:* The UMCWRUTARCC2 sample included 256 cases and 189 controls from the University of Miami, Case Western Reserve University, and the Texas Alzheimer's Research Care Consortium (wave 2). All Alzheimer's disease cases had onset of disease symptoms after age 65 years and met NINCDS-ADRDA/NIA-AA[11,12] criteria for probable or possible Alzheimer's disease. Controls were adjudicated to have MMSE scores greater than 28 and no clinically identified signs of cognitive impairment. Additional details of subject recruitment at these sites are described in the UM/CWRU/MSSM (formerly UM/VU/MSSM) and TARCC cohort descriptions in this supplement and elsewhere[1,3,4].

*UPITT*: The University of Pittsburgh dataset contains 1,255 NHW Alzheimer's disease cases (of which 277 were autopsy-confirmed) recruited by the University of Pittsburgh Alzheimer's Disease Research Center, and 829 NHW, CNEs ages 60 and older (2 were autopsy-confirmed). All Alzheimer's disease cases met NINCDS-ADRDA/NIA-AA[11,12] criteria for probable or definite Alzheimer's disease. Additional details of the cohort used for GWAS have been previously published[45].

*WASHU*: An NHW LOAD case-control dataset consisting of 377 cases and 281 healthy elderly controls was used in analyses for this study. This dataset was split between two analysis datasets (WASHU1 and WASHU2). Participants were recruited as part of a longitudinal study of healthy aging and dementia. Diagnosis of dementia etiology was made in accordance with standard criteria and methods[10]. Severity of dementia was assessed using the Clinical Dementia Rating scale[46].

*WHICAP*: WHICAP[47,48] is a community-based longitudinal study of aging and dementia among elderly, urban-dwelling residents. Beginning enrolment in 1989, WHICAP has followed more than 5,900 residents over 65 years of age, including white, African American, and Hispanic participants. Detailed clinical assessments were performed at approximately 24-month intervals over the seven years of the initial study. All interviews were conducted in either English or Spanish. The choice of language was decided by the subject to ensure the best performance, and the majority of assessments were performed in the subject's home, which included medical, neurological, and neuropsychological evaluations. Results of the neurological, psychiatric, and neuropsychological assessments were reviewed in a consensus conference comprised of neurologists, psychiatrists, and neuropsychologists. Based on this review all participants were assigned to one of three categories: dementia, cognitive impairment, or normal cognitive function. The sample set available in the ADGC for genetic analyses included 73 Alzheimer's disease cases and 560 subjects with normal cognitive function.

*African American (AFA)*

The study included subjects from the Adult Changes in Thought (ACT) Study[7], the National Institute on Aging (NIA) Alzheimer's Disease Centers (ADCs)[9], the University of Miami/Case Western Reserve University[41,42] (UM/CWRU), the Mount Sinai School of Medicine (MSSM) Brain Bank[44], the Washington Heights Inwood Columbia Aging Project (WHICAP)[49], The African American Alzheimer's Disease Genetics (AAG) Study[50], the MIRAGE Study[26], NIA-LOAD/NCRAD[29], the Mayo Clinic[25], the Rush University Alzheimer's disease Center (ROS/MAP, MARS/CORE)[32-34,51],the Chicago Health and Aging Project (CHAP)[21,52], the Indianapolis Ibadan Dementia Study (IU)[53], the Genetic and Environmental Risk Factors for Alzheimer's Disease Among African Americans (GenerAAtions) Study[54], the University of Pittsburgh (UPITT)[45], and Washington University (WASHU)[10,55-57]. As described in the main text, the analyses were restricted to individuals of African American ancestry. All subjects were recruited under protocols approved by the appropriate Institutional Review Boards.

*ACT*: The ACT cohort[7] is an urban and suburban elderly population from a stable HMO. The original cohort of 2,581 cognitively intact participants age ≥ 65 were enrolled between 1994 and 1998; of these 4% were African American. An additional 811 participants were enrolled in 2000-2002 using the same methods except oversampling clinics with more minorities, resulting in an overall rate of 5% African Americans. More recently, a continuous enrollment strategy was initiated in which new participants are contacted, screened, and enrolled to keep 2,000 active at-risk person-years accruing in each calendar year. This resulted in an overall enrollment of 4,729 participants as of June 2012, of whom 193 (4.1%) were African American. All clinical data are reviewed at a consensus conference. Dementia onset is assigned halfway between the prior biennial and the exam that diagnosed dementia. Enrollment for the eMERGE Study began in 2007. A waiver of consent was obtained from the IRB to enroll deceased ACT participants, and consent for data sharing was obtained from living participants. In total, ACT/eMERGE contributed data on 32 individuals with probable or possible AD and on 65 CNEs who were included in analyses.

*AAG*: Participants of the multisite AAG study[50] that contributed to this study were recruited between 2008 and 2011 from communities surrounding four locations: Columbia University in New York City, NY; North Carolina State A&T University in Greensboro, NC; University of Miami in Miami, FL; and Vanderbilt University in Nashville, TN. Participants were recruited from various sources, including naturally occurring retirement communities, churches, Black fraternal and other organizations, community centers, health fairs, physician's offices, newspaper ads, and word of mouth. All participants were age 60 and older and described themselves as non-Hispanic and Black. A one-time, in-person evaluation included a comprehensive neuropsychological test battery, a medical and neurological examination, and assessment of memory complaints, as well as an informant interview assessing functional status and possible change in cognitional and daily activities. These data were evaluated in a consensus conference and diagnoses were based on standard research criteria[12] and categorized according to National Alzheimer's Coordinating Center criteria. Blood was drawn and sent to the National Cell Repository for Alzheimer's Disease (NCRAD). The current study included 624 people with AD and 161 controls from the AAG cohort. DNA was prepared by NCRAD for genotyping and sent to the genotyping site at the Children's Hospital of Philadelphia.

*ADCs*: The NIA ADC cohort[9] included subjects ascertained and evaluated by the clinical and neuropathology cores of the 35 NIA-funded ADCs. Data collection was coordinated by NACC, which collected phenotype data from the ADCs, then cleaned all data, adjudicated AD affection status controls, and coordinated biological specimen collection including whole blood. The ADC cohort consists of 53 autopsy AD cases, 612 clinical AD cases, and 1,149 autopsy-confirmed or clinically-confirmed or cognitively-normal elders (CNEs) of self-reported African American background who were older than 60 years of age at death or at assessment. Protocols are consistent with ADC data collection described above. The subjects included in this study were genotyped in six waves.

*CHAP*: The Chicago Health and Aging Project (CHAP) is a longitudinal cohort study[21,52] of all participating residents 65 years of age and older of a geographically-defined biracial community located on the southwest side of Chicago. At each of six data collection cycles (every three years), all subjects have undergone brief cognitive testing and a stratified random sample of about 500-600 subjects (aggregate 2844) has undergone detailed clinical evaluation. The subjects provided for analysis were diagnosed with prevalent or incident Alzheimer's disease at these clinical evaluations.

*GenerAAtions*: Participants of the GenerAAtions Study[54] were identified through the electronic claims database of the Henry Ford Health System. Community-dwelling African Americans aged 65 years and older who had at least one encounter with the Henry Ford Health System in the three years prior to their recruitment and who had an available proxy informant were eligible for this study. Cases met NINCDS/ADRDA criteria[11] for possible or probable AD, determined in a consensus conference which included a behavioral neurologist, psychiatrist, neuropsychologist, and a behavioral neurology nurse practitioner. Phenotypic and GWAS data were available for 242 AD cases and 204 cognitively normal controls. GWAS genotyping of this sample was performed using the Illumina 660 chip as previously described[54].

*IU*:  The African American participants of the Indianapolis Cohort of the Indianapolis Ibadan Dementia Study at Indiana University[53] included in this study (173 cases, 1002 controls) were part of the community-based longitudinal comparative epidemiological study of African Americans in Indianapolis, and Yoruba Nigerians living in the city of Ibadan. In 1992, enrollment staff employed home visits to randomly sampled residential addresses in 29 contiguous U.S. Census tracts. Entry criteria were age ≥ 65 years, self-identified African American, and lived at the sampled address. At that time, 2,212 participants were enrolled. In 2001, new participants were enrolled using random sampling from Medicare rolls, with entry criteria of age ≥ 70 years and were self-identified as African American. At the time, 1,892 participants were enrolled. Participants were evaluated every two to three years with the Community Screening Interview for Dementia (CSI-D). Based on CSI-D scores individuals were selected for a full diagnostic clinical assessment including: CERAD neuropsychological battery, physical and neurological exam, and informant interview. Diagnoses were made by a panel of clinicians using standard criteria.

*MAYO*: There were 64 cases and 195 CNEs included from the Mayo Clinic[25]. All subjects were diagnosed by a neurologist at the Mayo Clinic in Jacksonville, Florida or Rochester, Minnesota.

The neurologist confirmed a Clinical Dementia Rating score of 0 for all controls; cases had diagnoses of possible or probable AD made according to NINCDS-ADRDA criteria[11].

*MIRAGE*: The MIRAGE study[26] is a family-based genetic epidemiological study of AD that enrolled AD cases and unaffected sibling controls at 17 clinical centers in the United Stat es, Canada, Germany, and Greece, and contributed 51 African American cases and 65 CNEs that were genotyped on the Illumina 300k chip and 188 African American cases and 236 CNEs that were genotyped on the Illumina 660k chip. In brief, families were ascertained through a proband meeting the NINCDS-ADRDA criteria for definite or probable AD. Unaffected sibling controls were verified as cognitively healthy based on a Modified Telephone Interview of Cognitive Status score ≥ 86.

*MSSM*: The Mount Sinai School of Medicine dataset[44] contains 29 African American AD cases (all neuropathologically confirmed) and 14 CNEs (all neuropathologically confirmed), recruited to the Mount Sinai Brain Bank. Subjects had been residents of the Jewish Home and Hospital in Manhattan and The Bronx, NY and were participants in a longitudinal study of aging and dementia[34]. Brains were donated by the next of kin of deceased residents. AD diagnoses were based on clinical assessment including neuropathological assessments and subjects met CERAD criteria for definite AD or probable AD. CDR assessments, based on cognitive and functional status during the last six months of life, had been performed for every subject.

*NIA-LOAD/NCRAD*: The NIA-LOAD Family Study[29] recruited families with two or more affected siblings with LOAD and unrelated, CNEs similar in age and ethnic background. A total of 35 African American familial cases and 61 unaffected individuals were recruited through the NIA-LOAD study, NCRAD, and the University of Kentucky and included for analysis. One case per family was selected after determining the individual with the strictest diagnosis (definite > probable > possible LOAD). If there were multiple individuals with the strictest diagnosis, then the individual with the earliest age of onset was selected. The controls included only those samples that were neurologically evaluated to be normal and were not related to a study participant.

*The Rush University Studies (ROS/MAP/MARS/CORE)*: ROS/MAP are two community-based cohort studies[32-34]. The ROS has been on-going since 1993, with a rolling admission. Through July of 2010, 1,147 older nuns, priests, and brothers from across the United States initially free of dementia who agreed to annual clinical evaluation and brain donation at the time of death completed their baseline evaluation. Of these, 89 self-reported African Americans were included in the current study. The MAP has been on-going since 1997, also with a rolling admission. Through July of 2010, 1,392 older persons from across northeastern Illinois initially free of dementia who agreed to annual clinical evaluation and organ donation at the time of death completed their baseline evaluation and 97 self-reported African Americans were included in this meta-analysis. Details of the clinical and neuropathologic evaluations have been previously reported[32-34]. A total of 130 persons passed genotyping QC. Of these, 30 met clinical criteria for AD at the time of their last clinical evaluation or time of death and met neuropathologic criteria for AD for those on whom neuropathologic data were available, and 100 were without dementia or MCI at the time of their last clinical evaluation or time of death and did not meet neuropathologic criteria for AD for those on whom neuropathologic data were available.

MARS[51] is a community-based cohort study of older African Americans with a rolling admission. Through July of 2010, 356 self-reported African Americans without known dementia who agreed to annual clinical evaluation completed their baseline evaluation. CORE[51] is a community-based cohort study of older African Americans with and without dementia at baseline. Through July 2010, CORE has enrolled 218 older Africans without dementia at baseline.

*UM/CWRU*: The University of Miami/Case Western Reserve University dataset[41,42] contains 110 African American cases and 189 CNEs ascertained at the University of Miami and Vanderbilt University. Each affected individual met NINCDS-ADRDA criteria for probable or definite AD with age at onset greater than 60 years as determined from specific probe questions within the clinical history provided by a reliable family informant or from documentation of significant cognitive impairment in the medical record. Cognitively healthy controls were unrelated individuals from the same catchment areas and frequency matched by age and sex and had a documented MMSE or 3MS score in the normal range.

*UPITT*: The University of Pittsburgh dataset contains 114 African American AD cases (of which 6 confirmed) recruited by the University of Pittsburgh Alzheimer's Disease Research Center, and 79 African American CNEs ages 60 and older (2 were autopsy-confirmed). All AD cases met NINCDS-ADRDA criteria for probable or definite AD[11].

*WHICAP*: African American participants from the Washington Heights-Inwood Columbia Aging Project that were included in the present study (170 cases, 299 controls) were part of a longitudinal cohort study enrolled by a random sampling of Medicare recipients 65 years or older residing in northern Manhattan, New York[47,49]. Each participant underwent an interview of general health and function, medical history, a neurological examination, and a neuropsychological battery. Baseline data were collected from 1999 through 2001. Follow-up data were collected at sequential intervals of 18 months. Diagnosis of dementia etiology was made based on standard criteria[12], and severity of was assessed using the Clinical Dementia Rating scale.

*WASHU*: An African American LOAD case-control dataset consisting of 87 cases and 30 healthy elderly controls was used in analyses for this study[10,55-57]. Participants were recruited as part of a longitudinal study of healthy aging and dementia. Diagnosis of dementia etiology was made in accordance with standard criteria and methods[12]. Severity of dementia was assessed using the Clinical Dementia Rating scale.

***Hispanic (HIS)***

*ADCs*: As for other ancestry groups, the NIA ADC cohort[9] included subjects ascertained and evaluated by the clinical and neuropathology cores of the 35 NIA-funded ADCs. Data collection was coordinated by NACC, which collected phenotype data from the ADCs, then cleaned all data, adjudicated AD affection status controls, and coordinated biological specimen collection including whole blood. The ADC cohort consists of 328 autopsy autopsy-confirmed or clinically-confirmed AD cases and 364 autopsy-confirmed or clinically-confirmed cognitively-normal elders (CNEs) of self-reported Hispanic ethnicity and any self-reported race classification who were older than 60 years of age at death or at assessment. Protocols are consistent with

ADC data collection described above. The subjects included in this study were genotyped over 12 waves and were divided into five harmonized subject sets.

*Estudio Familiar de Influencia Genetica en la Enfermedad de Alzheimer (EFIGA):* EFIGA includes 683 at-risk family members from 242 AD-affected families of Caribbean Hispanic descent, where each family has two or more AD-affected individuals. This study was initiated in 1998 and recruited subjects from the Taub Institute for Research on Alzheimer's Disease and the Aging Brain in New York as well as from clinics in the Dominican Republic with the help of local physicians and the Dominican Society of Geriatrics and Gerontology. All affected and unaffected family members in the Dominican Republic and New York were evaluated in person, with cases defined using the NINCDS-ADRDA criteria for probable or possible LOAD and the CDR was used to rate the severity of dementia, with additional review of brain imaging and other laboratory study results when available. All AD patients underwent standardized neurological and neuropsychological evaluations. Structured family history interviews were then conducted with available family members to determine whether patients had living siblings or relatives with the disease. Medical and neurological examinations were completed for all family members. Brains of participants with dementia and history of stroke were administered magnetic resonance imaging scans to exclude patients with comorbid cerebrovascular disease. DNA samples and cell lines were collected and stored for all participating individuals.

*WHICAP:* Hispanic participants from the Washington Heights-Inwood Columbia Aging Project that were included in the present study (512 cases, 1,187 controls) were part of a longitudinal cohort study enrolled by a random sampling of Medicare recipients 65 years or older residing in northern Manhattan, New York. Each participant underwent an interview of general health and function, medical history, a neurological examination, and a neuropsychological battery. Baseline data were collected from 1999 through 2001. Follow-up data were collected at sequential intervals of 18 months. Diagnosis of dementia etiology was made based on standard criteria and severity of was assessed using the Clinical Dementia Rating scale.

*The Puerto Rican 10/66 Study (PR1066):* is an international study of AD begun in 2007 (Dr. Ivonne Jimenez-Velazquez, PI) as part of the 10/66 Dementia Research Group (https://www.alz.co.uk/1066/). The 10/66 dementia study is a population-based research study focusing on developing nations, which represent 66% of people with dementia but less than one-tenth of population-based research conducted in developing nations (China, India, Latin America)[58,59]. Individuals were recruited as part of the 10/66 Population-Based Study of Dementia using standard protocols[58]. As part of this study, sociodemographic information and detailed clinical history of memory decline were collected. In addition, the Clinical Dementia Rating scale (CDR), Community Screening Interview for Dementia, and Petersen ADL criteria were collected for all individuals. Neurocognitive testing (CERAD battery) was available for some participants. All participants were adjudicated for dementia. The study protocol for the 10/66 population-based study and the consent procedures were approved by the King's College London research ethics committee and University of Puerto Rico, Medical Sciences Campus Institutional Review Board (IRB). Informed consent was documented in writing in all cases.

*The Puerto Rican Alzheimer's Disease Initiative (PRADI):* PRADI[60] is an NIH/NIA study of late-onset AD focused on the Caribbean-Hispanic population of Puerto Rico and the Puerto Rican diaspora in the continental US. Participants including cases and cognitively-intact, similarly-aged controls were ascertained for an AD/ADRD memory study and eligibility was based on self-reported Puerto Rican heritage. Recruitment was performed predominantly in Puerto Rico with a small proportion of subjects ascertained in South Florida, New York, and Connecticut. Participants were ascertained and evaluated through community centers, private memory clinics, and adult day care centers, and some participants received in-home evaluations. All participants greater than 60 years of age underwent a standard clinical evaluation consisting of a medical and family history interview, neuropsychological testing, behavioral and emotional assessments, and functional measures. Venous blood samples (or saliva samples when needed) were collected on all participants. All assessments were conducted in the preferred language of the participant or knowledgeable informant.

***East Asian (EAS)***

*ASA-JPN:* Clinically defined subjects were recruited by the Japanese Genetic Study Consortium of Alzheimer's Disease (JGSCAD: principal investigator, Y.I.)[61,62] Probable AD cases were ascertained on the basis of the criteria of the National Institute of Neurological and Communicative Disorders, and Stroke-Alzheimer's Disease and Related Disorders (NINCDS/ADRDA)[11]. The Mini-Mental State Examination[63], Clinical Dementia Rating[64], and/or Function Assessment Staging[65] were primarily used for evaluation of cognitive impairment. Elders living in an unassisted manner in the local community with no signs of dementia were used as controls. DNA was extracted from peripheral blood leukocytes using standard protocols[61].

*ADCs*: The NIA ADC cohort[9] included East Asian subjects ascertained and evaluated by the clinical and neuropathology cores of the 35 NIA-funded ADCs. The ADC East Asian cohort consists of 94 autopsy-confirmed or clinically-confirmed AD cases, and 160 autopsy-confirmed or clinically-confirmed cognitively-normal elders (CNEs) of self-reported Asian ancestry who were older than 60 years at death or at assessment. East Asian ancestry subjects were included in four ADC dataset waves (ADC9-12) and genotype data for these subjects underwent QC by genotyping batch/wave along with subjects from multiple ancestry groups to identify genotyping quality issues. Once batch-specific QC steps were performed, subjects of self-reported Asian ancestry were extracted into a separate subset for variant-level assessment of Hardy-Weinberg equilibrium, and within-ancestry population substructure estimation.

**Supplementary Table 1.** Basic characteristics of ADGC subjects by ancestry group

| Ancestry | Total | Cases (n) | Female (%) | Mean AAO (SE) | Controls (n) | Female (%) | Mean AAE (SE) |
|---|---|---|---|---|---|---|---|
| Non-Hispanic White (NHW) | 37,382 | 17,839 | 57.6% | 73.2 (8.5) | 19,543 | 59.5% | 75.7 (8.0) |
| African American (AFA) | 6,728 | 2,114 | 68.7% | 78.6 (8.0) | 4,614 | 72.1% | 76.6 (8.2) |
| Hispanic (HIS) | 8,899 | 3,005 | 68.1% | 79.9 (8.0) | 5,894 | 68.1% | 72.2 (8.0) |
| East Asian (EAS) | 3,232 | 1,430 | 70.9% | 73.5 (4.9) | 1,802 | 52.8% | 76.3 (6.6) |
| All ADGC Subjects | 56,241 | 24,388 | 60.5% | 74.4 (8.2) | 31,853 | 62.4% | 75.3 (8.0) |

**Supplementary Tables in Excel Spreadsheet**

**Supplementary Table 2.** Counts of cases and controls/non-cases and genotyping platforms by dataset within each ancestry group

**Supplementary Table 3.** Genome-wide association findings with $P<10^{-5}$ in within-ancestry meta-analyses of non-Hispanic white (NHW) subjects **(a)** with adjustment for age-at-onset (cases)/age-at-last exam (controls), sex, and PCs for population substructure and **(b)** with additional adjustment for dosage of the *APOE* ε4 allele.

**Supplementary Table 4.** Genome-wide association findings with $P<10^{-5}$ in within-ancestry meta-analyses of African American (AFA) subjects **(a)** with adjustment for age-at-onset (cases)/age-at-last exam (controls), sex, and PCs for population substructure and **(b)** with additional adjustment for dosage of the *APOE* ε4 allele.

**Supplementary Table 5.** Genome-wide association findings with $P<10^{-5}$ in within-ancestry meta-analyses of Hispanic (HIS) subjects **(a)** with adjustment for age-at-onset (cases)/age-at-last exam (controls), sex, and PCs for population substructure and **(b)** with additional adjustment for dosage of the *APOE* ε4 allele.

**Supplementary Table 6.** Genome-wide association findings with $P<10^{-5}$ in within-ancestry meta-analyses of East Asian (EAS) subjects **(a)** with adjustment for age-at-onset (cases)/age-at-last exam (controls), sex, and PCs for population substructure and **(b)** with additional adjustment for dosage of the *APOE* ε4 allele.

**Supplementary Table 7.** Genome-wide association findings with $P<10^{-5}$ from cross-ancestry meta-analyses **(a)** with adjustment for age-at-onset (cases)/age-at-last exam (controls), sex, and PCs for population substructure and **(b)** with additional adjustment for dosage of the *APOE* ε4 allele.

**Supplementary Table 8.** Associations at loci identified in previous cross-ancestry/ancestry-specific GWAS.

**Supplementary Table 9.** Differential expression of genes at/near novel loci with AD-related traits in FHS

| Gene | Chr | AD vs. Control | | Braak Stages | | | Plaque Scores | | |
|---|---|---|---|---|---|---|---|---|---|
| | | **$Log_2FC$** | ***P*** | **β** | **SE** | ***P*** | **β** | **SE** | ***P*** |
| *GRB14* | 2 | -0.006 | 0.224 | -0.084 | 0.076 | 0.266 | 0.035 | 0.087 | 0.683 |
| *KIAA0825* | 5 | -0.008 | 0.0502 | -0.079 | 0.050 | 0.114 | 0.017 | 0.057 | 0.768 |
| *PTPRK* | 6 | -0.004 | 0.0702 | -0.030 | 0.037 | 0.409 | 0.017 | 0.044 | 0.704 |
| *API5* (flanking *LRRC4C*) | 11 | -0.002 | 0.142 | -0.085 | 0.035 | 0.0158 | 0.022 | 0.039 | 0.581 |
| *LRRC4C* | 11 | **-0.010** | **$8.79\times10^{-4}$** | -0.111 | 0.039 | 0.00465 | 0.064 | 0.044 | 0.151 |
| *LHX5* (flanking *LHX5-AS1*) | 12 | 0.009 | 0.783 | -- | -- | -- | -- | -- | -- |
| *LHX5-AS1* | 12 | -0.006 | 0.935 | -- | -- | -- | -- | -- | -- |
| *RBM19* (flanking *LHX5-AS1*) | 12 | 0.000 | 0.793 | -0.013 | 0.041 | 0.754 | -0.015 | 0.046 | 0.741 |

**Supplementary Tables in Excel Spreadsheet**

**Supplementary Table 10.** Pathways using genes flanking a SNP with $P<10^{-6}$ in the **(a)** *APOE*-unadjusted and **(b)** *APOE*-adjusted (b) models.

**Supplementary Table 11.** Summary of common pathways with $P_{adjusted}$<0.05 across different ancestries from Venn diagram in **Supplementary Figure 17a**.

| Rank | Ancestry and Pathways with $P_{adjusted}$<0.05 |
|---|---|
| **Shared pathways across 4 ancestries from NHW, AFA, HIS, and EAS** | |
| 1 | chylomicron remnant clearance (GO:0034382) |
| 2 | positive regulation of cholesterol esterification (GO:0010873) |
| 3 | regulation of cholesterol esterification (GO:0010872) |
| 4 | phospholipid efflux (GO:0033700) |
| 5 | positive regulation of steroid metabolic process (GO:0045940) |
| 6 | high-density lipoprotein particle remodeling (GO:0034375) |
| 7 | cholesterol efflux (GO:0033344) |
| **Shared pathways across AFA, HIS, and EAS** | |
| 1 | cholesterol transport (GO:0030301) |
| 2 | phospholipid transport (GO:0015914) |
| **Shared pathways between NHW and EAS** | |
| 1 | positive regulation of natural killer cell mediated cytotoxicity (GO:0045954) |
| **Shared pathways between AFA and EAS** | |
| 1 | acylglycerol metabolic process (GO:0006639) |
| 2 | triglyceride metabolic process (GO:0006641) |
| 3 | positive regulation of cellular metabolic process (GO:0031325) |

**Supplementary Figure 1.** Manhattan plots of ancestry-specific meta-analysis results for the *APOE* ε4 dosage-unadjusted model in NHW **(a)**, HIS **(b)**, AFA **(c)**, and EAS **(d)**.

**(a)** NHW

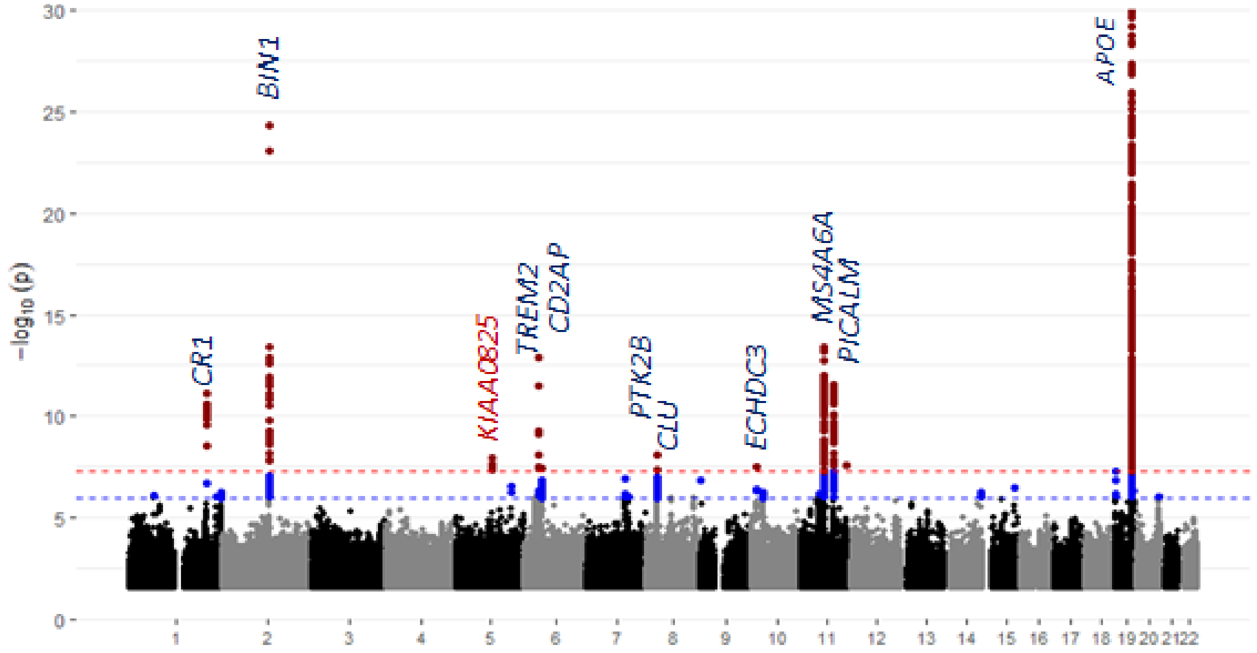

**(b)** HIS

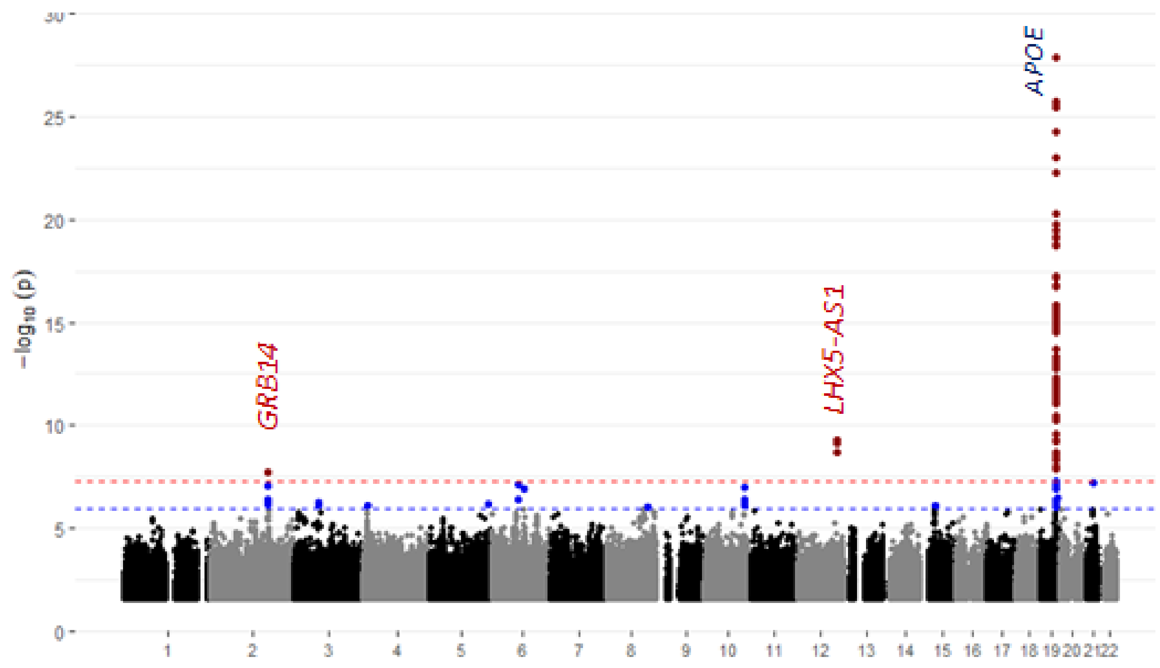

**(c)** AFA

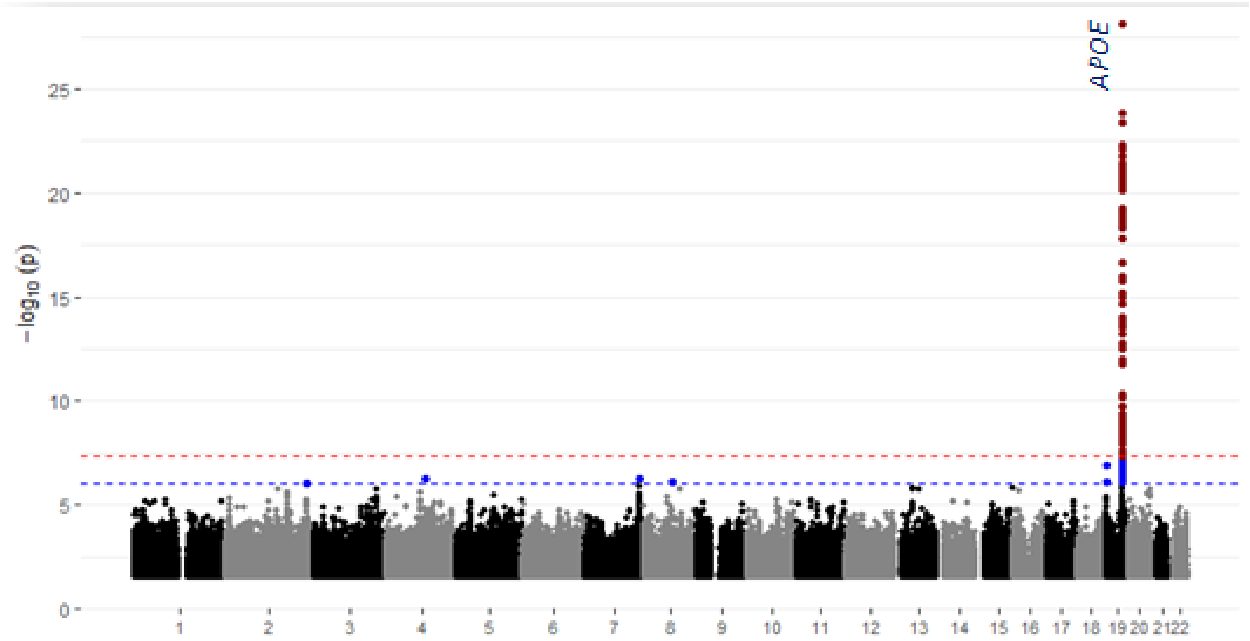
APOE
25
20
15
10
5
0
−log10 (p)

**(d)** EAS

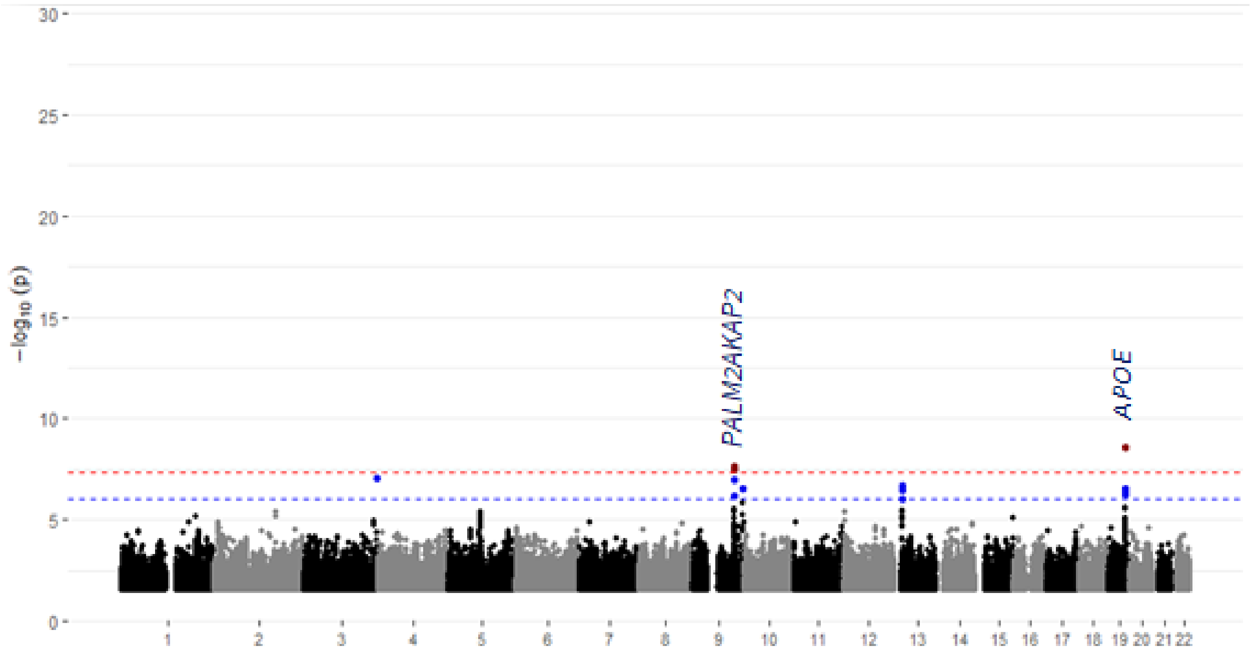
30
25
20
15
10
5
0
−log10 (p)
PALM2AKAP2
APOE

**Supplementary Figure 2.** Manhattan plots of ancestry-specific meta-analysis results for the *APOE* ε4 dosage-adjusted model in NHW **(a)**, HIS **(b)**, AFA **(c)**, and EAS **(d)**.

**(a)** NHW

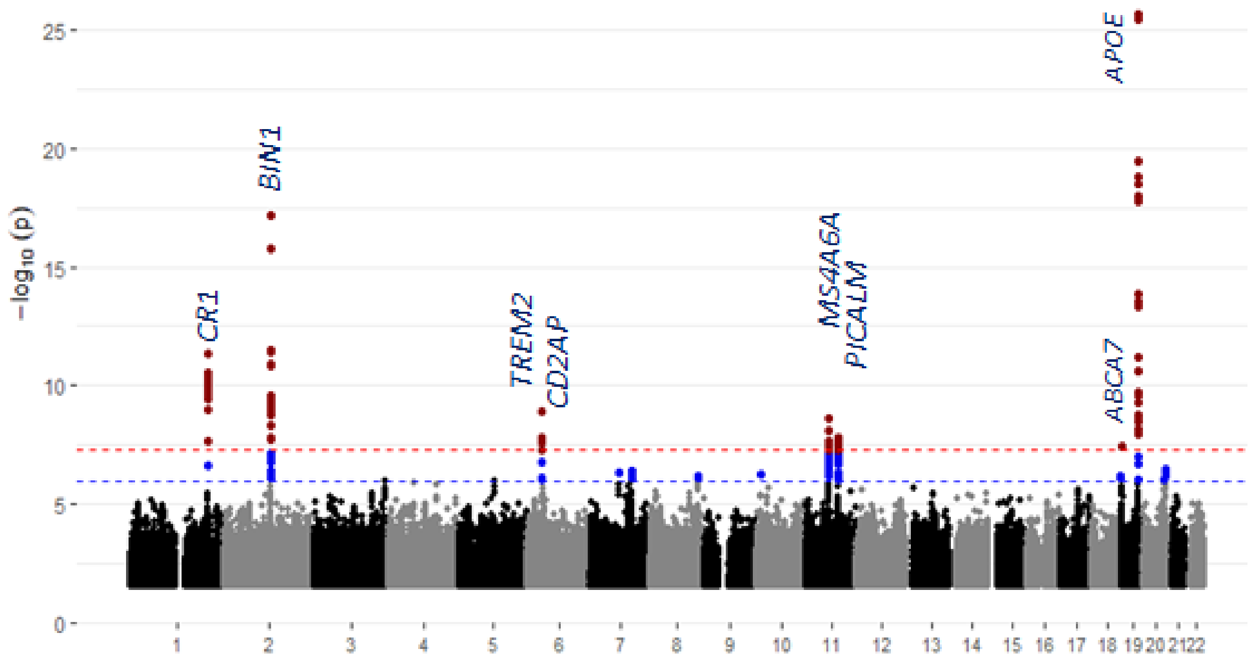

**(b)** HIS

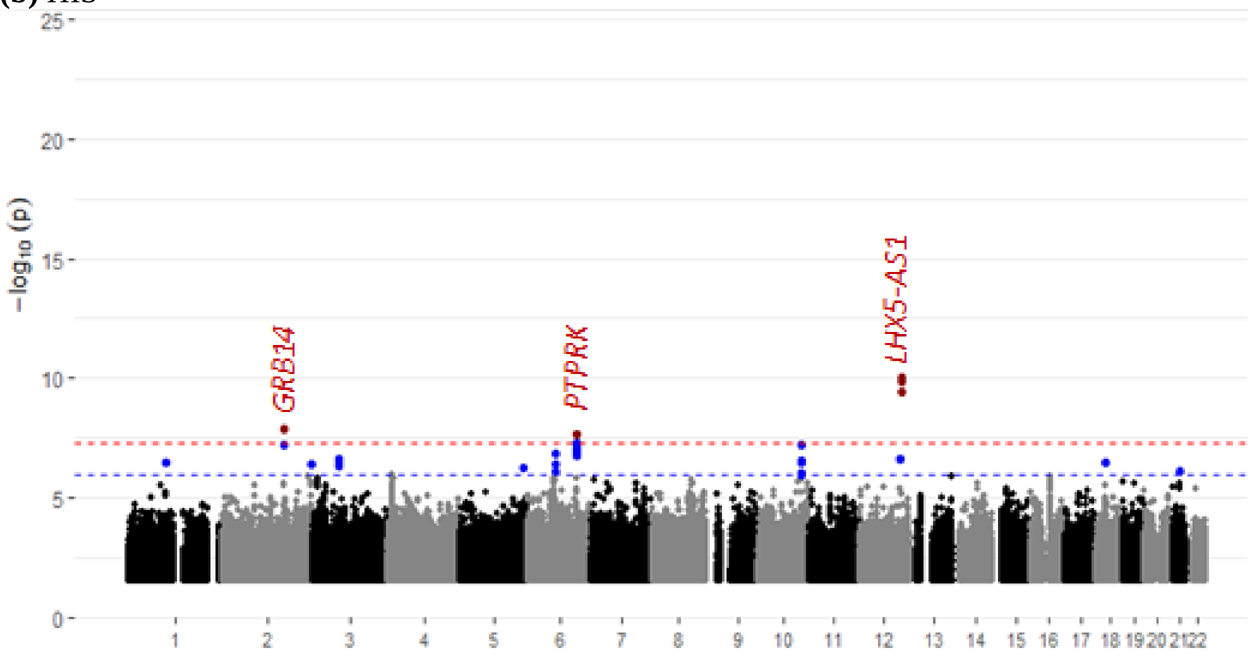

**(c)** AFA

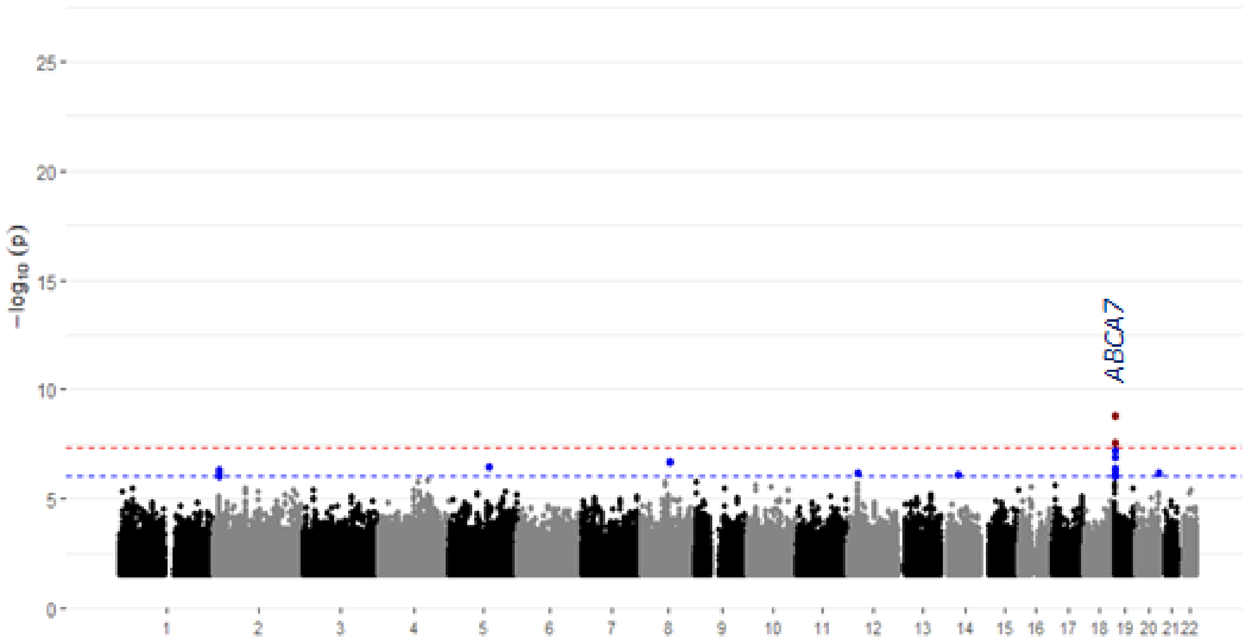

ABCA7
−log10 (p)
25
20
15
10
5

**(d)** EAS

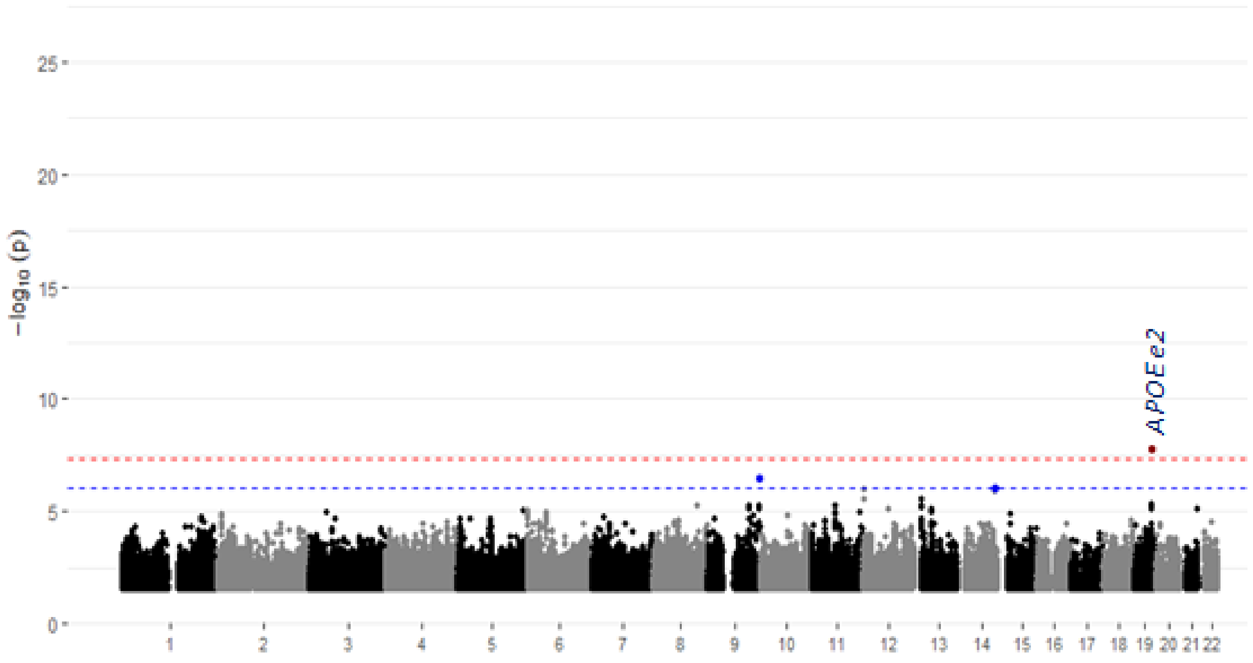

APOE e2
−log10 (p)
25
20
15
10
5

**Supplementary Figure 3.** Quantile-quantile (Q-Q) plots of ancestry-specific meta-analysis results for the *APOE* ε4 dosage-unadjusted model in NHW **(a)**, HIS **(b)**, AFA **(c)**, and EAS **(d)**.

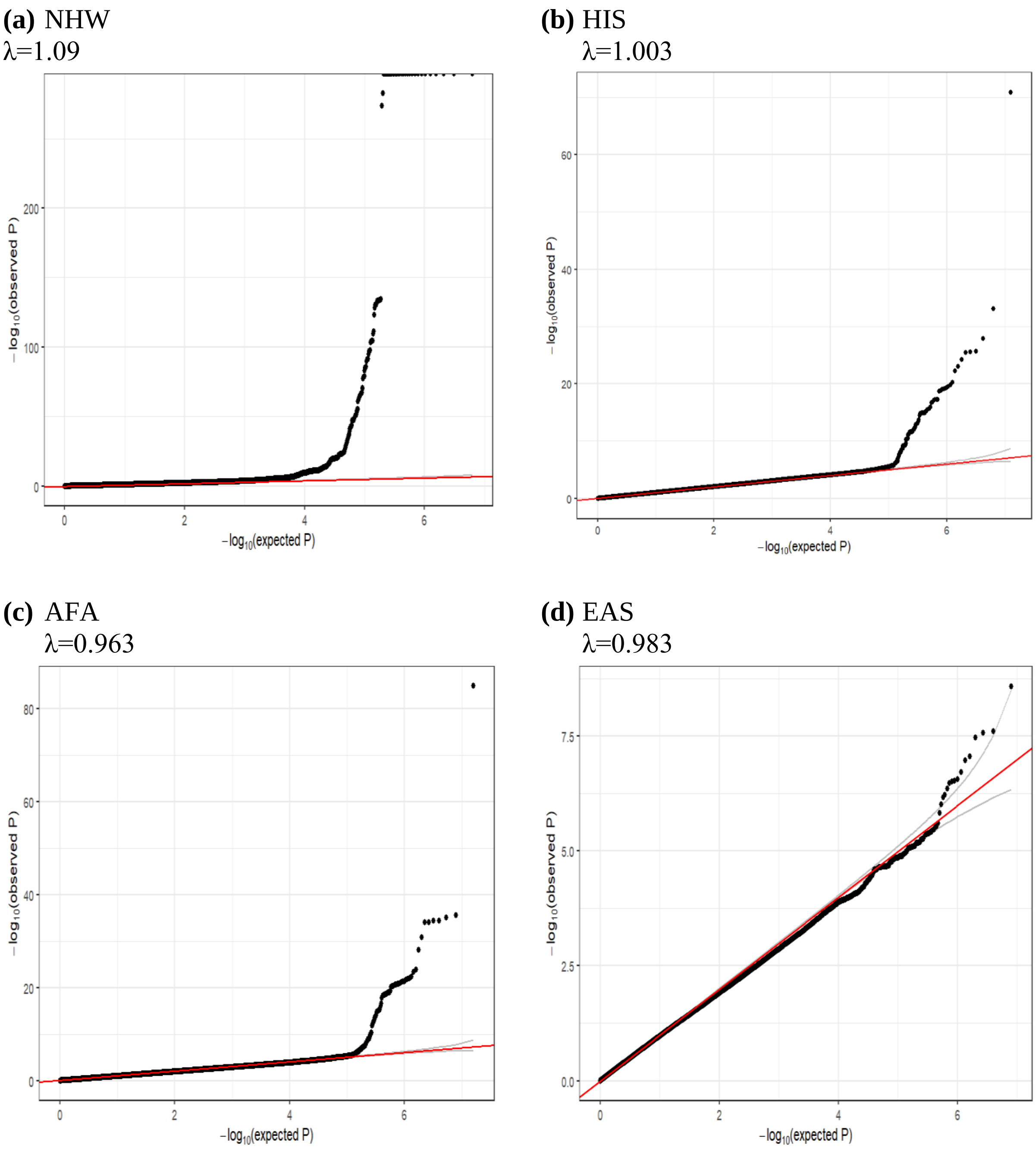

**Supplementary Figure 4.** Quantile-quantile (Q-Q) plots of ancestry-specific meta-analysis results for the *APOE* ε4 dosage adjusted model in NHW **(a)**, HIS **(b)**, AFA **(c)**, and EAS **(d)**.

**(a)** NHW
λ=1.06

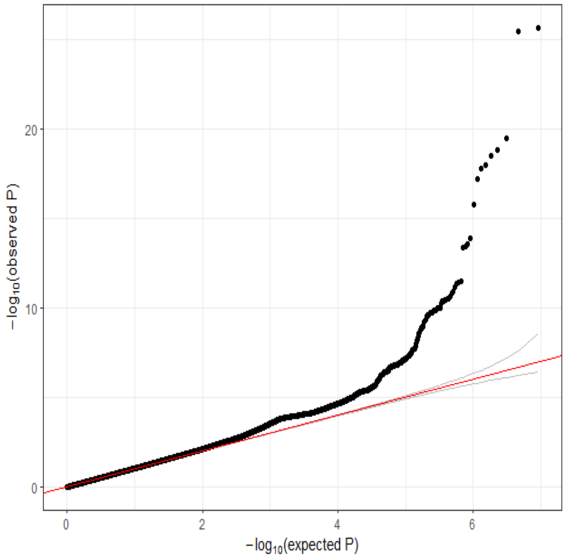

**(b)** HIS
λ=1.003

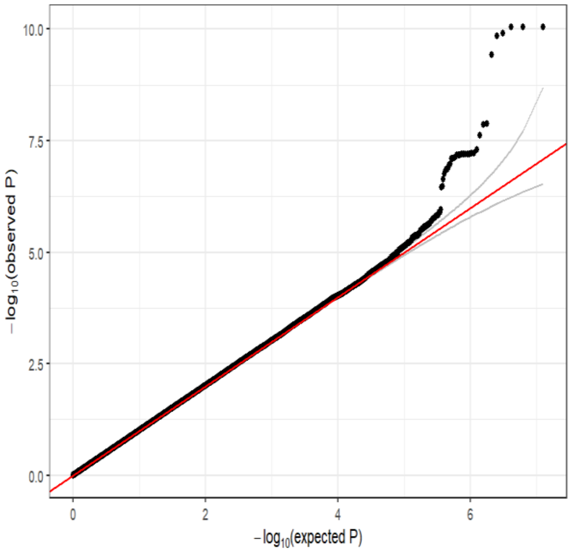

**(c)** AFA
λ=0.99

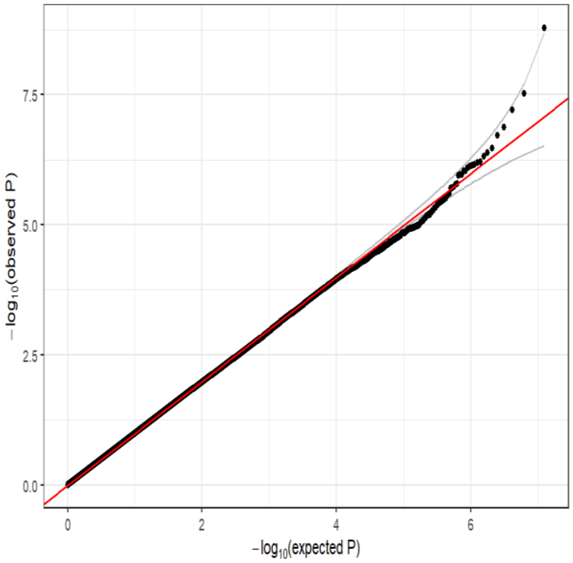

**(d)** EAS
λ=0.971

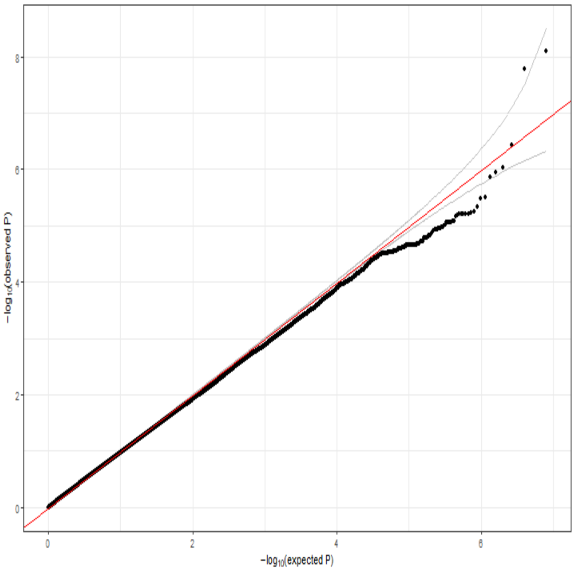

**Supplementary Figure 5.** Manhattan plot of cross-ancestry meta-analysis of *APOE* ε4-adjusted model (Age, sex, PCs and *APOE* ε4) for genome-wide association with Alzheimer's disease.

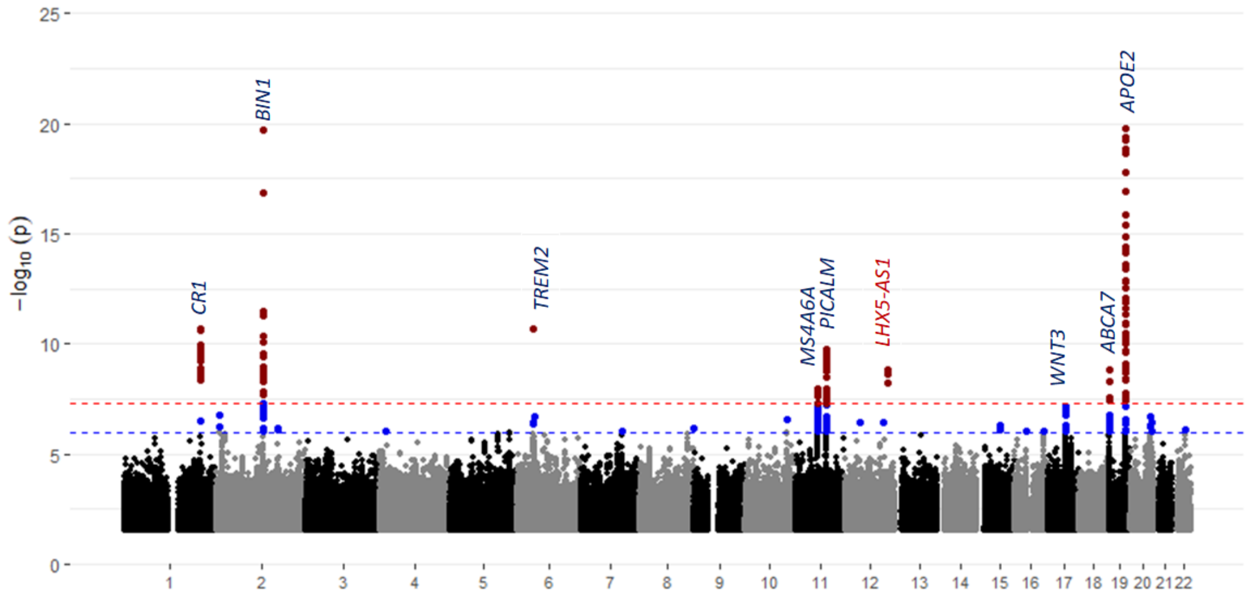

**Supplementary Figure 6.** Quantile-quantile (Q-Q) Plots for association results from the cross-ancestry genome-wide association analyses **(a)** with adjustment for age-at-onset (cases)/age-at-last exam (controls), sex, and PCs for population substructure (*APOE* region removed) and **(b)** with additional adjustment for dosage of the *APOE* ε4 allele.

**(a)** λ=0.941

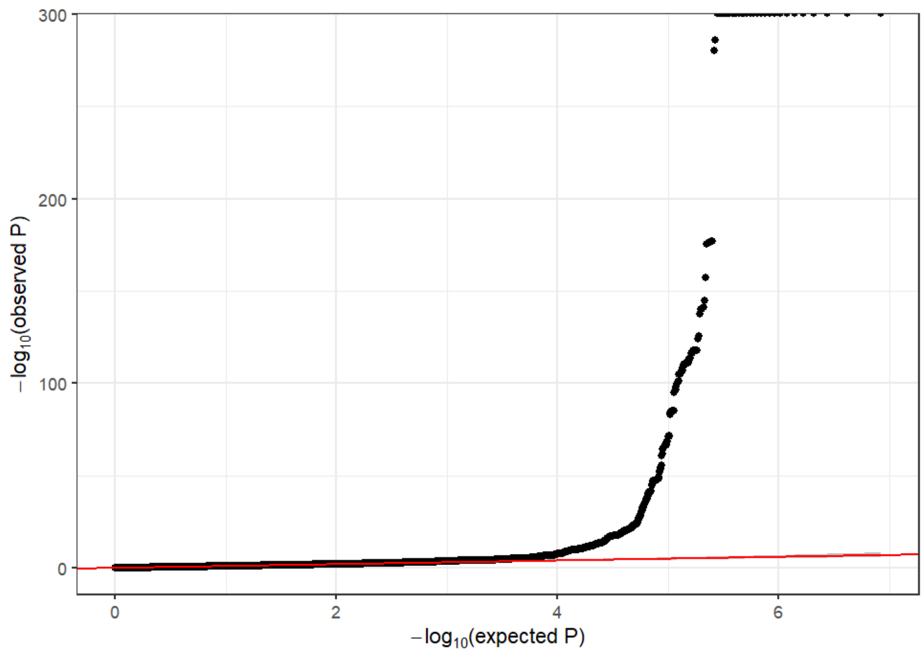

**(b)** λ=0.950

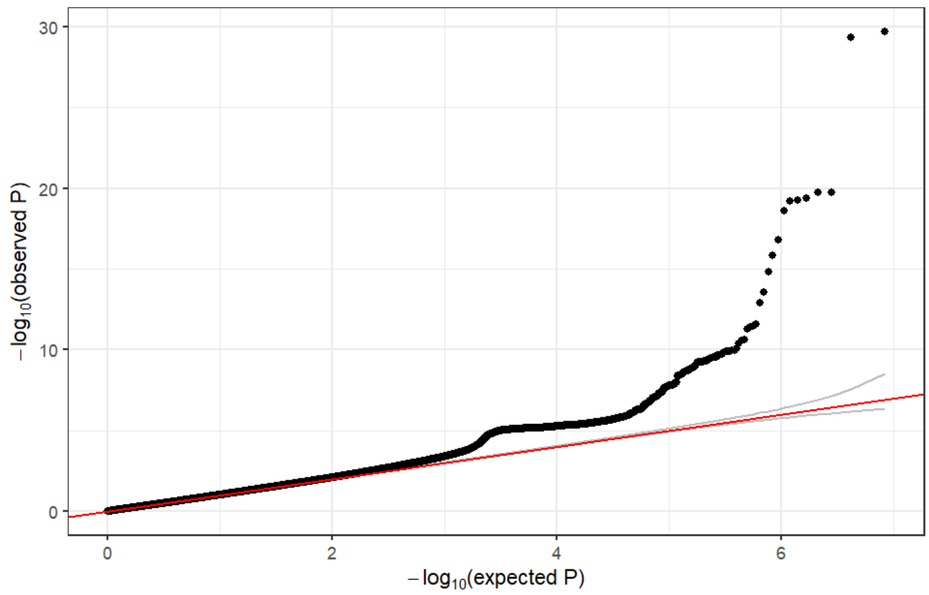

**Supplementary Figure 7.** Regional association plots for **(a)** cross-ancestry and **(b)** within-ancestry associations at the locus *BIN1*, as well as **(c)** Forest plots of the index variant rs6733839.

**(a)**

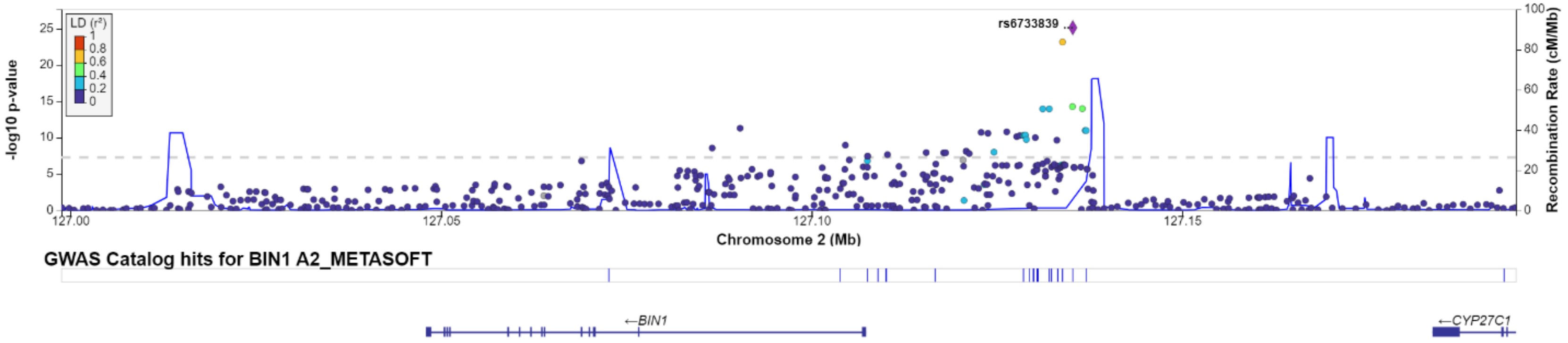

(b)

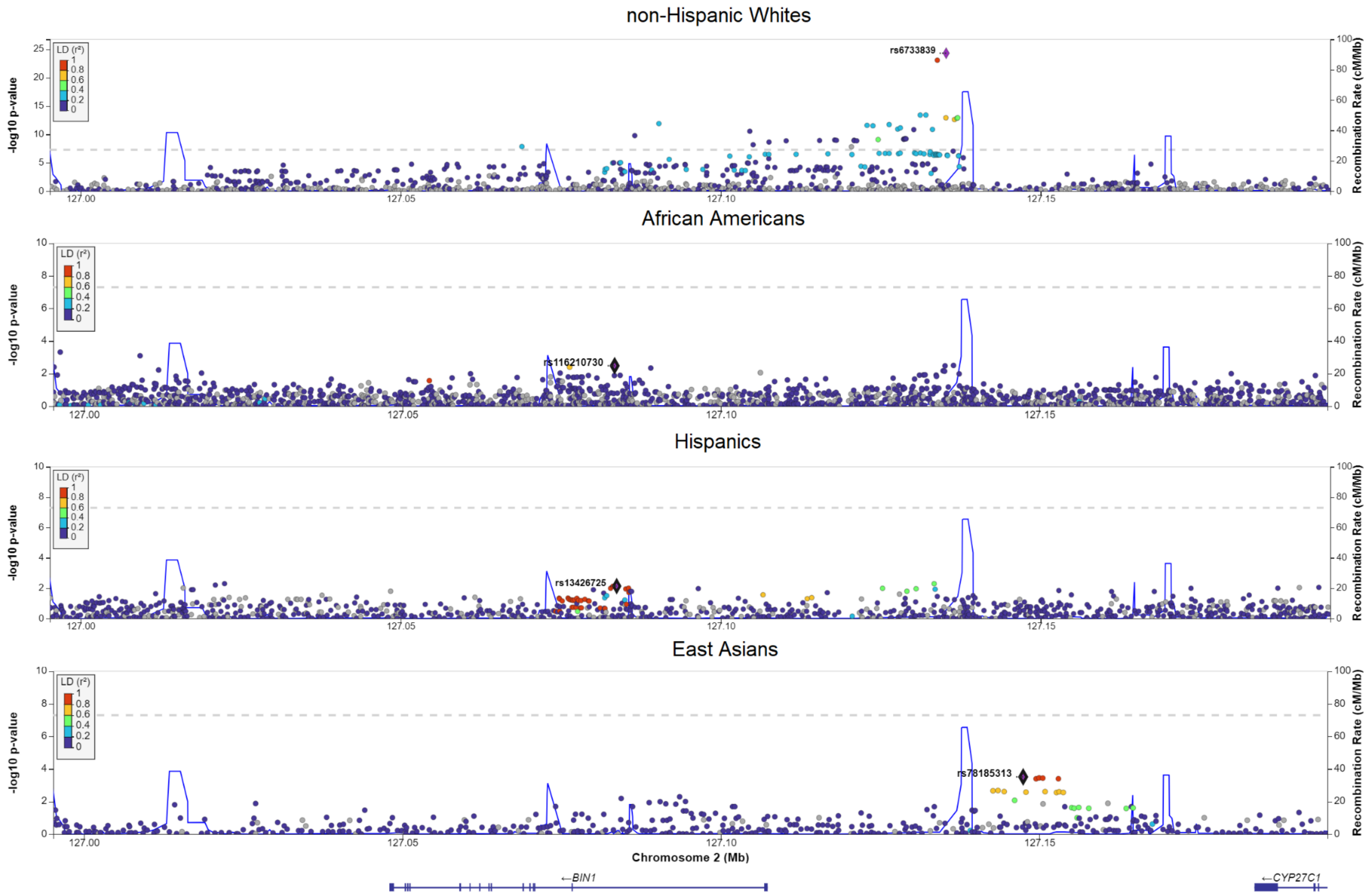
non-Hispanic Whites
rs6733839
African Americans
rs116210730
Hispanics
rs13426725
East Asians
rs78185313
LD (r²)
-log10 p-value
Recombination Rate (cM/Mb)
127.00
127.05
127.10
127.15
Chromosome 2 (Mb)
←BIN1
←CYP27C1

**(c)**

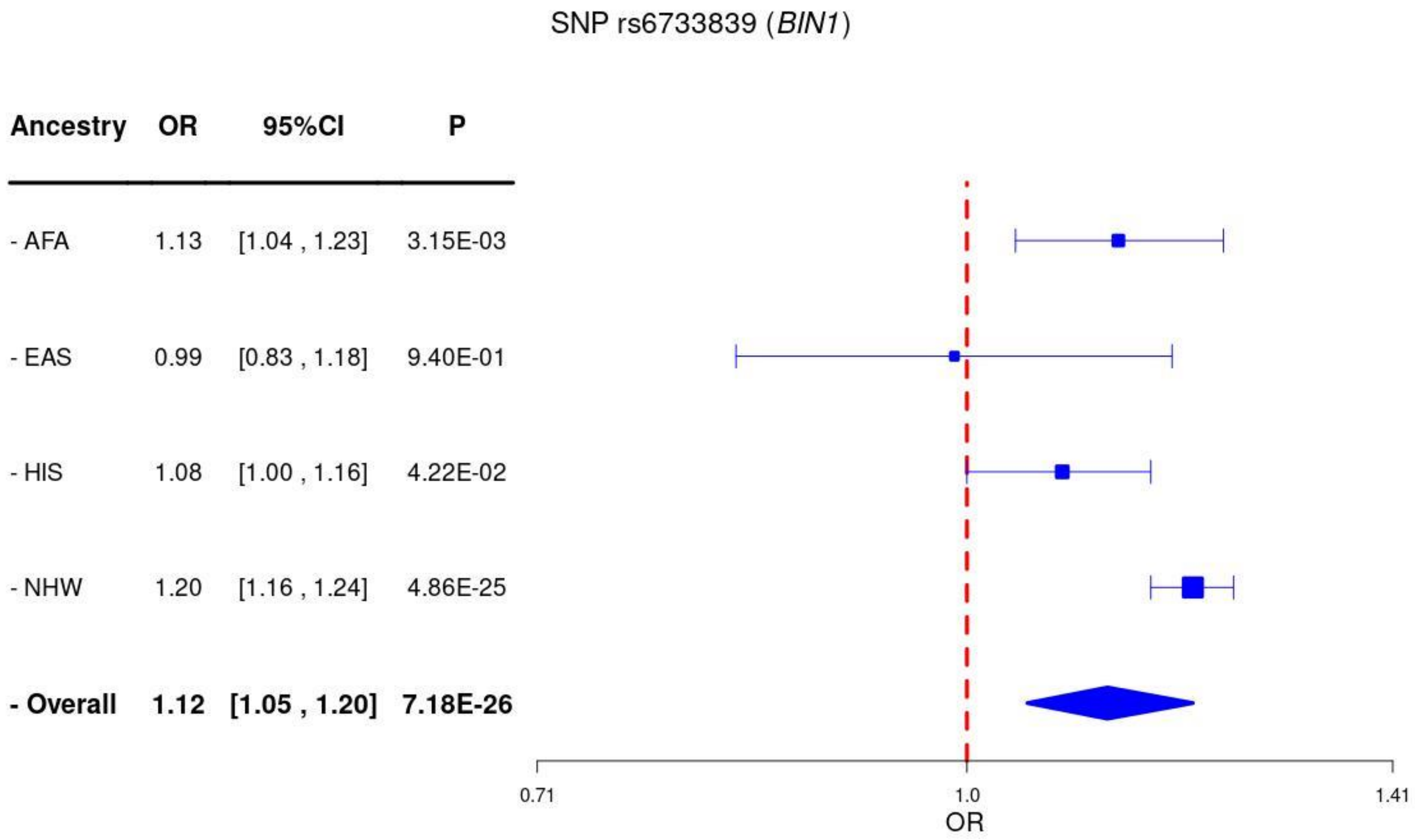
SNP rs6733839 (BIN1)
Ancestry
OR
95%CI
P
- AFA
1.13
[1.04 , 1.23]
3.15E-03
- EAS
0.99
[0.83 , 1.18]
9.40E-01
- HIS
1.08
[1.00 , 1.16]
4.22E-02
- NHW
1.20
[1.16 , 1.24]
4.86E-25
- Overall
1.12
[1.05 , 1.20]
7.18E-26
0.71
1.0
1.41
OR

**Supplementary Figure 8.** Regional association plots for **(a)** cross-ancestry and **(b)** within-ancestry associations at the locus *CD2AP*, , as well as **(c)** Forest plots of the index variant rs1385742.

**(a)**

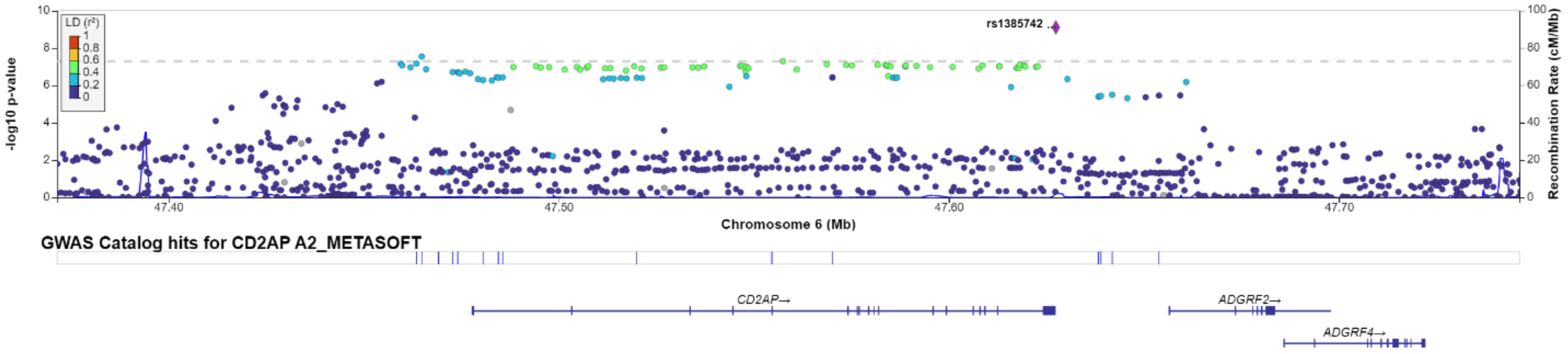

**(b)**

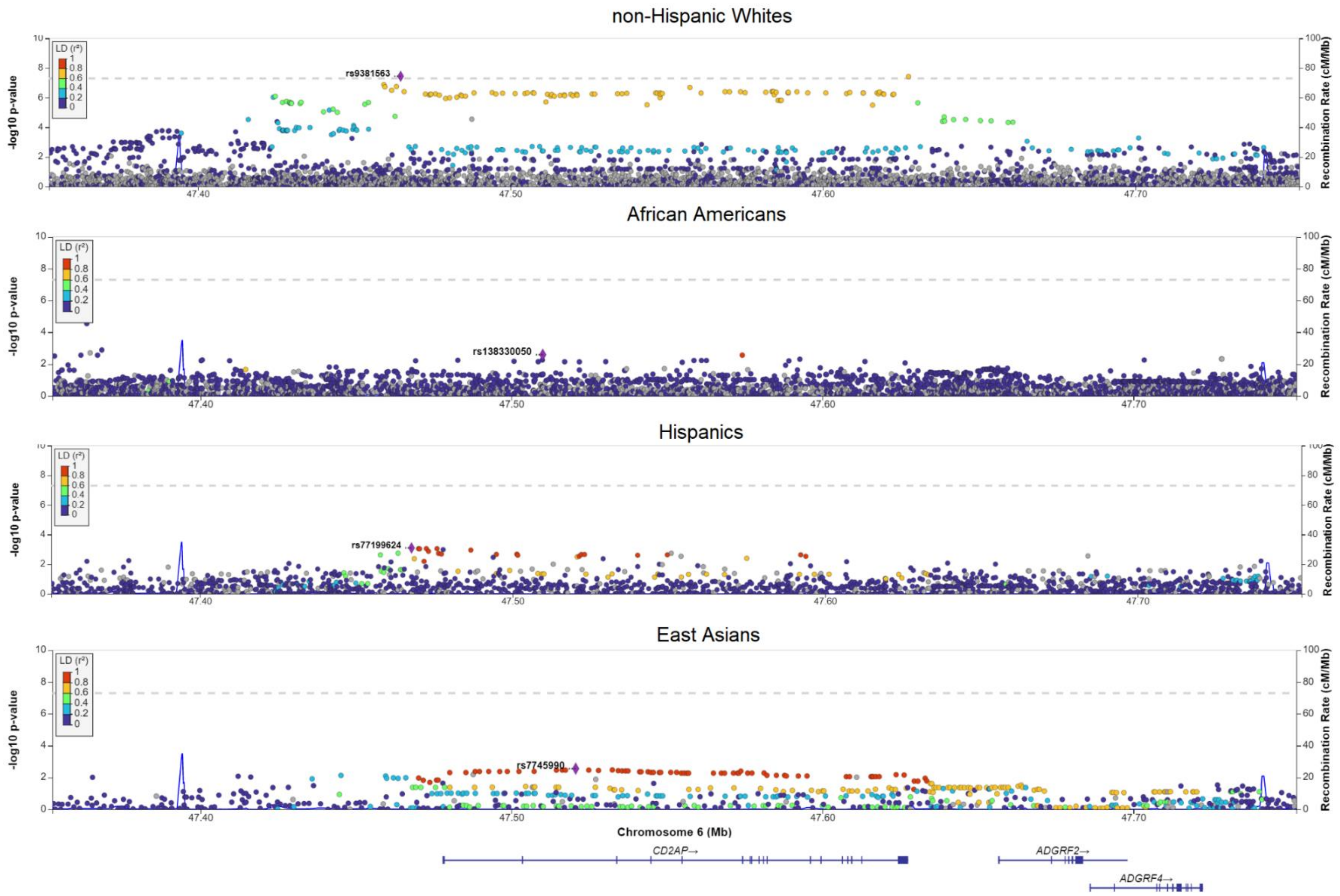

non-Hispanic Whites
rs9381563
African Americans
rs138330050
Hispanics
rs77199624
East Asians
rs7745990
LD (r²)
-log10 p-value
Recombination Rate (cM/Mb)
47.40
47.50
47.60
47.70
Chromosome 6 (Mb)
CD2AP→
ADGRF2→
ADGRF4→

(c)

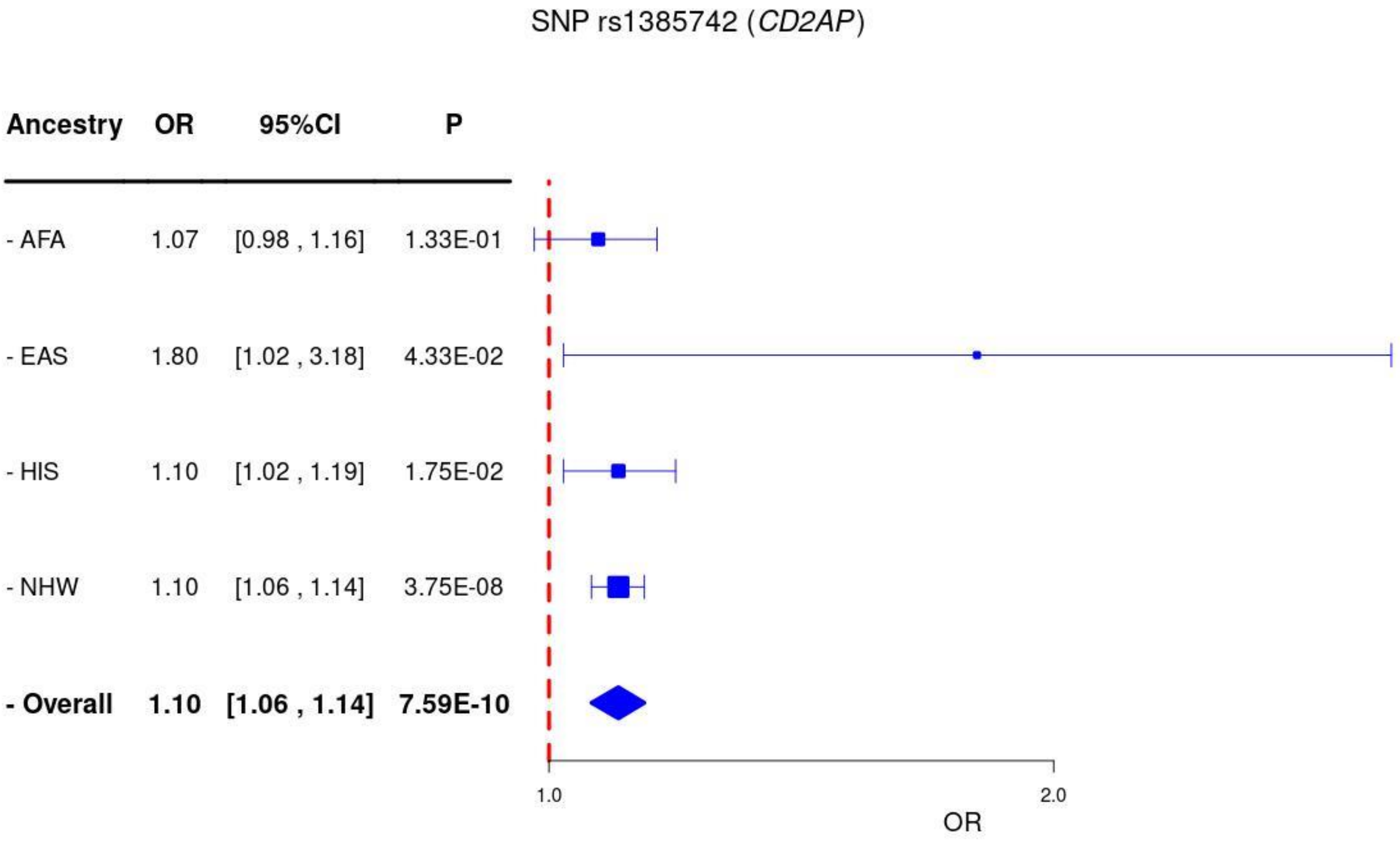
SNP rs1385742 (CD2AP)
Ancestry
OR
95%CI
P
- AFA
1.07
[0.98 , 1.16]
1.33E-01
- EAS
1.80
[1.02 , 3.18]
4.33E-02
- HIS
1.10
[1.02 , 1.19]
1.75E-02
- NHW
1.10
[1.06 , 1.14]
3.75E-08
- Overall
1.10
[1.06 , 1.14]
7.59E-10
1.0
2.0
OR

**Supplementary Figure 9.** Regional association plots for **(a)** cross-ancestry and **(b)** within-ancestry associations at the locus *PTK2B*, as well as **(c)** Forest plots of the index variant rs2741342.

**(a)**

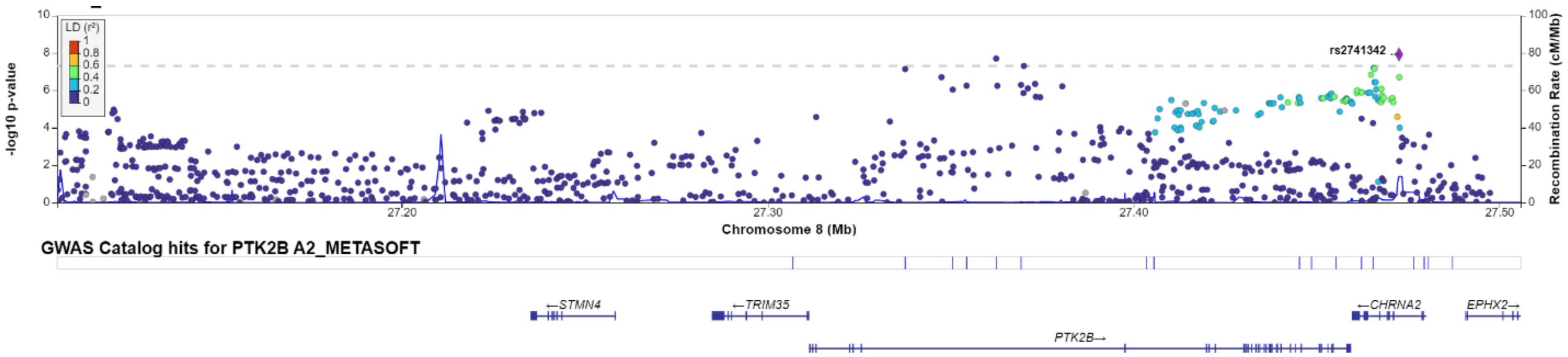

(b)

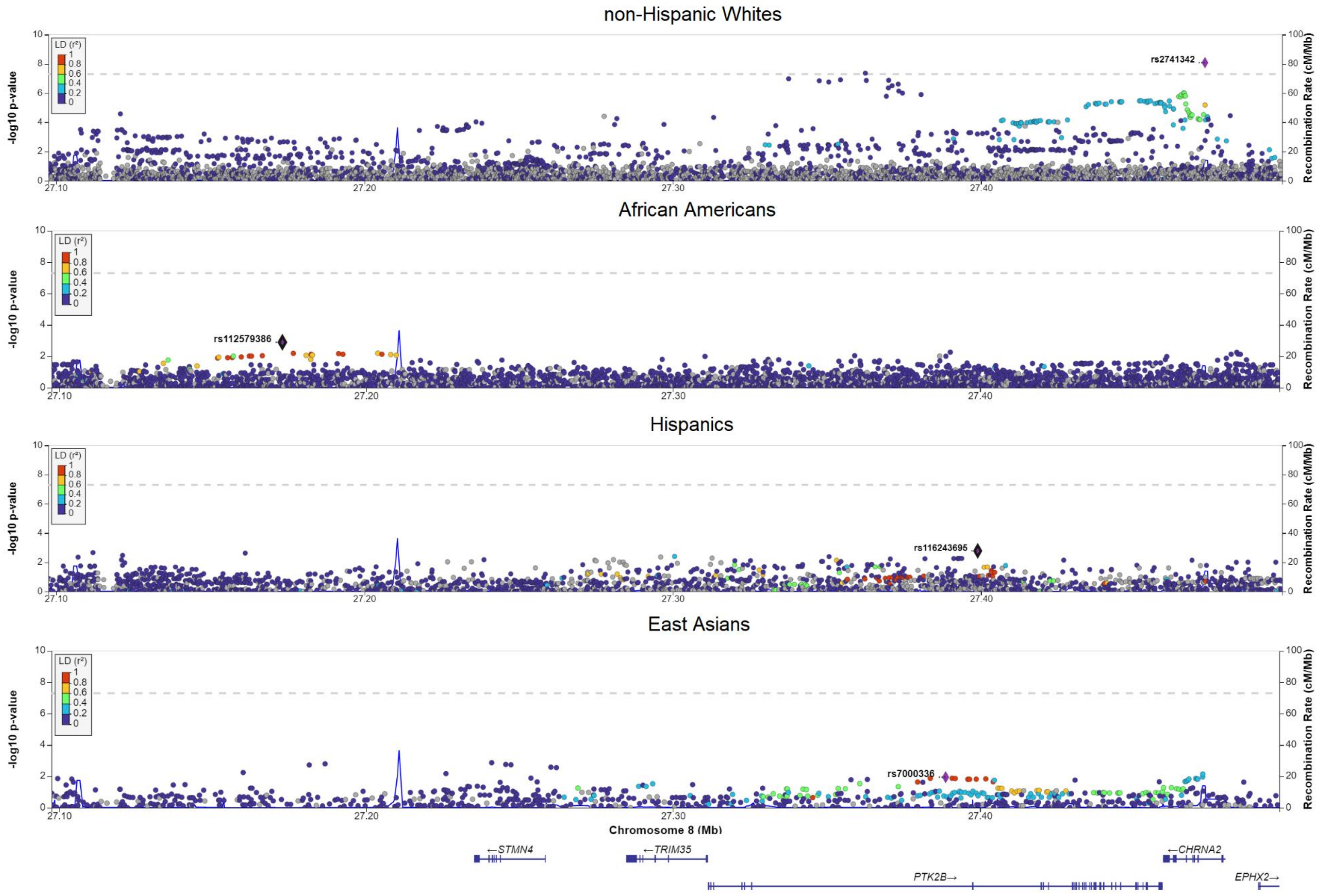

non-Hispanic Whites
African Americans
Hispanics
East Asians
LD (r²)
-log10 p-value
Recombination Rate (cM/Mb)
rs2741342
rs112579386
rs116243695
rs7000336
Chromosome 8 (Mb)
←STMN4
←TRIM35
←CHRNA2
PTK2B→
EPHX2→

**(c)**

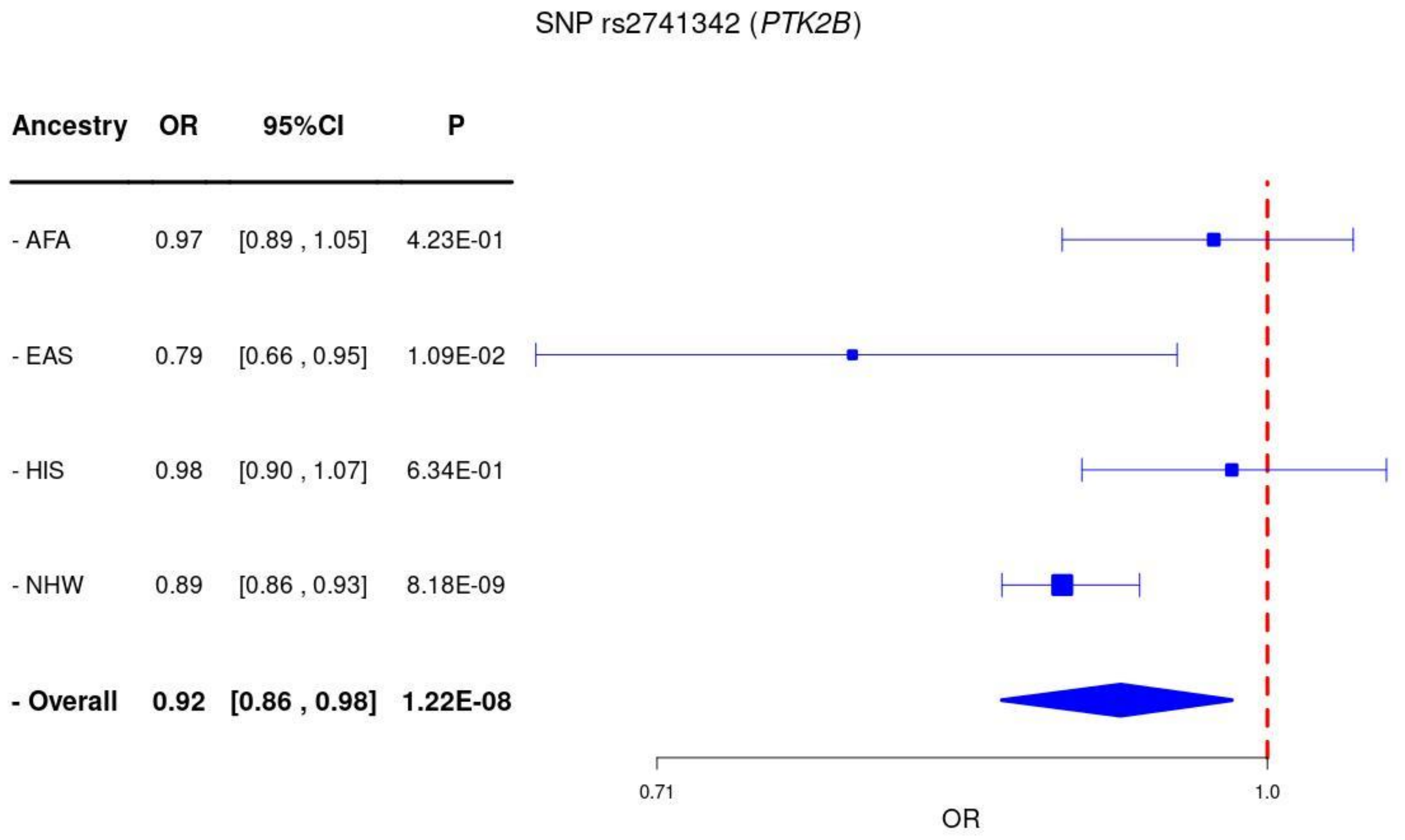
SNP rs2741342 (*PTK2B*)
Ancestry OR 95%CI P
- AFA 0.97 [0.89 , 1.05] 4.23E-01
- EAS 0.79 [0.66 , 0.95] 1.09E-02
- HIS 0.98 [0.90 , 1.07] 6.34E-01
- NHW 0.89 [0.86 , 0.93] 8.18E-09
- Overall 0.92 [0.86 , 0.98] 1.22E-08
0.71
1.0
OR

**Supplementary Figure 10.** Regional association plots for **(a)** cross-ancestry and **(b)** within-ancestry associations at the locus *CLU*, as well as **(c)** Forest plots of the index variant rs1532278.

**(a)**

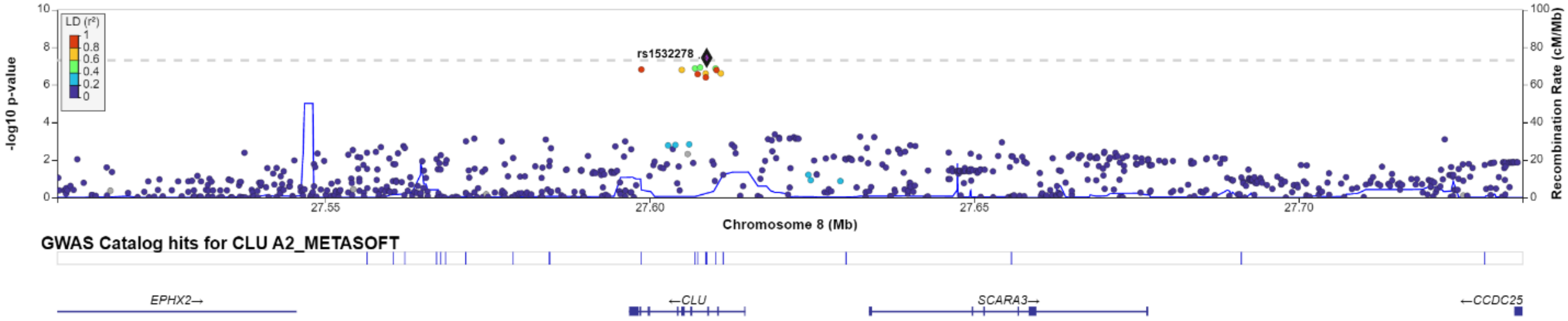

(b)

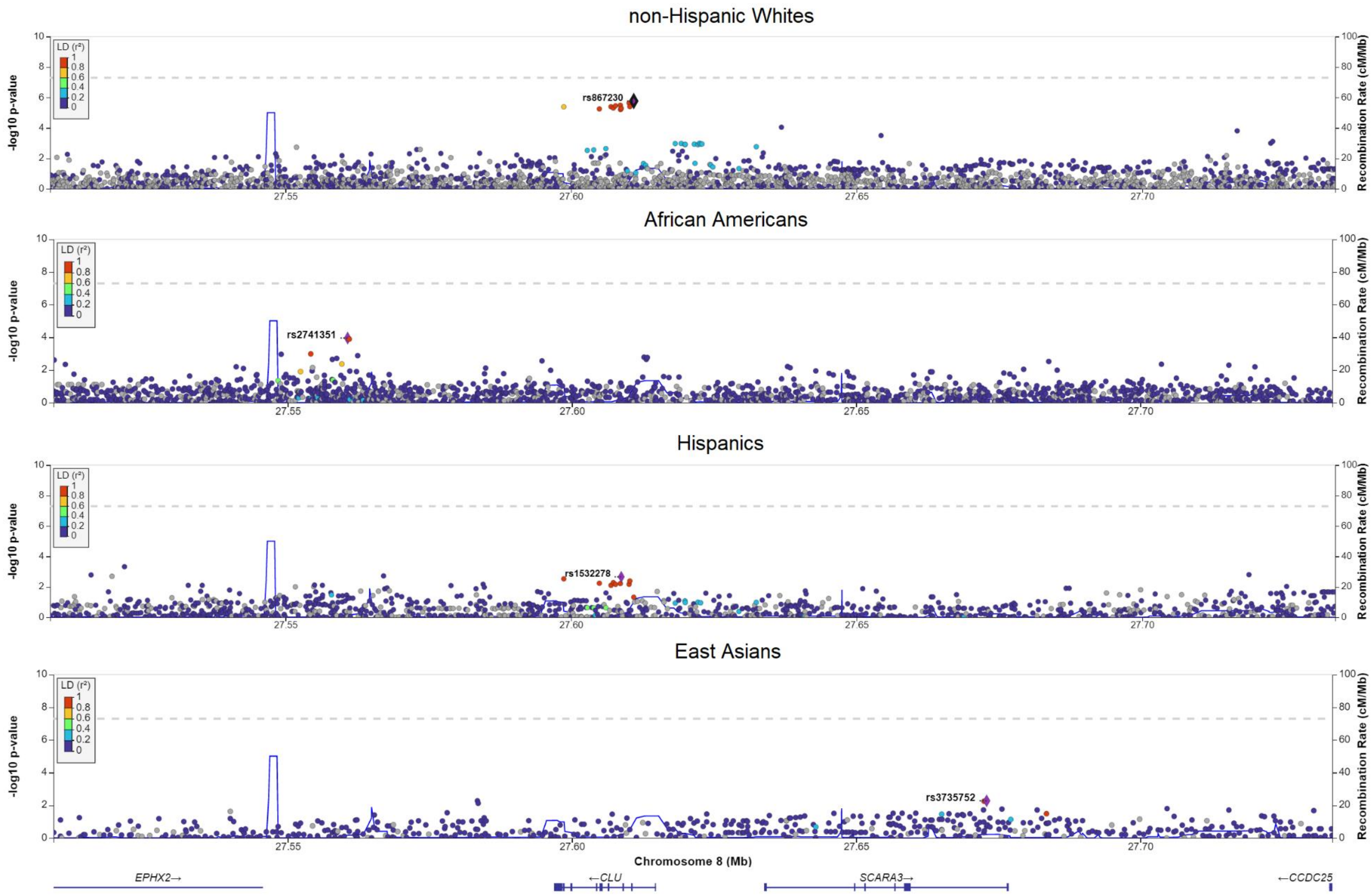
non-Hispanic Whites
African Americans
Hispanics
East Asians
LD (r²)
-log10 p-value
Recombination Rate (cM/Mb)
rs867230
rs2741351
rs1532278
rs3735752
27.55
27.60
27.65
27.70
Chromosome 8 (Mb)
EPHX2→
←CLU
SCARA3→
←CCDC25

(c)

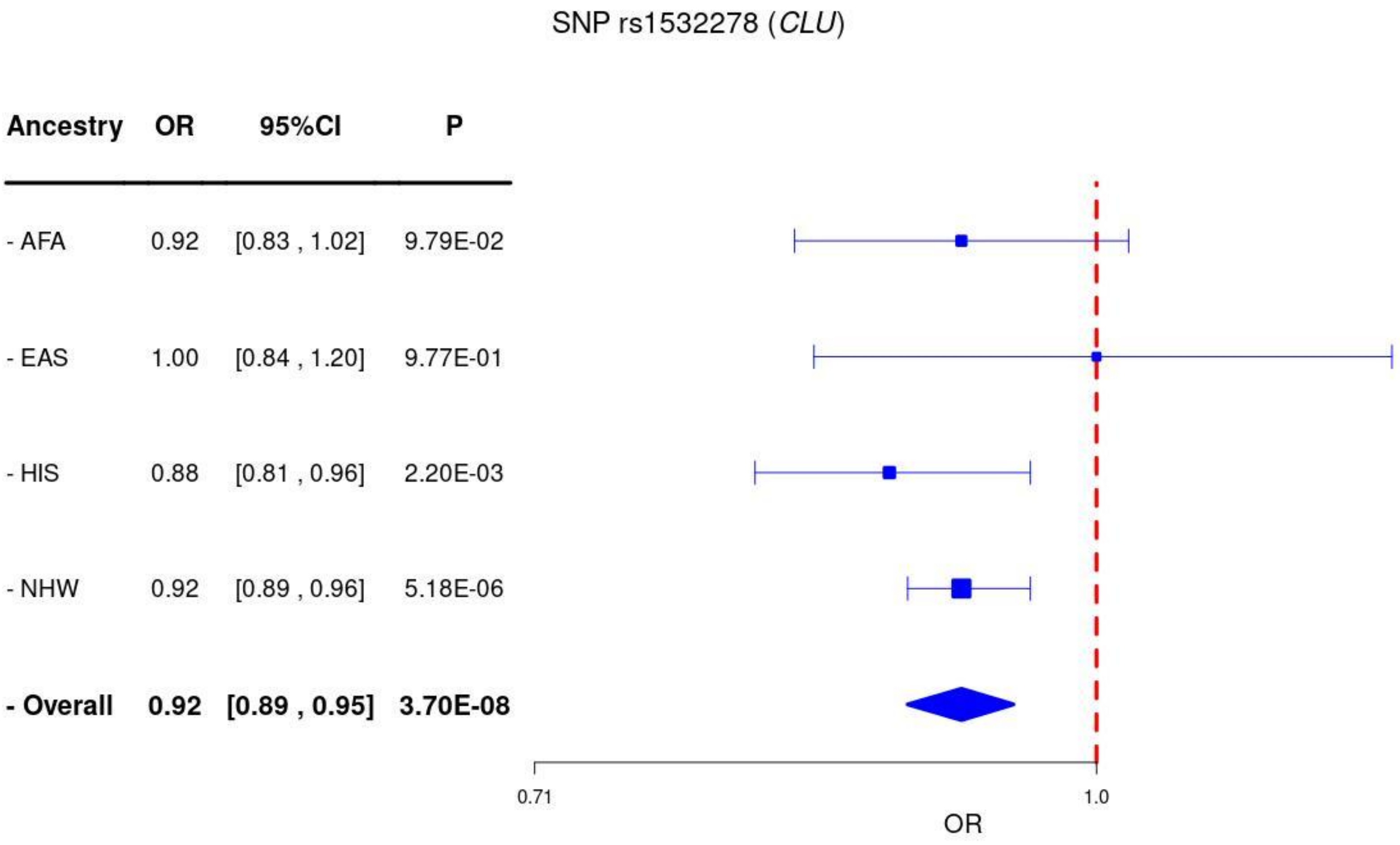

SNP rs1532278 (CLU)
Ancestry OR 95%CI P
- AFA 0.92 [0.83 , 1.02] 9.79E-02
- EAS 1.00 [0.84 , 1.20] 9.77E-01
- HIS 0.88 [0.81 , 0.96] 2.20E-03
- NHW 0.92 [0.89 , 0.96] 5.18E-06
- Overall 0.92 [0.89 , 0.95] 3.70E-08
0.71
1.0
OR

**Supplementary Figure 11.** Regional association plots for **(a)** cross-ancestry and **(b)** within-ancestry associations at the locus *SHARPIN*, as well as **(c)** Forest plots of the index variant rs34173062.

**(a)**

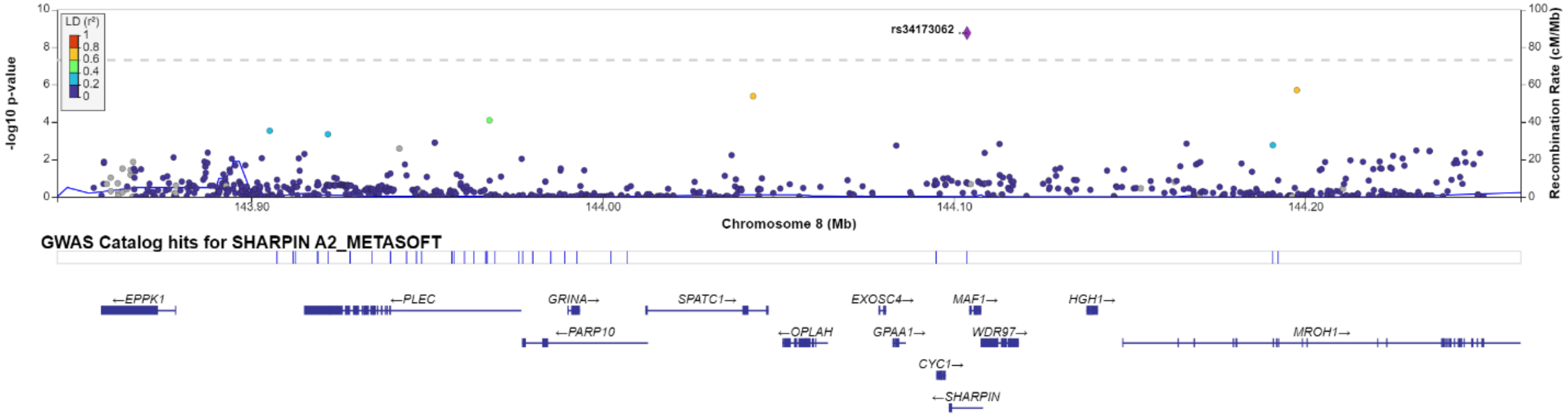

(b)

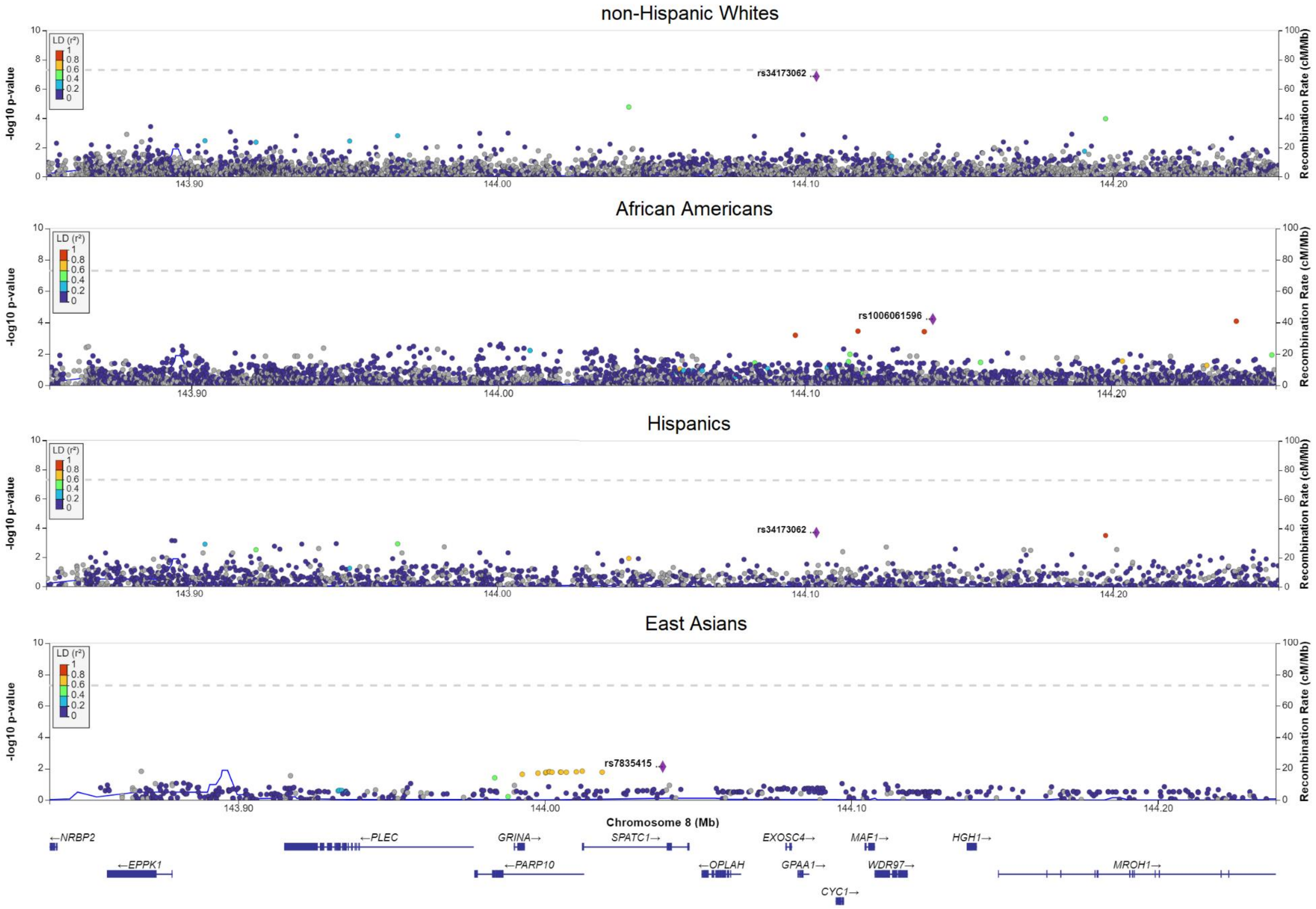
non-Hispanic Whites
African Americans
Hispanics
East Asians
LD (r²)
-log10 p-value
Recombination Rate (cM/Mb)
rs34173062
rs1006061596
rs7835415
143.90
144.00
144.10
144.20
Chromosome 8 (Mb)
←NRBP2
←EPPK1
←PLEC
GRINA→
←PARP10
SPATC1→
←OPLAH
EXOSC4→
GPAA1→
CYC1→
MAF1→
WDR97→
HGH1→
MROH1→

(c)

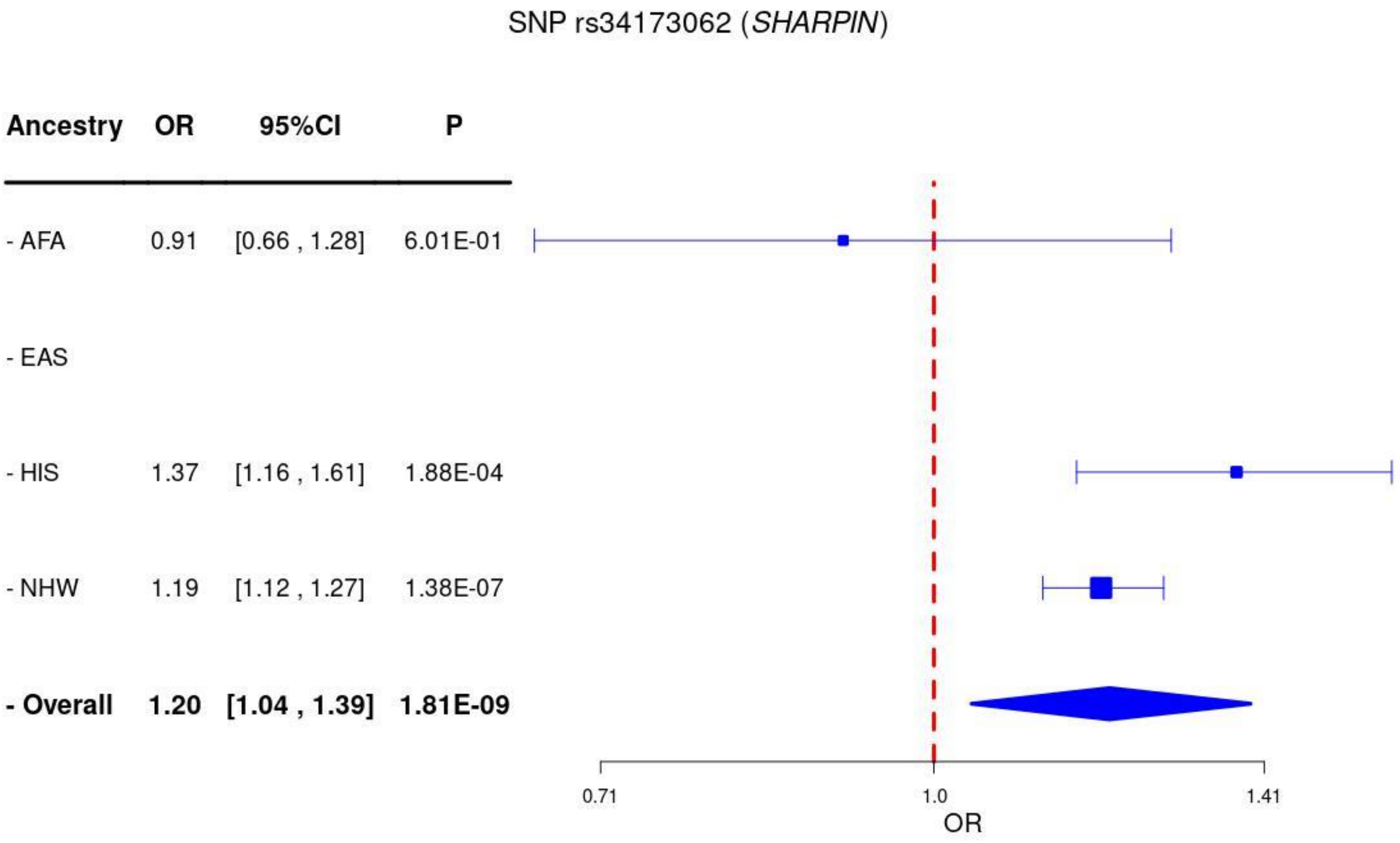
SNP rs34173062 (SHARPIN)
Ancestry
OR
95%CI
P
- AFA
0.91
[0.66 , 1.28]
6.01E-01
- EAS
- HIS
1.37
[1.16 , 1.61]
1.88E-04
- NHW
1.19
[1.12 , 1.27]
1.38E-07
- Overall
1.20
[1.04 , 1.39]
1.81E-09
0.71
1.0
1.41
OR

**Supplementary Figure 12.** Regional association plots for **(a)** cross-ancestry and **(b)** within-ancestry associations at the locus *MS4A6A*, as well as **(c)** Forest plots of the index variant rs1582763.

**(a)**

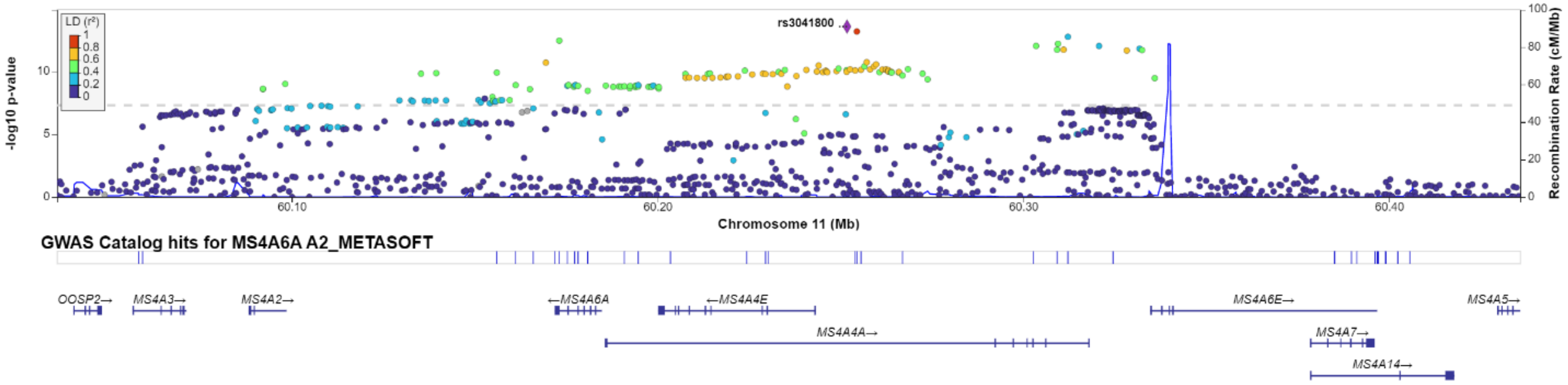

**(b)**

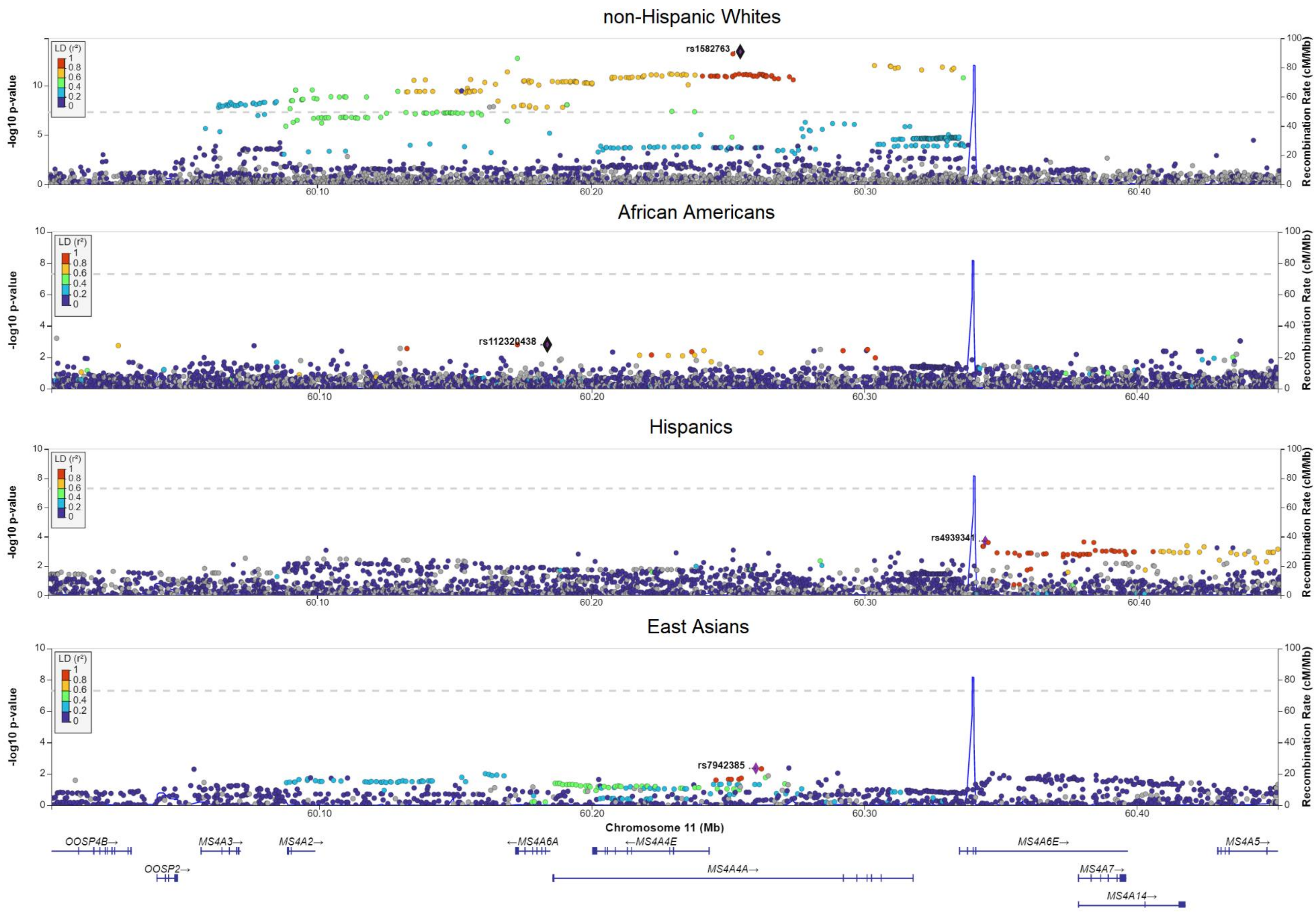

non-Hispanic Whites
African Americans
Hispanics
East Asians
-log10 p-value
Recombination Rate (cM/Mb)
LD (r²)
rs1582763
rs112320438
rs4939341
rs7942385
60.10
60.20
60.30
60.40
Chromosome 11 (Mb)
OOSP4B→
MS4A3→
MS4A2→
←MS4A6A
←MS4A4E
MS4A6E→
MS4A5→
OOSP2→
MS4A4A→
MS4A7→
MS4A14→

**(c)**

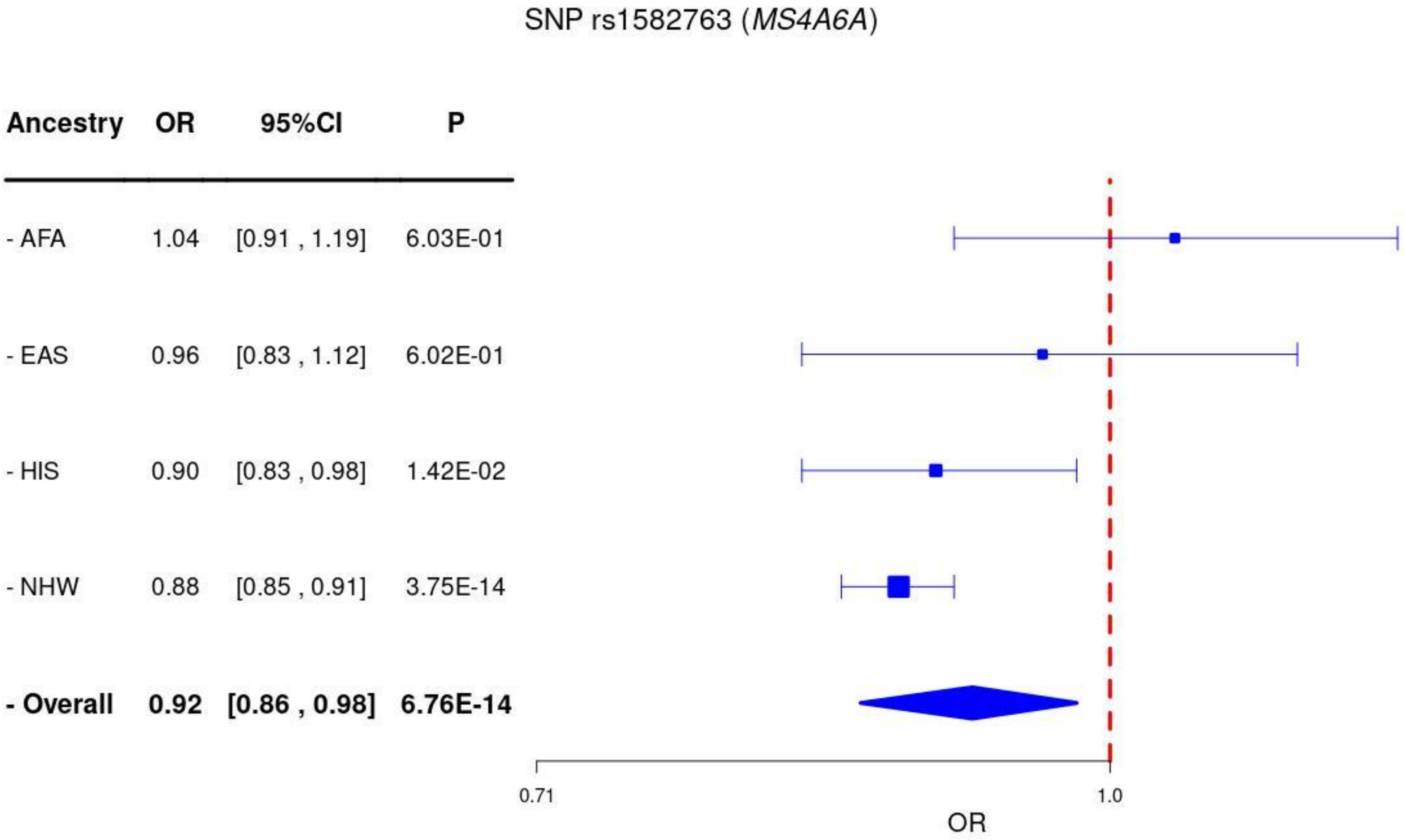
SNP rs1582763 (*MS4A6A*)
Ancestry
OR
95%CI
P
- AFA
1.04
[0.91 , 1.19]
6.03E-01
- EAS
0.96
[0.83 , 1.12]
6.02E-01
- HIS
0.90
[0.83 , 0.98]
1.42E-02
- NHW
0.88
[0.85 , 0.91]
3.75E-14
- Overall
0.92
[0.86 , 0.98]
6.76E-14
0.71
1.0
OR

**Supplementary Figure 13.** Regional association plots for **(a)** cross-ancestry and **(b)** within-ancestry associations at the locus *PICALM*, as well as **(c)** Forest plots of the index variant rs1898895.

**(a)**

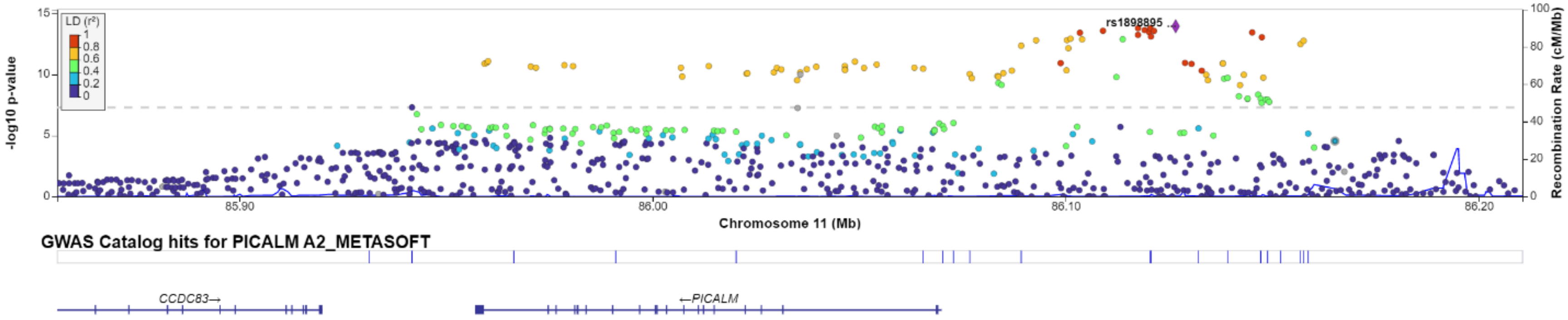

(b)

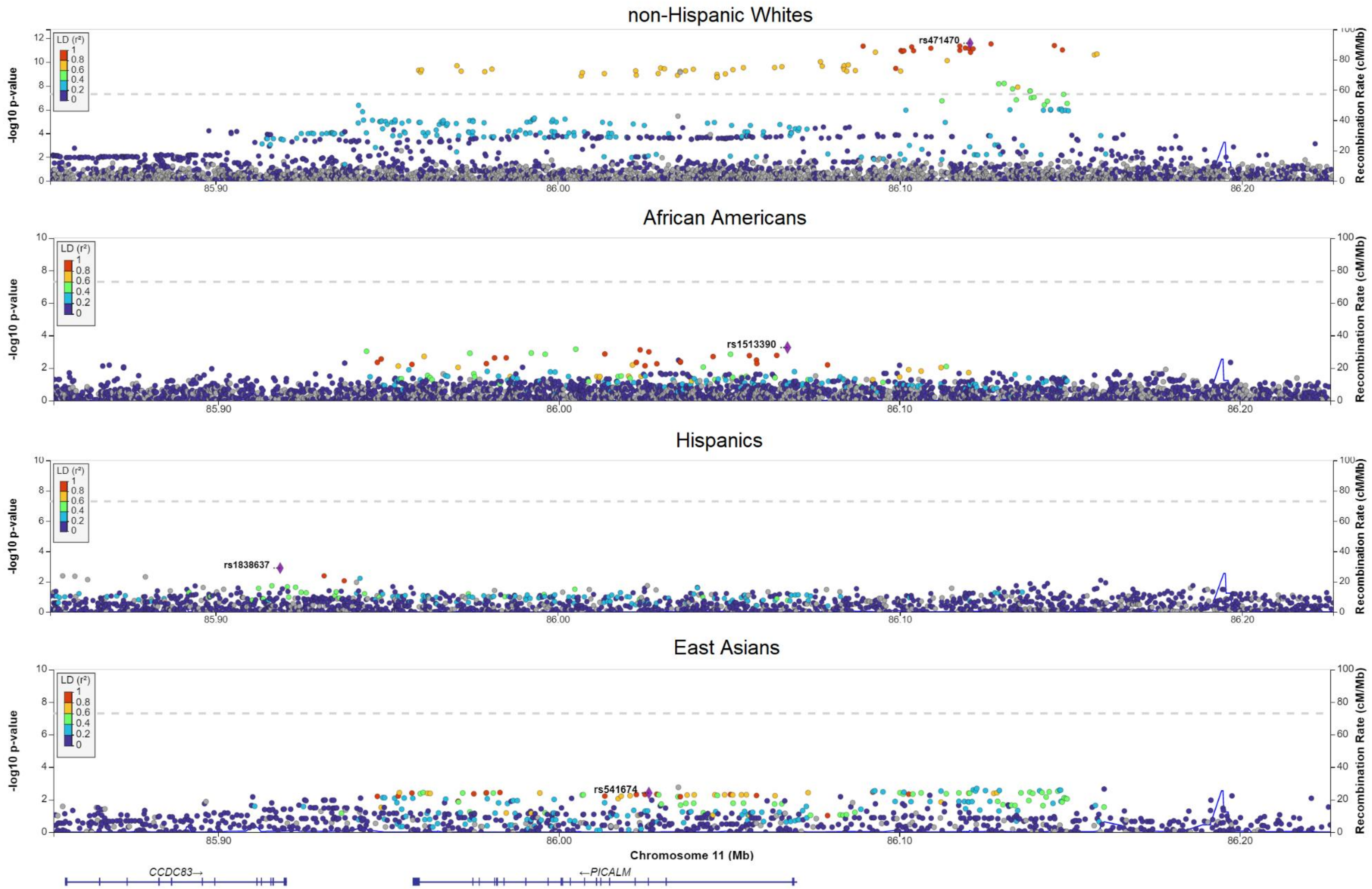
non-Hispanic Whites
rs471470
African Americans
rs1513390
Hispanics
rs1838637
East Asians
rs541674
LD (r²)
-log10 p-value
Recombination Rate (cM/Mb)
85.90
86.00
86.10
86.20
Chromosome 11 (Mb)
CCDC83→
←PICALM

(c)

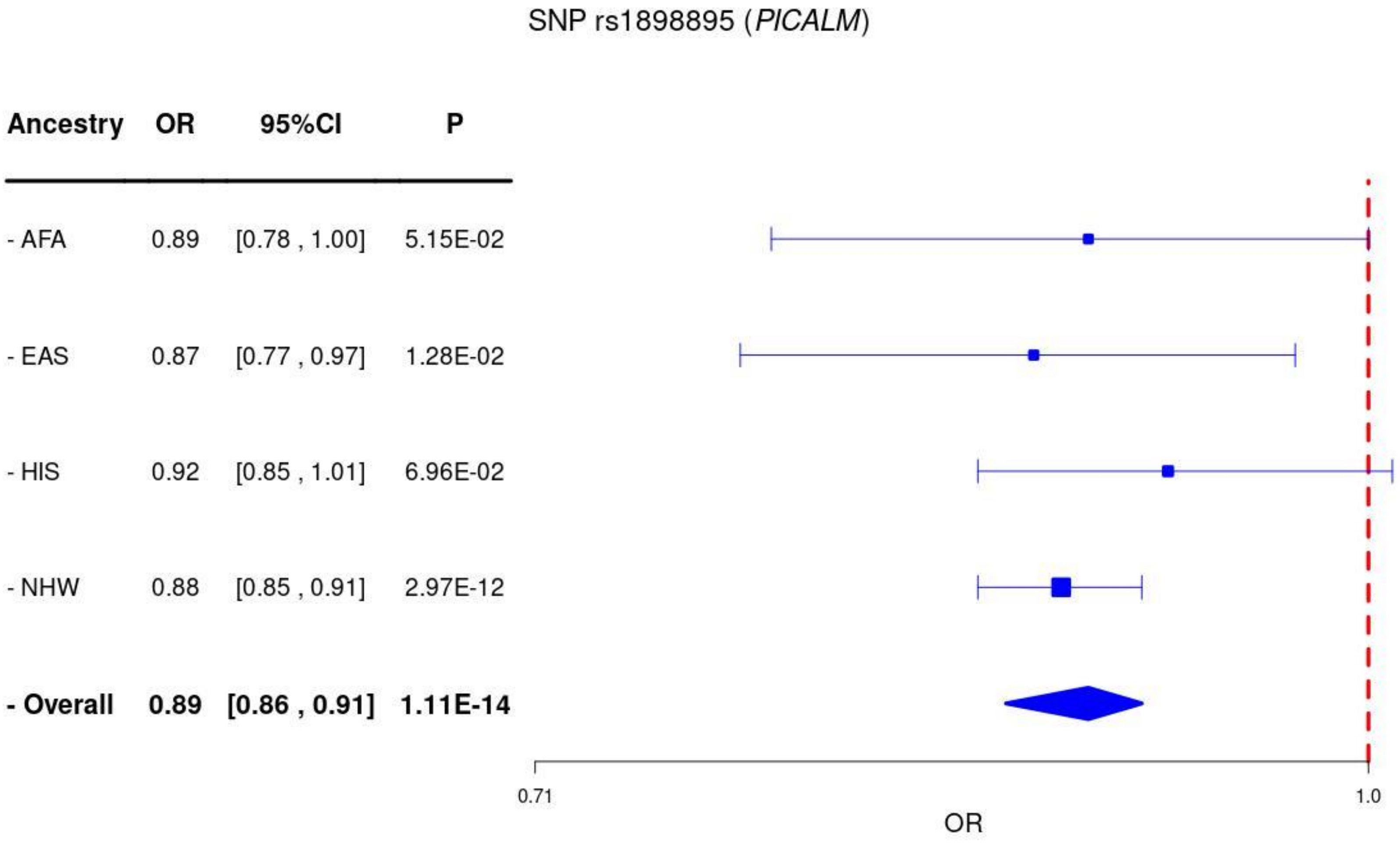

SNP rs1898895 (*PICALM*)
Ancestry
OR
95%CI
P
- AFA
0.89
[0.78 , 1.00]
5.15E-02
- EAS
0.87
[0.77 , 0.97]
1.28E-02
- HIS
0.92
[0.85 , 1.01]
6.96E-02
- NHW
0.88
[0.85 , 0.91]
2.97E-12
- Overall
0.89
[0.86 , 0.91]
1.11E-14
0.71
1.0
OR

**Supplementary Figure 14.** Regional association plots for **(a)** cross-ancestry and **(b)** within-ancestry associations at the locus *ABCA7*, as well as **(c)** Forest plots of the index variant rs12151021.

**(a)**

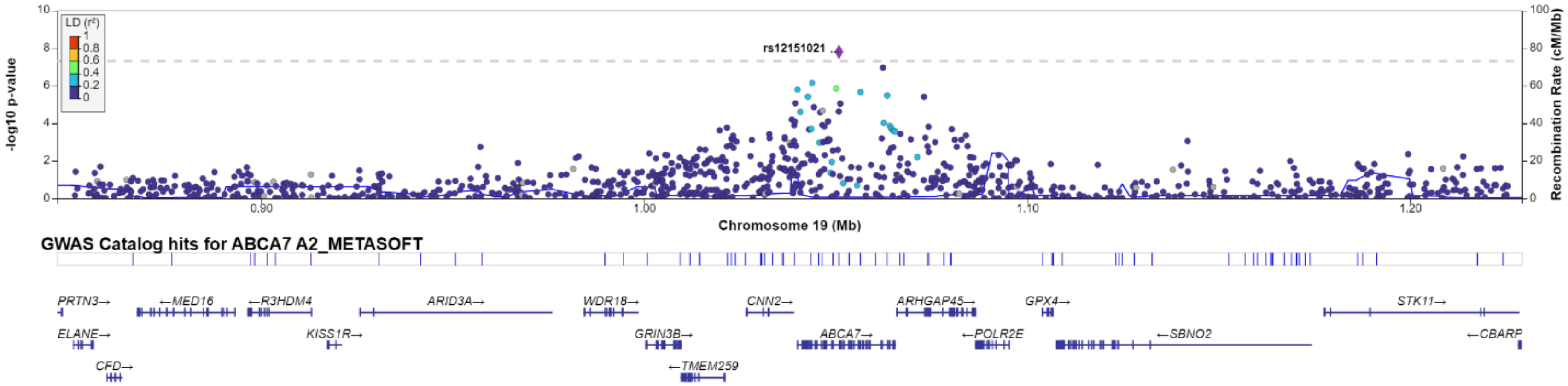

**(b)**

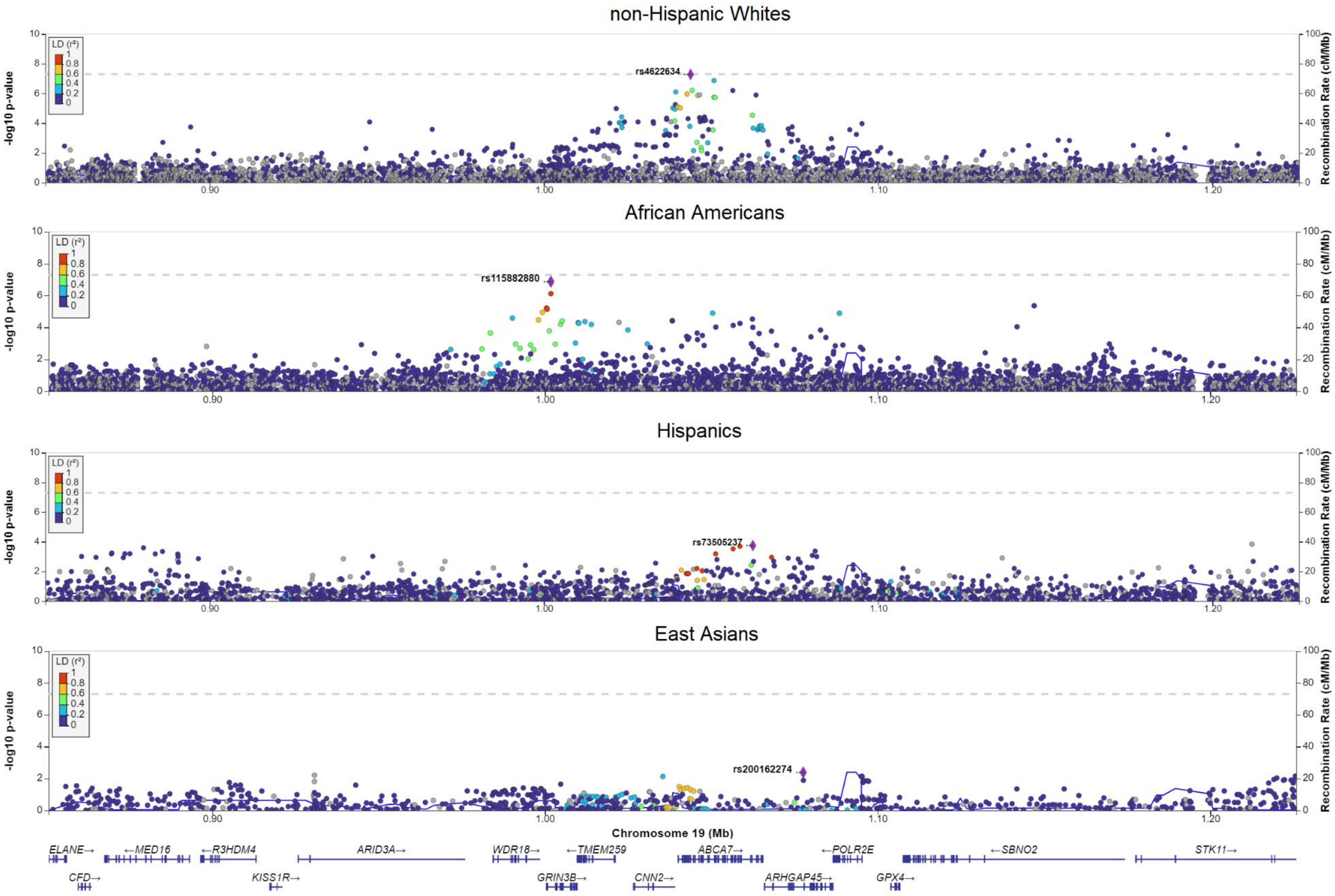

non-Hispanic Whites
African Americans
Hispanics
East Asians
-log10 p-value
Recombination Rate (cM/Mb)
LD (r²)
rs4622634
rs115882880
rs73505237
rs200162274
Chromosome 19 (Mb)
0.90
1.00
1.10
1.20
ELANE→
←MED16
←R3HDM4
ARID3A→
WDR18→
←TMEM259
ABCA7→
←POLR2E
←SBNO2
STK11→
CFD→
KISS1R→
GRIN3B→
CNN2→
ARHGAP45→
GPX4→

(c)

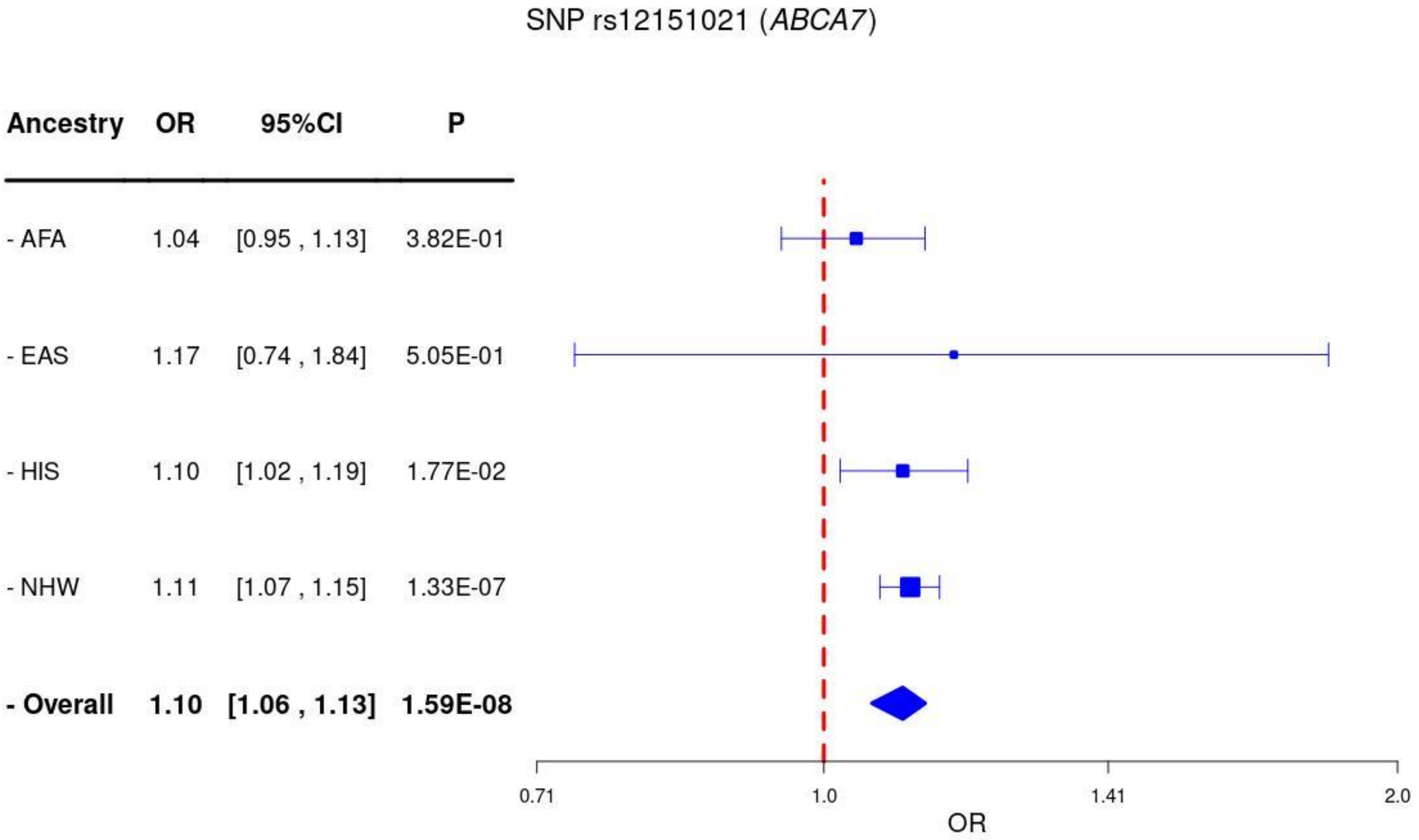
SNP rs12151021 (*ABCA7*)
Ancestry OR 95%CI P
- AFA 1.04 [0.95 , 1.13] 3.82E-01
- EAS 1.17 [0.74 , 1.84] 5.05E-01
- HIS 1.10 [1.02 , 1.19] 1.77E-02
- NHW 1.11 [1.07 , 1.15] 1.33E-07
- Overall 1.10 [1.06 , 1.13] 1.59E-08
0.71
1.0
1.41
2.0
OR

**Supplementary Figure 15.** Regional association plots for **(a)** cross-ancestry and **(b)** within-ancestry associations at the locus *APOE*.

**(a)**

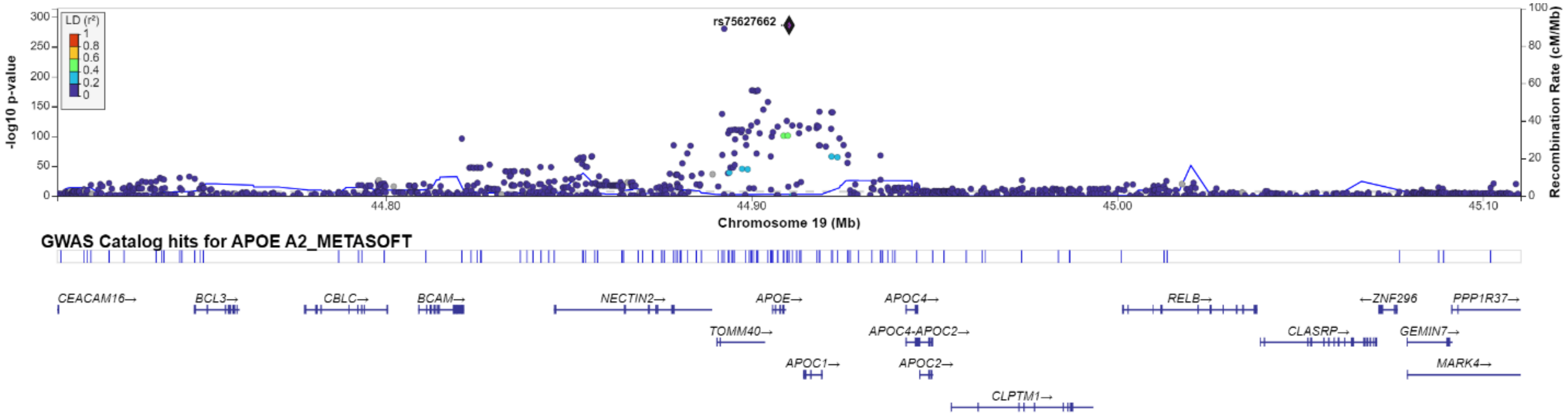

**(b)**

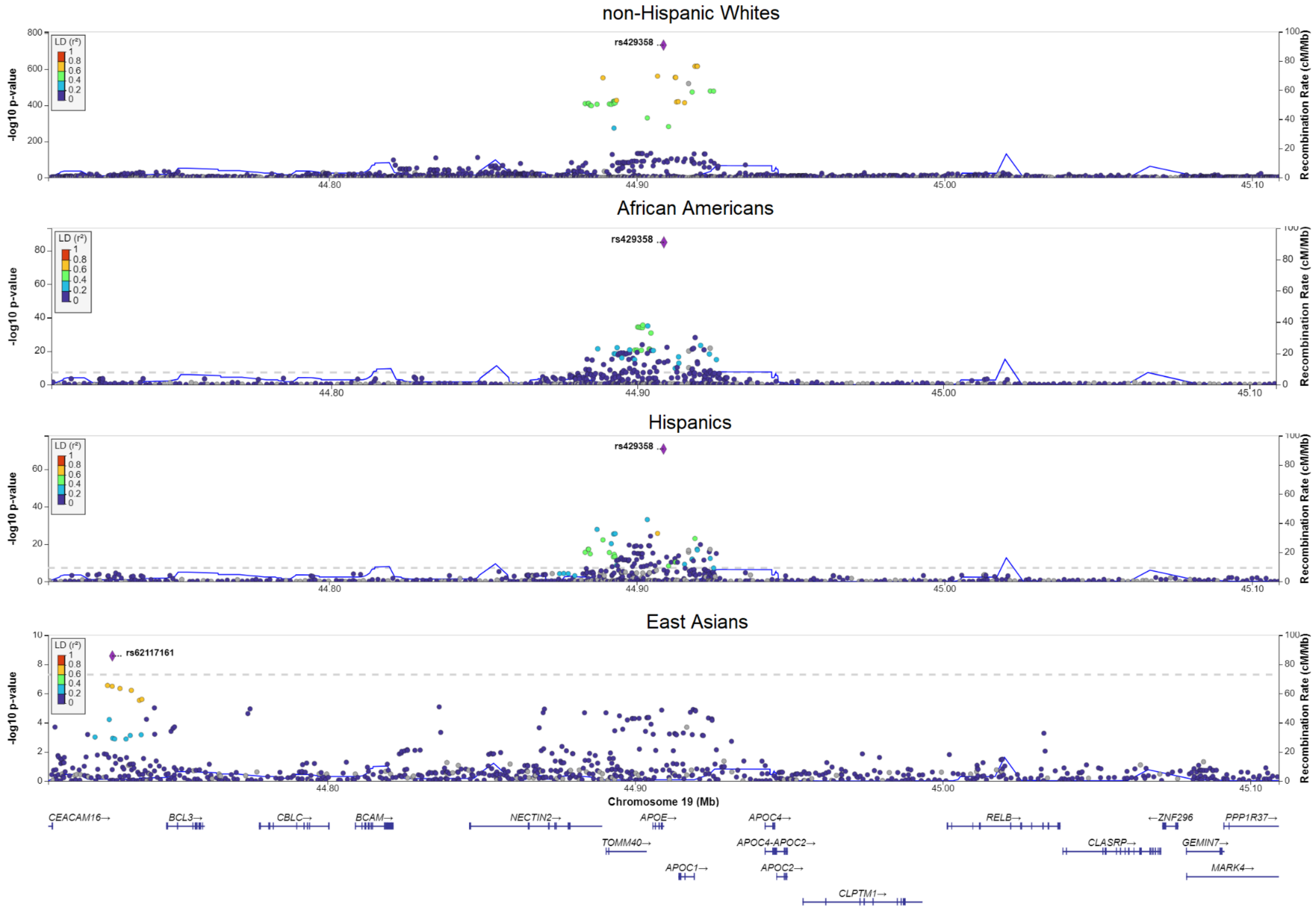
non-Hispanic Whites
African Americans
Hispanics
East Asians
rs429358
rs62117161
LD (r²)
-log10 p-value
Recombination Rate (cM/Mb)
Chromosome 19 (Mb)
44.80
44.90
45.00
45.10
CEACAM16→
BCL3→
CBLC→
BCAM→
NECTIN2→
APOE→
TOMM40→
APOC1→
APOC4→
APOC4-APOC2→
APOC2→
CLPTM1→
RELB→
CLASRP→
←ZNF296
GEMIN7→
MARK4→
PPP1R37→

**Supplementary Figure 16. *LRRC4C* locus annotation plot.** Shown is the region surrounding the LRRCRC locus and gene, with the location of the peak cross-ancestry association, rs12576934 ($P$=5.37×10$^{-9}$) shown with the vertical orange line. SNP rs12576934 is a near-significant splicing QTL (sQTL) ($P$=5.89×10$^{-4}$; nominal significance threshold $P$=6.99×10$^{-5}$) for LRRC4C in GTEx v8 brain frontal cortex and is located within ROADMAP-annotated neural progenitor cell (NPC) and neuronal enhancers.

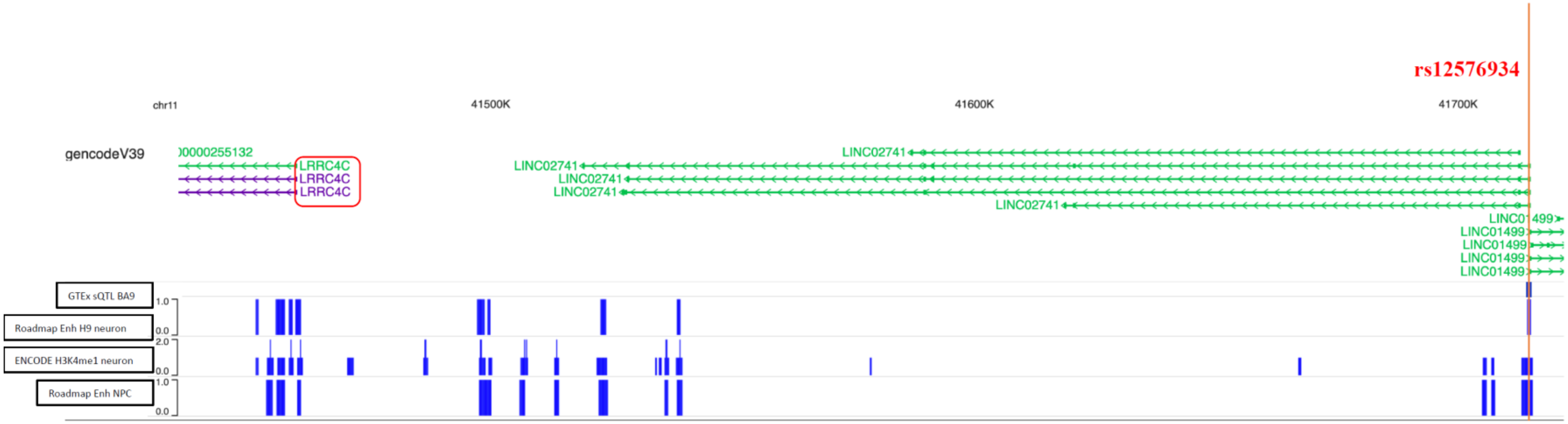

**Supplementary Figure 17.** Venn diagram with unique and common pathways cross different ancestries using genes flanking $P<10^{-6}$ within ancestry meta-analysis. **(a)** Pathways using genes from the *APOE*-unadjusted model. **(b)** Pathways using genes from the *APOE*-adjusted model.

**a.**

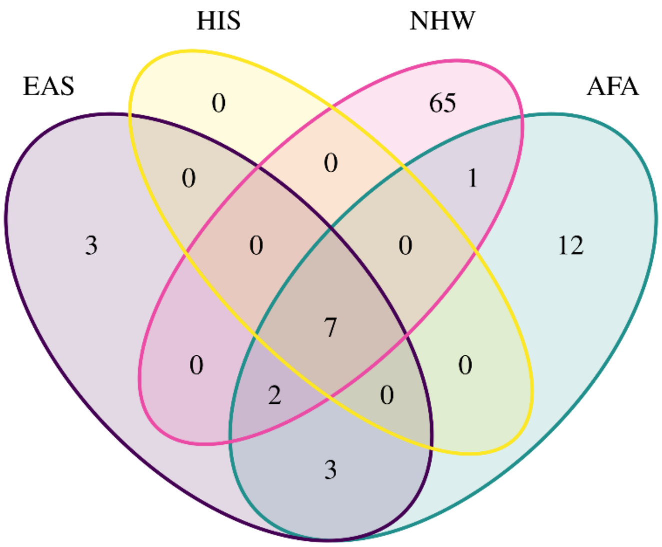

**b.**

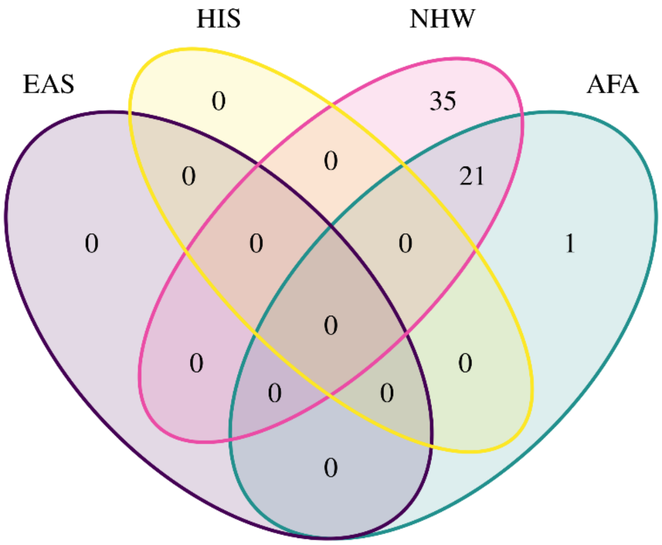
